# Antivirals for post-exposure prophylaxis of influenza: a systematic review and network meta-analysis

**DOI:** 10.1101/2024.05.28.24307995

**Authors:** Yunli Zhao, Ya Gao, Gordon Guyatt, Timothy M. Uyeki, Ping Liu, Ming Liu, Yanjiao Shen, Xiaoyan Chen, Shuyue Luo, Xingsheng Li, Rongzhong Huang, Qiukui Hao

## Abstract

**Background:** To support an update of WHO influenza guidelines, we performed a systematic review and network meta-analysis of the evidence on antiviral drugs for prophylaxis of influenza.

**Methods:** We analyzed randomized controlled trials published as of September 2023 on the efficacy and safety of antivirals compared to another antiviral or placebo, standard care, or no prophylaxis for prevention of symptomatic influenza. Paired reviewers independently screened studies, extracted data and assessed the risk of bias. We used frequentist random effects to perform network meta-analyses and assessed the certainty of evidence using the grading of recommendations assessment, development and evaluation (GRADE) methodology.

**Findings:** We included thirty-three trials of six antivirals (zanamivir, oseltamivir, laninamivir, baloxavir, amantadine, and rimantadine) that enrolled 19096 individuals. Zanamivir, oseltamivir, laninamivir and baloxavir probably achieve important reductions in symptomatic influenza in persons at high risk of severe disease (moderate certainty) when given promptly after exposure to seasonal influenza. These antivirals probably do not achieve important reductions in symptomatic influenza in persons at low risk of severe disease when given promptly after exposure to seasonal influenza (moderate certainty). Zanamivir, oseltamivir, laninamivir and baloxavir might achieve important reductions in symptomatic zoonotic influenza in persons exposed to novel influenza A viruses associated with severe disease in infected humans when given promptly after exposure (low certainty). These antivirals do not result in an important incidence of adverse events related to drugs or serious adverse events, with varying certainty of evidence.

**Interpretation:** Post-exposure prophylaxis with zanamivir, oseltamivir, laninamivir or baloxavir probably decreases the risk of symptomatic seasonal influenza in persons at high risk for severe disease after exposure to seasonal influenza viruses. Post-exposure prophylaxis with zanamivir, oseltamivir, laninamivir or baloxavir might reduce the risk of symptomatic zoonotic influenza after exposure to novel influenza A viruses associated with severe disease in infected humans.

**Funding:** WHO.

**Research in context:** *Evidence before this study:* Antivirals can be used to prevent influenza in people who have close contact to sick persons or animals infected with influenza viruses. Although previous reviews have found that antivirals (oseltamivir, zanamivir) are effective in preventing symptomatic influenza, these reviews assessed selected antivirals and did not rate the quality of evidence or consider the importance of effects in their interpretation. Additionally, a randomized controlled trial (RCT) of baloxavir for influenza post-exposure prophylaxis was not included in previous reviews.

*Added value of this study:* This systematic review and network meta-analyses of RCTs addressing antiviral prophylaxis against influenza was performed in support of a World Health Organisation (WHO) guidelines development group panel to formulate recommendations on use of antivirals for influenza. We present our analyses of the efficacy of antiviral prophylaxis to prevent symptomatic influenza for high or low-risk (non-high-risk) populations and for preventing symptomatic zoonotic influenza. We found moderate certainty evidence that zanamivir, oseltamivir, laninamivir and baloxavir all probably result in an important reduction in the risk of symptomatic seasonal influenza in high-risk persons when given promptly after exposure, but probably have no important effect for low-risk populations. Rimantadine probably has little or no effect on symptomatic seasonal influenza A virus infection (moderate certainty). Zanamivir, oseltamivir, laninamivir and baloxavir may decrease the risk of symptomatic zoonotic influenza (low certainty). The evidence for amantadine to prevent influenza A virus infection is limited. All of these antivirals have no important impact on adverse events.

*Implications of the available evidence:* The findings of this systematic review and network meta-analysis support use of zanamivir, oseltamivir, laninamivir or baloxavir for post-exposure prophylaxis of seasonal influenza in persons at high risk of severe influenza, and also provide some support for the use of these antivirals for post-exposure prophylaxis of zoonotic influenza. The systematic review did not support using these antivirals among low-risk populations for post-exposure prophylaxis of seasonal influenza and did not support the use of amantadine and rimantadine for preventing symptomatic influenza A virus infection.

## Introduction

Influenza is an acute respiratory viral illness characterized often by sudden onset of dry cough, nasal congestion, sore throat with or without fever and myalgia, headache, and weakness.^1^ These symptoms may be accompanied by pulmonary and extrapulmonary complications, such as neurological, cardiac and renal dysfunction. ^2,3^ Influenza viruses cause annual seasonal epidemics and unpredictable rare pandemics.^4^ All age groups can be affected by influenza, but young children, pregnant women, older adults, and individuals with chronic diseases are at a higher risk of severe disease. While most people recover from influenza without medical attention, the World Health Organization (WHO) estimated that 3 to 5 million cases of severe illness, and up to 650,000 influenza-related deaths occur annually worldwide.^4^

Annual vaccination is recommended to prevent influenza, especially for those at a higher risk of severe disease.^1^ However, influenza vaccine effectiveness can vary based on virus strain, population and year, ranging from 10% to 60% in the United States between 2004 and 2022.^5^ When influenza vaccine is unavailable or ineffective due to antigenic drift, viral immune evasion, or waning immunity; pre-exposure and post-exposure antiviral prophylaxis are strategies that can be important for influenza prevention, particularly in persons at increased risk of severe influenza complications.^6–8^

Previous systematic reviews have shown that antiviral post-exposure prophylaxis (PEP) with neuraminidase inhibitors can reduce the incidence of influenza,^9^ and the risk of symptomatic influenza in individuals and households.^10,11^ However, none of these reviews addressed all available antivirals from the different classes of drugs with different mechanisms of action, and no reviews assessed the quality of the supportive evidence. Moreover, previous systematic reviews did not include all randomized controlled trials (RCTs) of antivirals for prevention of influenza, including one large trial.^12^ To support an update of the WHO influenza guidelines,^13^ we conducted a systematic review and network meta-analysis to assess the efficacy and safety of all available approved antivirals for prophylaxis of influenza.

## Methods

This protocol for systematic review and network meta-analysis adheres to the Preferred Reporting Items for Systematic Review and Meta-Analysis Protocols (PRISMA-P) statement and is available on PROSPERO (CRD42023466450). The review team worked with the WHO guidelines panel to identify potential patient-important outcomes, pre-set subgroup analyses, and establish minimal important difference (MID) values.

### Literature search

To identify all RCTs of antiviral prophylaxis compared to placebo, standard care or another antiviral for prevention of influenza, we conducted a comprehensive search of published studies using EMBASE, MEDLINE, the Cochrane Central Registry of Controlled Trials, Cumulative Index to Nursing and Allied Health Literature (CINAHL), Global Health, Epistemonikos and ClincalTrials.gov databases as of 20 September 2023. We collaborated with an experienced medical librarian to refine our search strategy for each database without language restrictions. Appendix 1 presents the details of the search strategy. Additionally, we searched the reference lists of included studies and relevant systematic reviews to identify additional potentially eligible studies.

### Eligibility criteria and study selection

We included RCTs of direct-acting antivirals for prevention of influenza, including but not limited to neuraminidase inhibitors, viral polymerase complex inhibitors, cap-dependent endonuclease inhibitors, and matrix protein 2 ion channel inhibitors compared to placebo or standard care alone or to another antiviral in individuals exposed to influenza viruses. Eligible RCTs diagnosed influenza virus infection in respiratory specimens by reverse transcription polymerase chain reaction, rapid antigen test, or immunofluorescence assay.

Reviewers, working in pairs, independently performed study selection, including screening titles and abstracts, and evaluated full-text eligibility of potentially eligible studies using standardized forms. Reviewers resolved disagreements by discussion or, when necessary, by consultation with a third reviewer.

### Data extraction

For each eligible study, two reviewers independently extracted data on the following: study characteristics; patient characteristics; antiviral characteristics, specific influenza testing used to confirm influenza, follow-up time and all potentially important patient outcomes. Potentially important patient outcomes included data on events or time to symptom onset; asymptomatic and/or symptomatic infection, duration of symptoms, admission to hospital, length of hospitalization, progression to invasive mechanical ventilation, admission to an intensive care unit (ICU), length of mechanical ventilation, progression of disease severity, emergence of antiviral resistance, all-cause mortality, adverse events related to antivirals, and serious adverse events.

### Risk of bias assessment

For included trials that used individual randomization, reviewers independently assessed the risk of bias using the modified Cochrane risk of bias instrument (Appendix 2).^14,15^ For trials using cluster randomization, reviewers independently assessed the risk of bias using the Cochrane risk of bias instrument 2 (Appendix 2).^16^

### Statistical analysis

We performed network meta-analyses using the “netmeta” package of R version 4.0.2.^17^ We used frequentist random-effects to estimate the relative effect of all interventions, a design-by-treatment interaction model (global test) to assess the coherence assumption for the entire network, and node-splitting methods to assess the local incoherence between direct and indirect estimates in every closed loop of evidence.^18^

We performed pairwise meta-analysis using the “meta” and “metafor” package of R version 4.0.2 and applied the Hartung-Knapp-Sidik-Jonkman (HKSJ) random effects model to synthesize the data for pairwise meta-analyses when there were fewer than twenty studies included; otherwise, we used the DerSimonian and Laird (DL) random effects model.^19^

For binary outcomes, we calculated relative risk (RR) and the associated 95% confidence intervals (CIs) and assessed the risk differences (RD) by applying the median in the control group from eligible RCTs as the baseline mean.^20^ When the control event rate was <1%, we calculated the pooled absolute RD and its CI directly. For the cluster RCTs that failed to conduct appropriate analyses and did not report their effective sample sizes, we used the intracluster correlation coefficient (ICC=0.02 in main analyses and ICC=0.10 as one of sensitivity analyses) and the number of clusters to recalculate the design effects and the number of events.^16^ We reported the adjusted sample sizes and numbers of events in the meta-analyses. For any zero number cell, we added 0.5 to the cell with no event.^21^

### Subgroup analysis and investigation of heterogeneity

We assessed heterogeneity between studies by using restricted maximum likelihood (REML) models to calculate τ^2^ and I^2^ ^22^ and by visually inspecting the forest plots for differences in magnitude. We conducted within-trial comparisons when data were available and if not available between-trial comparisons if there were at least two studies in each subgroup. If there was potential subgroup effect (p<0.10), we used the Instrument for the Credibility of Effect Modification Analyses (ICEMAN) to assess the credibility of the subgroup effect.^23^ Our planned subgroup analyses included influenza virus type, age, exposure status, source of infection, influenza vaccination status, and disease severity. Appendix 3 details our prior hypotheses and the anticipated direction of effects.

### Publication bias

When there were ten or more eligible studies, we used funnel plots, Egger’s (for continuous variable), and Harbord tests (for discontinuous variable)^24^ to assess publication bias.

### Certainty of evidence

We used the GRADE (Grading of Recommendations Assessment, Development and Evaluation) approach to assess the certainty of evidence for network meta-analysis.^25,26^ We rated the overall certainty of evidence in absolute effects. When using RR as the relative effect measure, we calculated absolute effects using the relative effect estimates and the baseline risk estimates. We used the median rate in the placebo or standard care group of participants in eligible RCTs as the baseline risk. For the development of symptomatic zoonotic influenza following exposure to animals or persons infected with novel influenza A viruses associated with severe disease and high mortality in infected humans [e.g., avian influenza A(H5N1), A(H5N6), A(H7N9) viruses], the WHO guidelines panel estimated the baseline risk of symptomatic zoonotic influenza to be 3%, with 80% of patients with symptomatic zoonotic influenza requiring hospital admission, with 30% mortality. Appendix 4 details the methods for rating the certainty of evidence.

To assess imprecision for each outcome, we collaborated with the WHO guideline panel to establish MID values as the thresholds for assessing important patient outcomes. The panel established MID thresholds of 0.3% for all-cause mortality, 1.5% for hospitalization, 1% for drug-related adverse events and 0.5% for serious drug-related adverse events. For antiviral prophylaxis of lab-confirmed symptomatic seaonal influenza, the MID threshold was 5.5% for low-risk populations and 3% for high-risk populations. The panel defined the high-risk population based on the 2022 WHO influenza guidelines (Appendix 5);^13^ low-risk were non-high-risk persons.

## Results

### Description of included studies

Figure 1 shows the details of the study selection process. We identified 11,845 publications through database research and 18 publications from relevant reviews, of which 434 studies proved potentially eligible during the screening of titles and abstracts, and 33 studies proved eligible on full-text review.^12,27–58^ These studies included a total of 19,096 individuals with a mean age from 6.75 to 81.15 years.

**Figure 1.**
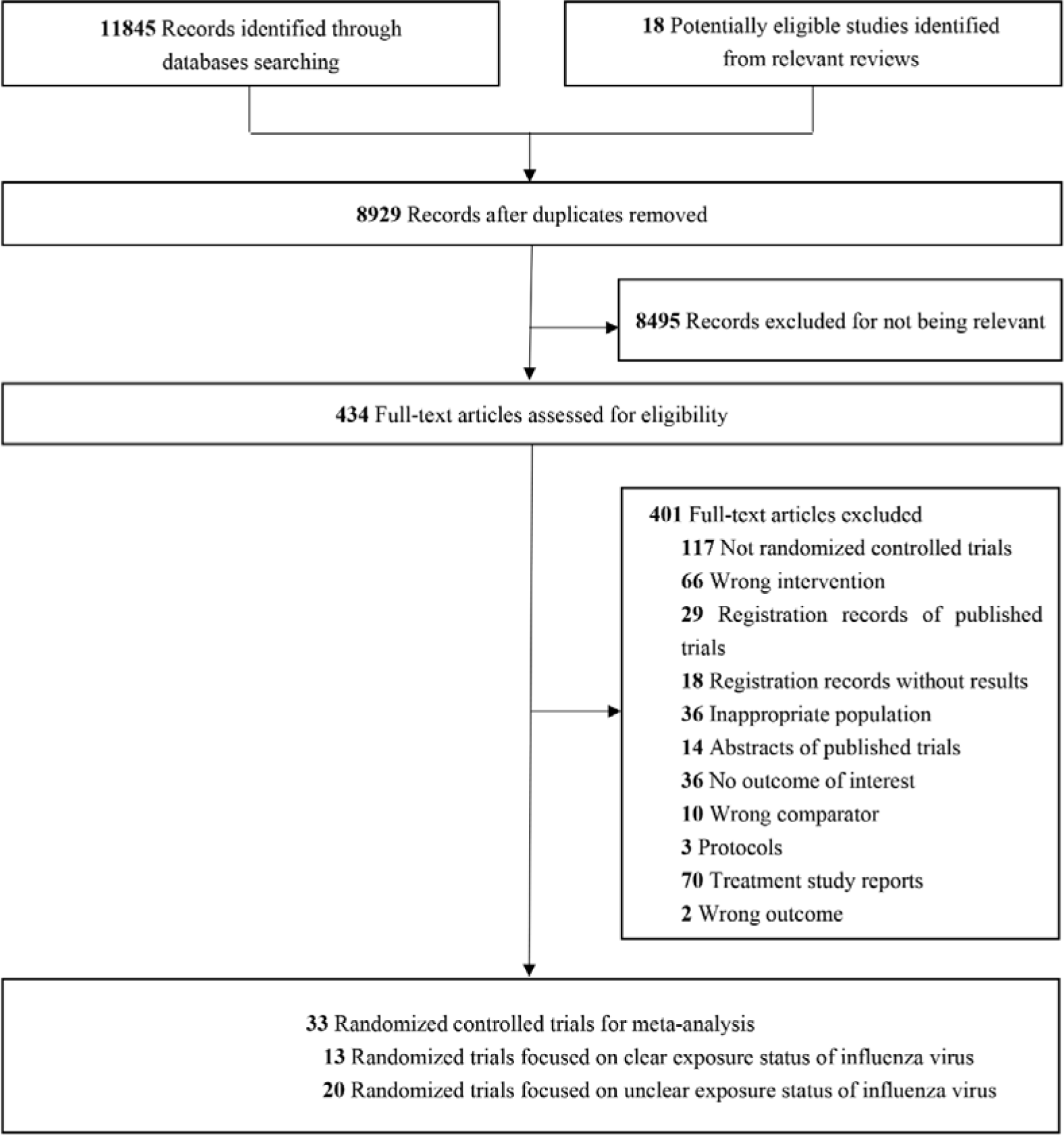
PRISMA flow diagram of studies included in the review.

Among the eligible studies, 13 trials assessed antivirals for PEP against seasonal or pandemic influenza for the populations with clear definition of exposure to influenza virus (e.g., close contact with patients with laboratory-confirmed influenza or influenza-like illness),^12,29,34–38,42,43,46,48,51,55^ and 20 trials assessed antiviral prophylaxis for populations with unclear definition of exposure status or pre-exposure prophylaxis against influenza (e.g., influenza season or outbreak in community or nursing home),^27,28,30–33,39–41,44,45,47,49,50,52–54,56–58^ and no trials were identified that assessed antivirals for prevention of human-to-human or animal-to-human transmission of novel influenza A viruses (zoonotic influenza). Of the 33 eligible trials, 7 used a cluster (household or nursing home) as the randomized unit ^29,34,35,46,48,54,55^ and 26 randomized individual participants.^27,28,30–33,39–41,44,45,47,49,50,52–54,56–58^ Appendix 6 presents characteristics of the included studies.

The included RCTs addressed 6 antivirals for prophylaxis of influenza: zanamivir, oseltamivir, laninamivir, baloxavir, amantadine, and rimantadine. Most of studies were rated at low risk of bias. Appendix 7 provides details of the risk of bias assessment for each trial.

Of thirteen trials that assessed the efficacy of antivirals for PEP against seasonal or pandemic influenza, ten trials studied antiviral prophylaxis of individuals started within 48 hours of exposure to a symptomatic index patient,^12,35,37,38,42,43,46,48,51,55^ one trial studied antiviral prophylaxis started as soon as possible after exposure to a symptomatic person with influenza,^34^ another trial assessed antiviral prophylaxis started up to 4 days after exposure,^36^ while one trial lacked clarity on the timing of antiviral prophylaxis after exposure to influenza.^29^ Among these trials of antiviral PEP of exposed persons, 8 trials also administered antiviral treatment to symptomatic index patients,^12,34,35,37,38,46,48,51^ in 4 trials the index patients did not receive antiviral treatment,^42,43,48,55^ and one did not specify whether index patients received antiviral treatment.^29^ The duration of antiviral PEP of exposed participants in the included trials ranged from one day^12,51^ to 10 days.^29,35,46,48,53^ For 20 trials that assessed pre-exposure antiviral prophylaxis, the trials usually initiated antiviral prophylaxis during influenza season or during an institutional influenza outbreak for 14 days ^27,54,57^ to 56 days.^47^

### Outcomes

Figure 2 presents the network analysis plots for all lab-confirmed symptomatic influenza (Figure 2a) and lab-confirmed influenza (Figure 2b). Other network analysis plots, forest plots, direct, indirect and network estimates, and details of the GRADE assessment for each outcome are presented in Appendices 8-12. Most of outcomes had no serious concerns for incoherence (Appendi× 10, 13) and heterogeneity (Appendix 14). No analyses suggested potential publication bias (Appendix 15).

**Figure 2.**
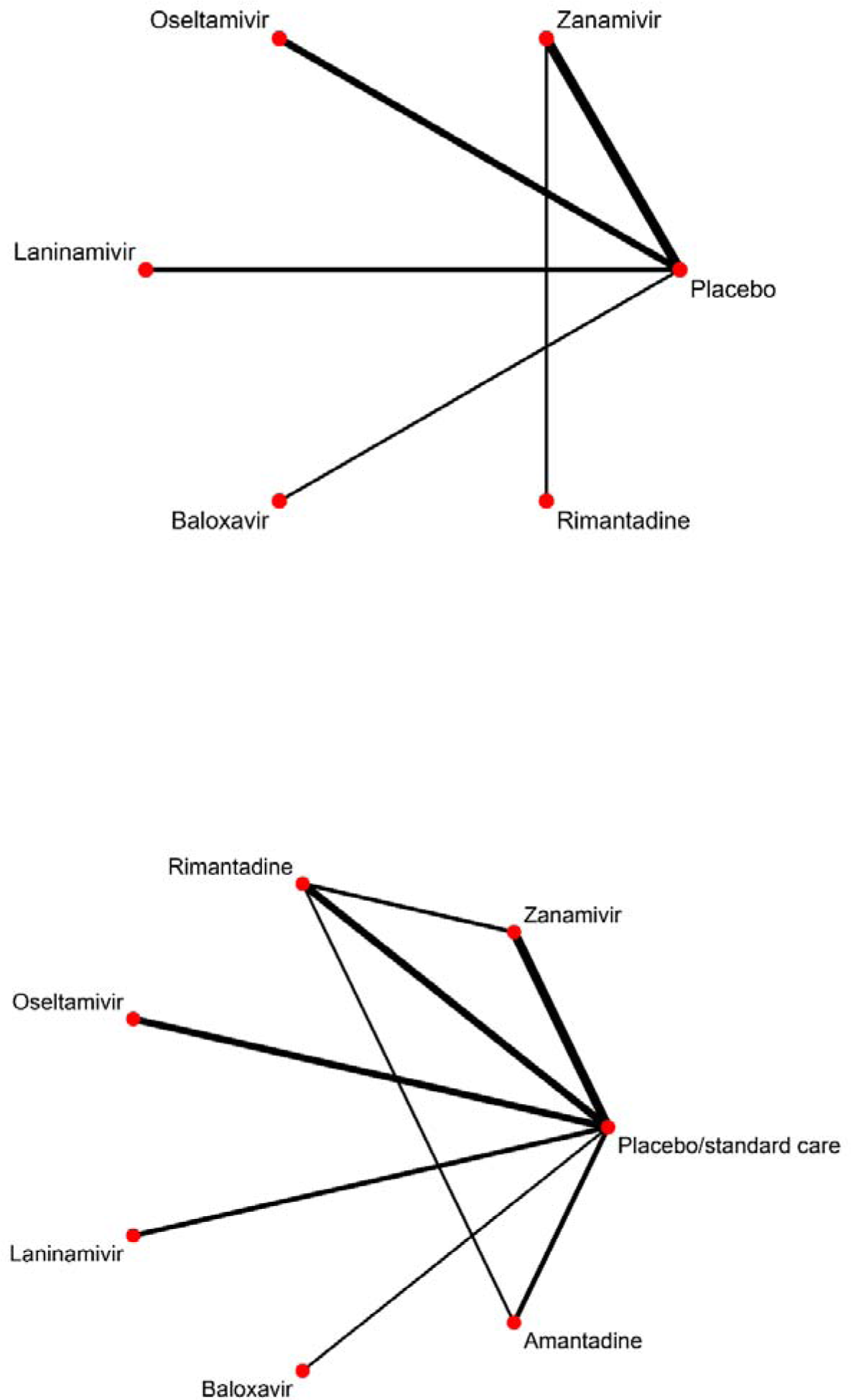
a Network plot of all included studies for lab-confirmed symptomatic influenza; b Network plot of all included studies for lab-confirmed influenza.

### Seasonal influenza

#### Symptomatic influenza (lab-confirmed)

Nineteen trials, including 15645 individuals, reported symptomatic influenza. All relative effect estimates come from direct comparisons between antivirals and placebo or standard care. In comparison to placebo or standard care, all antivirals, except for amantadine (no data) and rimantadine, had similar RR estimates ranging from 0.35 to 0.43 with 95% CIs that did not include no effect, indicating a reduction in the risk of symptomatic influenza. The RR for rimantadine for symptomatic influenza A virus infection was 0.76 (95% CI: 0.28 to 2.06) (Appendix 11-12).

For populations with low-risk of severe influenza such as admission to hospital or all-cause mortality, the effect of the drugs (zanamivir, oseltamivir, laninamivir, baloxavir and rimantadine) in reducing symptomatic influenza fell below the threshold of importance as defined by MIDs (relative risk estimates from 0.35 to 0.76 and absolute risk reductions ranged from 19 fewer to 51 fewer per 1000). For populations with a high risk of severe influenza, zanamivir, oseltamivir, laninamivir, and baloxavir all probably have an important effects in reducing symptomatic influenza (moderate certainty). In contrast, rimantadine may have little or no effect on symptomatic influenza A virus infection (low certainty). Whether amantadine reduces the development of symptomatic influenza A virus infection is very uncertain (Table 1).

#### Influenza and asymptomatic influenza virus infection (lab-confirmed)

Thirty-three trials with 19,096 individuals reported influenza virus infection regardless of symptoms. Compared to placebo or standard care, all antivirals had a similar effect showing a decrease in the risk of influenza virus infection, with relative risk estimates from 0.46 to 0.58 and absolute risk reductions from 96 fewer to 74 fewer per 1000. Oseltamivir, laninamivir, and baloxavir all probably decrease the risk of influenza virus infection, and amantadine probably decreases the risk of influenza A virus infection (moderate certainty, Table 1 and Appendix 7). Zanamivir possibly decreases the risk of influenza virus infection and rimantadine possibly decreases the risk of influenza A virus infection (low certainty). Antivirals reduced symptomatic influenza, driving the results for all influenza (symptomatic and asymptomatic infection). In contrast, antivirals probably have little or no effect on prevention of asymptomatic influenza virus infection (moderate certainty, Appendix 9-12).

#### Admission to hospital

Four studies with 3434 participants reported admission to hospital for the comparison of oseltamivir and placebo. Oseltamivir probably has little or no effect on admission to hospital (moderate certainty). No data were available on admission to hospital for other antiviral drugs (Table 1).

#### All-cause mortality

As the baseline event rate was lower than 1%, we calculated the risk difference for all-cause mortality. Fifteen studies with 10068 participants provided evidence that zanamivir, oseltamivir, laninamivir, and baloxavir all probably have little or no effect on all-cause mortality with absolute risk reductions from 0 fewer to 1 more per 1000 patients (high or moderate certainty). Whether amantadine or rimantadine reduces all-cause mortality from influenza A virus infection is very uncertain (Table 1).

#### Adverse events

Thirteen trials with 10838 participants reported adverse events related to drugs. Zanamivir, laninamivir, and rimantadine all probably result in few drug-related adverse events with relative risks ranging from 1.01 to 1.40 and absolute risks ranging from 3 to 14 more per 1000 (moderate certainty). Compared with placebo, baloxavir may have little or no effect on drug-related adverse events (6 more per 1000, 95% CI 22 fewer to 88 more, low certainty; Table 1).

Sixteen studies with 11755 participants reported serious adverse events. Compared to placebo, all antivirals may have little or no effect on serious adverse events with absolute risk increases ranging from 0 to 4 per 1000. The certainty of evidence was high for zanamivir; moderate for oseltamivir and rimantadine, and low for laninamivir, baloxavir, and amantadine (Table 1).

#### Zoonotic influenza

No RCTs were identified for antiviral PEP of persons exposed to symptomatic persons or to animals infected with novel influenza A viruses (zoonotic influenza) associated with severe disease and high mortality in infected humans. Therefore, we considered indirect evidence from trials of antiviral PEP for seasonal influenza. For populations exposed to novel influenza A viruses associated with severe disease and high mortality in infected humans, zanamivir, oseltamivir, laninamivir, and baloxavir may have an important effect in reducing development of symptomatic zoonotic influenza (low certainty). Whether amantadine or rimantadine reduce the development of symptomatic zoonotic influenza is very uncertain (Table 1). We infer that when using antivirals for PEP against zoonotic influenza, the adverse events are similar to those when using antivirals for PEP of seasonal influenza (Table 1).

Although the evidence for seasonal influenza raises the possibility that zanamivir, oseltamivir, laninamivir, and baloxavir may have important effects on admission to hospital and all-cause mortality for people exposed to novel influenza A viruses associated with severe disease and high mortality in infected humans. However, the certainty of evidence is very low (Table 1, Appendix 11-12).

### Sensitivity analyses and subgroup analysis

Our sensitivity analyses showed similar results to the primary analyses. No significant subgroup effects were found between different age groups and influenza vaccine status on symptomatic influenza (interaction p>0.10). When analyzing antiviral treatment in subgroups of index patients, a significant interaction (p=0.01) was found, but the subgroup effect was low according to ICMEAN criteria. Appendix 16-21 shows the details of the sensitivity and subgroup analyses.

## Discussion

In this systematic review and network metanalysis of RCTs of antiviral prophylaxis of influenza compared to placebo or standard care, we found that oseltamivir, laninamivir, zanamivir, and baloxavir all can reduce the risk of symptomatic seasonal influenza in high-risk persons exposed to a symptomatic close contact with influenza when antiviral prophylaxis is initiated promptly after exposure. However, PEP with these antivirals may have little or no important effects on reducing symptomatic seasonal influenza in low-risk populations. For populations exposed to novel influenza A viruses associated with severe disease in infected humans, prompt administration of PEP with oseltamivir, zanamivir, laninamivir, or baloxavir may reduce the risk of symptomatic zoonotic influenza. We did not find convincing evidence to suggest that antiviral prophylaxis provides important reductions in the risk of admission to hospital or all-cause mortality due to seasonal influenza. These antivirals may also have no important effects on adverse events or severe adverse events related to the drugs.

### Strength and limitations

This is the first network meta-analysis to assess antivirals for prophylaxis of influenza. We specified explicit eligibility criteria, conducted a comprehensive literature search for eligible studies and performed duplicate selection, data extraction, and risk of bias assessment. We assessed the effects of antivirals in preventing seasonal influenza for both low-risk and high-risk populations, and based on the WHO guideline panel’s discussions, incorporated data modeling for prevention of zoonotic influenza. The WHO panel provided MID values of importance to patients that we used to interpret the results and guide the ratings for imprecision. We performed within-trial subgroup analyses to explore possible effect modification according to age and influenza vaccination status and performed sensitivity analyses to assess risk of bias, missing data, exposures status to influenza viruses, and ICC values, all of which yielded results similar to our primary analyses.

Our review has limitations. First, data were unavailable to assess some outcomes identified by the WHO guidelines panel as important, including length of hospitalization, ICU admission, invasive mechanical ventilation, and influenza disease severity, and there were very limited data for pregnant people or infants aged <1 year. Second, although studies varied in the route of drug administration, dosage, and duration of PEP (Appendix 2), we combined them in the network meta-analysis. Third, we included three trials in which the index case had influenza-like illness because the results were unchanged when these studies were excluded in sensitivity analyses. Fourth, because no RCTs have been conducted of antivirals for PEP to prevent symptomatic zoonotic influenza, we used indirect evidence from RCTs of seasonal influenza as the evidence base for zoonotic influenza. Also, because the overall mortality for zoonotic influenza is unknown, we estimated the case fatality proportion for zoonotic influenza to be 30%, based upon sporadic human infections with different novel influenza A viruses associated with severe disease. This overestimates the mortality for other avian influenza A virus infections and swine influenza A virus infections of humans that are associated with lower disease severity.

### Comparison with prior work

Previous systematic reviews suggested that prophylaxis with antivirals is effective for preventing symptomatic influenza but did not assess the certainty of the evidence.^9–11,59–63^ Our findings are consistent with those of previous systematic reviews of no significant effect on asymptomatic influenza virus infection,^10,11^ mortality^59^ or serious adverse events.^10^ Previous reviews assessed zanamivir, oseltamivir and laninamivir but did not include one recent RCT for baloxavir. Our systematic review is updated and more comprehensive than prior reviews, provides ratings of the certainty of evidence, and is the first review to specify MIDs and thus to explicitly address the importance to patients of the intervention effects.

### Clinical and research implications

Our systematic review provides evidence for the clinical benefit and safety of antiviral prophylaxis to prevent symptomatic seasonal influenza in persons who are at high risk for severe influenza when started within 48 hours of exposure to a symptomatic person with influenza.

There are a number of important populations for which data from RCTs on post-exposure antiviral prophylaxis remain limited, including for exposed pregnant people, infants, persons with kidney dysfunction, liver disease, and other chronic medical conditions. Other gaps are in understanding the risk of infection and severe zoonotic influenza, and benefit of post-exposure antiviral prophylaxis in people exposed to animals or humans infected with novel influenza A viruses associated with severe disease and high mortality.

## Conclusions

Following exposure to persons with seasonal influenza, prompt initiation of PEP with oseltamivir, zanamivir, laninamivir or baloxavir all probably provide important reductions in the risk of symptomatic influenza in persons who are at high-risk for severe influenza. Similarly, based upon indirect evidence for seasonal influenza, prompt initiation of PEP with these antivirals may also provide important reductions in the risk of symptomatic zoonotic influenza.

## Supporting information

PRISMA_2020_checklist

## Authors’ contributions

Yunli Zhao, Gordon Guyatt and Qiukui Hao conceived and designed the study; Yunli Zhao, Ya Gao, Ping Liu, Ming Liu, Yanjiao Shen, Xiaoyan Chen, Shuyue Luo acquired data; Yunli Zhao, Ya Gao, Xingsheng Li, Rongzhong Huang and Qiukui Hao analyzed data; Yunli Zhao, Ya Gao, Gordon Guyatt, Timothy M. Uyeki and Qiukui Hao interpreted the data; Yunli Zhao and Qiukui Hao drafted the article; Gordon Guyatt and Timothy M. Uyeki critically revised the article; all authors reviewed the submitted version of manuscript and approved the final version of the manuscript.

## Data sharing

All data included were derived from publicly available documents cited in the references. Extracted data are available upon request to the corresponding author.

## Acknowledgments

We thank Rachel Couban (librarian at McMaster University) for helping with developing the search strategy. Yanjiao Shen acknowledges funding from China Scholarship Council.

