## Supplementary Appendix for "Antivirals for post-exposure prophylaxis of influenza: a systematic review and network meta-analysis"

Antiviral therapies for prophylaxis against influenza: a systematic review and network meta-analysis

Content of table

### Appendix 1 Search strategy

Ovid MEDLINE(R) ALL

1	exp Influenza, Human/	58097

2	exp Influenza A virus/	49553

3	exp Influenza B virus/	4644

4	exp Influenzavirus C/	363

5	(Influenza or flu or H1N1 or PH1N1 or H3N2 or AH1N1 or AH3N2 or H5N1 or H7N9).mp. [mp=title, book title, abstract, original title, name of substance word, subject heading word, floating sub-heading word, keyword heading word, organism supplementary concept word, protocol supplementary concept word, rare disease supplementary concept word, unique identifier, synonyms, population supplementary concept word, anatomy supplementary concept word]	140078

6	or/1-5	140084

7	Antiviral agents/	100530

8	Antiviral*.tw.	118002

9	(neuraminidase inhibitor* or NA inhibitor*).tw.	2443

10	Oseltamivir/ or Zanamivir/	3724

11	(oseltamivir or tamiflu or GS 4104 or GS4104 or GS-4104 or GS 4071 or GS4071 or GS-4071 or zanamivir or relenza or GG 167 or GG167 or GG-167 or CS-8958 or Dectova or Laninamivir or R-125489 or R125489 or R 125489 or Inavir or peramivir or BCX 1812 or BCX1812 or BCX-1812 or RWJ 270201 or RWJ270201 or RWJ-270201 or Rapivab or rapiacta).ti,ab.	5041

12	Viral Polymerase Complex Inhibitor*.tw.	0

13	(Favipiravir or T-705 or Avigan or FabiFlu or Pimodivir or VX-787 or JNJ-63623872 or AL-794 or ALS-033719 or ZSP1273 or Enisamium iodide or FAV00A or TG-1000 or GP681).ti,ab.	1339

14	matrix protein 2 ion channel inhibitor*.tw.	2

15	(Radavirsen or AVI-7100).ti,ab.	2

16	cap-dependent endonuclease inhibitor*.tw.	38

17	("Baloxavir marboxil" or Baloxavir or S-033188 or Xofluza).ti,ab.	274

18	(Umifenovir or Arbidol or Arbidole).ti,ab.	430

19	Amantadine/ or Rimantadine/	4282

20	(Amantadine or Symmetrel or Symetrel or Rimantadine or Flumadine or Roflual).ti,ab.	4429

21	or/7-20	178589

22	6 and 21	18091

23	randomized controlled trial.pt.	599492

24	controlled clinical trial.pt.	95418

25	randomized.ab.	616237

26	placebo.ab.	241273

27	drug therapy.fs.	2621016

28	randomly.ab.	416158

29	trial.ti.	292288

30	groups.ab.	2566299

31	or/23-30	5605395

32	(animals not (humans and animals)).sh.	5119274

33	31 not 32	4891800

34	22 and 33	6314

Ovid Embase

1	exp Influenza/ or Influenza virus/	121021

2	exp Influenza A virus/ or exp Influenza A virus/	20306

3	exp Influenza B/ or exp Influenza B virus/	6376

4	exp Influenza C/ or exp Influenza C virus/	240

5	(Influenza or flu or H1N1 or PH1N1 or H3N2 or AH1N1 or AH3N2 or H5N1 or H7N9).mp. [mp=title, abstract, heading word, drug trade name, original title, device manufacturer, drug manufacturer, device trade name, keyword heading word, floating subheading word, candidate term word]	203693

6	or/1-5	203693

7	Antivirus agent/	97344

8	Antiviral*.tw.	155192

9	(neuraminidase inhibitor* or NA inhibitor*).tw.	3010

10	Sialidase inhibitor/ or Oseltamivir/ or Zanamivir/ or Laninamivir/ or Peramivir/	17267

11	(oseltamivir or tamiflu or "GS 4104" or GS4104 or GS-4104 or "GS 4071" or GS4071 or GS-4071 or zanamivir or relenza or "GG 167" or GG167 or GG-167 or CS-8958 or Dectova or Laninamivir or R-125489 or R125489 or "R 125489" or Inavir or peramivir or "BCX 1812" or BCX1812 or BCX-1812 or "RWJ 270201" or RWJ270201 or RWJ-270201 or Rapivab or rapiacta).ti,ab.	7147

12	Viral Polymerase Complex Inhibitor*.tw.	0

13	Favipiravir/ or Pimodivir/ or (Favipiravir or T-705 or Avigan or FabiFlu or Pimodivir or VX-787 or JNJ-63623872 or AL-794 or ALS-033719 or ZSP1273 or Enisamium iodide or FAV00A or TG-1000 or GP681).ti,ab.	5003

14	matrix protein 2 ion channel inhibitor*.tw.	1

15	Radavirsen/ or (Radavirsen or AVI-7100).ti,ab.	15

16	cap-dependent endonuclease inhibitor*.tw.	49

17	Baloxavir marboxil/ or ("Baloxavir marboxil" or Baloxavir or S-033188 or Xofluza).ti,ab.	538

18	Umifenovir/ or (Umifenovir or Arbidol or Arbidole).ti,ab.	1836

19	Amantadine/ or Rimantadine/	17849

20	(Amantadine or Symmetrel or Symetrel or Rimantadine or Flumadine or Roflual).ti,ab.	5729

21	or/7-20	229508

22	6 and 21	29821

23	Randomized controlled trial/	782084

24	Controlled clinical study/	470919

25	random$.ti,ab.	1972415

26	randomization/	98334

27	intermethod comparison/	300395

28	placebo.ti,ab.	364870

29	(compare or compared or comparison).ti.	604471

30	((evaluated or evaluate or evaluating or assessed or assess) and (compare or compared or comparing or comparison)).ab.	2773393

31	(open adj label).ti,ab.	108525

32	((double or single or doubly or singly) adj (blind or blinded or blindly)).ti,ab.	273606

33	double blind procedure/	210398

34	parallel group$1.ti,ab.	32087

35	(crossover or cross over).ti,ab.	124492

36	((assign$ or match or matched or allocation) adj5 (alternate or group$1 or intervention$1 or patient$1 or subject$1 or participant$1)).ti,ab.	414740

37	(assigned or allocated).ti,ab.	490042

38	(controlled adj7 (study or design or trial)).ti,ab.	449985

39	(volunteer or volunteers).ti,ab.	282271

40	human experiment/	641156

41	trial.ti.	400728

42	or/23-41	6316622

43	(random$ adj sampl$ adj7 ("cross section$" or questionnaire$1 or survey$ or database$1)).ti,ab. not (comparative study/ or controlled study/ or randomi?ed controlled.ti,ab. or randomly assigned.ti,ab.)	9594

44	Cross-sectional study/ not (randomized controlled trial/ or controlled clinical study/ or controlled study/ or randomi?ed controlled.ti,ab. or control group$1.ti,ab.)	360295

45	(((case adj control$) and random$) not randomi?ed controlled).ti,ab.	21530

46	(Systematic review not (trial or study)).ti.	258944

47	(nonrandom$ not random$).ti,ab.	18933

48	"Random field$".ti,ab.	2959

49	(random cluster adj3 sampl$).ti,ab.	1578

50	(review.ab. and review.pt.) not trial.ti.	1128667

51	"we searched".ab. and (review.ti. or review.pt.)	49232

52	"update review".ab.	136

53	(databases adj4 searched).ab.	62280

54	(rat or rats or mouse or mice or swine or porcine or murine or sheep or lambs or pigs or piglets or rabbit or rabbits or cat or cats or dog or dogs or cattle or bovine or monkey or monkeys or trout or marmoset$1).ti. and animal experiment/	1219722

55	Animal experiment/ not (human experiment/ or human/)	2561951

56	or/43-55	4337577

57	42 not 56	5573353

58	22 and 57	3027

OVID EBM Reviews - Cochrane Central Register of Controlled Trials

1	exp Influenza, Human/	3241

2	exp Influenza A virus/	1021

3	exp Influenza B virus/	320

4	exp Influenzavirus C/	0

5	(Influenza or flu or H1N1 or PH1N1 or H3N2 or AH1N1 or AH3N2 or H5N1 or H7N9).mp. [mp=title, original title, abstract, floating sub-heading word, mesh headings, heading words, keyword]	10694

6	or/1-5	10694

7	Antiviral agents/	4914

8	Antiviral*.tw.	6923

9	(neuraminidase inhibitor* or NA inhibitor*).tw.	145

10	Oseltamivir/ or Zanamivir/	348

11	(oseltamivir or tamiflu or "GS 4104" or GS4104 or GS-4104 or "GS 4071" or GS4071 or GS-4071 or zanamivir or relenza or "GG 167" or GG167 or GG-167 or CS-8958 or Dectova or Laninamivir or R-125489 or R125489 or "R 125489" or Inavir or peramivir or "BCX 1812" or BCX1812 or BCX-1812 or "RWJ 270201" or RWJ270201 or RWJ-270201 or Rapivab or rapiacta).ti,ab.	700

12	Viral Polymerase Complex Inhibitor*.tw.	0

13	(Favipiravir or T-705 or Avigan or FabiFlu or Pimodivir or VX-787 or JNJ-63623872 or AL-794 or ALS-033719 or ZSP1273 or Enisamium iodide or FAV00A or TG-1000 or GP681).ti,ab.	310

14	matrix protein 2 ion channel inhibitor*.tw.	0

15	(Radavirsen or AVI-7100).ti,ab.	3

16	cap-dependent endonuclease inhibitor*.tw.	4

17	("Baloxavir marboxil" or Baloxavir or S-033188 or Xofluza).ti,ab.	59

18	(Umifenovir or Arbidol or Arbidole).ti,ab.	71

19	Amantadine/ or Rimantadine/	485

20	(Amantadine or Symmetrel or Symetrel or Rimantadine or Flumadine or Roflual).ti,ab.	913

21	or/7-20	11661

22	6 and 21	1277

23	randomized controlled trial.pt.	0

24	controlled clinical trial.pt.	0

25	randomized.ab.	680359

26	placebo.ab.	332394

27	drug therapy.fs.	261711

28	randomly.ab.	309521

29	trial.ti.	410998

30	groups.ab.	576574

31	or/23-30	1361436

32	(animals not (humans and animals)).sh.	2671

33	31 not 32	1359098

34	22 and 33	1006

Ovid Global Health

1	exp Influenza/ or Influenza viruses/	40928

2	exp Influenza A virus/ or exp Influenza A virus/	22916

3	exp Influenza B/ or exp Influenza B virus/	3545

4	exp Influenza C/ or exp Influenza C virus/	180

5	(Influenza or flu or H1N1 or PH1N1 or H3N2 or AH1N1 or AH3N2 or H5N1 or H7N9).mp. [mp=abstract, title, original title, heading words, cabicodes words]	50678

6	or/1-5	50678

7	Antiviral agents/	115951

8	Antiviral*.tw.	138051

9	(neuraminidase inhibitor* or NA inhibitor*).tw.	1090

10	Sialidase inhibitors/ or Oseltamivir/ or Zanamivir/ or Laninamivir/ or Peramivir/	1965

11	(oseltamivir or tamiflu or "GS 4104" or GS4104 or GS-4104 or "GS 4071" or GS4071 or GS-4071 or zanamivir or relenza or "GG 167" or GG167 or GG-167 or CS-8958 or Dectova or Laninamivir or R-125489 or R125489 or "R 125489" or Inavir or peramivir or "BCX 1812" or BCX1812 or BCX-1812 or "RWJ 270201" or RWJ270201 or RWJ-270201 or Rapivab or rapiacta).ti,ab.	2460

12	Viral Polymerase Complex Inhibitor*.tw.	0

13	Favipiravir/ or (Favipiravir or T-705 or Avigan or FabiFlu or Pimodivir or VX-787 or JNJ-63623872 or AL-794 or ALS-033719 or ZSP1273 or Enisamium iodide or FAV00A or TG-1000 or GP681).ti,ab.	682

14	matrix protein 2 ion channel inhibitor*.tw.	0

15	(Radavirsen or AVI-7100).ti,ab.	1

16	cap-dependent endonuclease inhibitor*.tw.	17

17	("Baloxavir marboxil" or Baloxavir or S-033188 or Xofluza).ti,ab.	113

18	(Umifenovir or Arbidol or Arbidole).ti,ab.	185

19	Amantadine/ or Rimantadine/	551

20	(Amantadine or Symmetrel or Symetrel or Rimantadine or Flumadine or Roflual).ti,ab.	708

21	or/7-20	138737

22	6 and 21	8017

23	exp randomized controlled trials/	61880

24	(randomized controlled trial or random* or blind* or placebo*).mp. [mp=abstract, title, original title, heading words, cabicodes words]	322171

25	23 or 24	322171

26	22 and 25	590

CINAHL

| # | Query | Limiters/Expanders | Results |
| --- | --- | --- | --- |
| S36 | S23 AND S26 AND S35 | Search modes - Boolean/Phrase | 391 |
| S35 | S27 OR S28 OR S29 OR S30 OR S31 OR S32 OR S33 OR S34 | Search modes - Boolean/Phrase | 27,192 |
| S34 | TI ( matrix protein 2 ion channel inhibitor* OR Radavirsen or AVI-7100 OR cap-dependent endonuclease inhibitor* OR "Baloxavir marboxil" or Baloxavir or S-033188 or Xofluza OR Umifenovir or Arbidol or Arbidole OR Amantadine or Symmetrel or Symetrel or Rimantadine or Flumadine or Roflual ) OR AB ( matrix protein 2 ion channel inhibitor* OR Radavirsen or AVI-7100 OR cap-dependent endonuclease inhibitor* OR "Baloxavir marboxil" or Baloxavir or S-033188 or Xofluza OR Umifenovir or Arbidol or Arbidole OR Amantadine or Symmetrel or Symetrel or Rimantadine or Flumadine or Roflual ) | Search modes - Boolean/Phrase | 711 |
| S33 | (MH "Amantadine") | Search modes - Boolean/Phrase | 587 |
| S32 | TI ( Viral Polymerase Complex Inhibitor* OR Favipiravir or T-705 or Avigan or FabiFlu or Pimodivir or VX-787 or JNJ-63623872 or AL-794 or ALS-033719 or ZSP1273 or Enisamium iodide or FAV00A or TG-1000 or GP681 ) OR AB ( Viral Polymerase Complex Inhibitor* OR Favipiravir or T-705 or Avigan or FabiFlu or Pimodivir or VX-787 or JNJ-63623872 or AL-794 or ALS-033719 or ZSP1273 or Enisamium iodide or FAV00A or TG-1000 or GP681 ) | Search modes - Boolean/Phrase | 179 |
| S31 | TI ( oseltamivir or tamiflu or "GS 4104" or GS4104 or GS-4104 or "GS 4071" or GS4071 or GS-4071 or zanamivir or relenza or "GG 167" or GG167 or GG-167 or CS-8958 or Dectova or Laninamivir or R-125489 or R125489 or "R 125489" or Inavir or peramivir or "BCX 1812" or BCX1812 or BCX-1812 or "RWJ 270201" or RWJ270201 or RWJ-270201 or Rapivab or rapiacta ) OR AB ( oseltamivir or tamiflu or "GS 4104" or GS4104 or GS-4104 or "GS 4071" or GS4071 or GS-4071 or zanamivir or relenza or "GG 167" or GG167 or GG-167 or CS-8958 or Dectova or Laninamivir or R-125489 or R125489 or "R 125489" or Inavir or peramivir or "BCX 1812" or BCX1812 or BCX-1812 or "RWJ 270201" or RWJ270201 or RWJ-270201 or Rapivab or rapiacta ) | Search modes - Boolean/Phrase | 1,256 |
| S30 | (MH "Oseltamivir") | Search modes - Boolean/Phrase | 787 |
| S29 | TI(neuraminidase inhibitor* or NA inhibitor*) OR AB(neuraminidase inhibitor* or NA inhibitor*) | Search modes - Boolean/Phrase | 397 |
| S28 | TI Antiviral* OR AB Antiviral* | Search modes - Boolean/Phrase | 11,069 |
| S27 | (MH "Antiviral Agents") | Search modes - Boolean/Phrase | 19,889 |
| S26 | S24 OR S25 | Search modes - Boolean/Phrase | 33,506 |
| S25 | TI ( Influenza or flu or H1N1 or PH1N1 or H3N2 or AH1N1 or AH3N2 or H5N1 or H7N9 ) OR AB ( Influenza or flu or H1N1 or PH1N1 or H3N2 or AH1N1 or AH3N2 or H5N1 or H7N9 ) | Search modes - Boolean/Phrase | 29,072 |
| S24 | (MH "Influenza+") OR (MH "Influenza A Virus+") OR (MH "Influenzavirus C") OR (MH "Influenza B Virus") | Search modes - Boolean/Phrase | 22,039 |
| S23 | S22 NOT S21 | Search modes - Boolean/Phrase | 971,896 |
| S22 | S1 OR S2 OR S3 OR S4 OR S5 OR S6 OR S7 OR S8 OR S9 OR S10 OR S11 OR S12 OR S13 OR S14 OR S15 | Search modes - Boolean/Phrase | 1,019,541 |
| S21 | S19 NOT S20 | Search modes - Boolean/Phrase | 213,830 |
| S20 | MH (human) | Search modes - Boolean/Phrase | 2,696,267 |
| S19 | S16 OR S17 OR S18 | Search modes - Boolean/Phrase | 247,788 |
| S18 | TI (animal model*) | Search modes - Boolean/Phrase | 3,563 |
| S17 | MH (animal studies) | Search modes - Boolean/Phrase | 153,253 |
| S16 | MH animals+ | Search modes - Boolean/Phrase | 103,495 |
| S15 | AB (cluster W3 RCT) | Search modes - Boolean/Phrase | 498 |
| S14 | MH (crossover design) OR MH (comparative studies) | Search modes - Boolean/Phrase | 478,421 |
| S13 | AB (control W5 group) | Search modes - Boolean/Phrase | 145,440 |
| S12 | PT (randomized controlled trial) | Search modes - Boolean/Phrase | 151,971 |
| S11 | MH (placebos) | Search modes - Boolean/Phrase | 13,800 |
| S10 | MH (sample size) AND AB (assigned OR allocated OR control) | Search modes - Boolean/Phrase | 4,424 |
| S9 | TI (trial) | Search modes - Boolean/Phrase | 183,598 |
| S8 | AB (random*) | Search modes - Boolean/Phrase | 400,359 |
| S7 | TI (randomised OR randomized) | Search modes - Boolean/Phrase | 142,994 |
| S6 | MH cluster sample | Search modes - Boolean/Phrase | 5,250 |
| S5 | MH pretest‐posttest design | Search modes - Boolean/Phrase | 53,382 |
| S4 | MH random assignment | Search modes - Boolean/Phrase | 80,504 |
| S3 | MH single‐blind studies | Search modes - Boolean/Phrase | 16,026 |
| S2 | MH double‐blind studies | Search modes - Boolean/Phrase | Display |
| S1 | MH randomized controlled trials | Search modes - Boolean/Phrase | Display |

### Appendix 2 Risk of bias tool

The modified Cochrane instrument evaluates eight domains, including random sequence generation; allocation concealment; blinding of participants, healthcare providers, data collectors and outcome assessors; incomplete outcome data (if the rate of missing data was less than 10%, we judged it as low risk of bias); and other potential sources of bias (e.g., early trial discontinuation). We rated each domain at the outcome level as either: definitely or probably low risk of bias (low risk of bias), probably or definitely high risk of bias (high risk of bias).

The Cochrane risk of bias instrument 2 evaluates six domains, including randomization, the timing of identification and recruitment of participants, intended interventions, incomplete outcome data, measurement of the outcome and selection of the reported result. We rated each domain at the outcome level as either: low risk of bias, unclear risk of bias and high risk of bias. When there were more than two domains with high risk of bias, we rated the outcome of that study as high risk of bias.

### Appendix 3 Prior hypotheses and the anticipated direction of effects for subgroup analysis.

We planned to conduct subgroup analyses with hypothesized direction of effect as follows:

1. Influenza virus type (antivirals have greater effectiveness for prevention of pandemic influenza than for seasonal or zoonotic influenza);
2. Age (antivirals are more effective for prevention of influenza in non-older adults and adolescents than other age groups exposed to influenza such as young children aged <2 years, children aged 2-12 years, and older adults (≥65 years);
3. Exposure status (antivirals are more effective in exposed persons who utilized personal protective measures (e.g., facemasks) than in persons with unprotected exposures to influenza viruses;
4. Exposure to source of infection (antivirals are more effective for persons exposed to ill humans with influenza than for persons exposed to animal sources infected with influenza A viruses);
5. Influenza vaccination status (antivirals are more effective in vaccinated persons exposed to influenza viruses than in unvaccinated persons);
6. Disease severity (antivirals are more effective at reducing severe influenza in persons exposed to influenza viruses who are at high risk for severe disease, such as people who are pregnant or up to 2 weeks postpartum, extreme obesity (BMI≥40), or those have underlying health conditions (e.g., chronic respiratory, cardiovascular disease, immunocompromised) than in low risk persons)

When there were only between-trial data but ten or more studies were available, we performed random effects meta-regressions to examine the impact of continuous variables (e.g., age) on the effect of antivirals for preventing influenza.

### Appendix 4 Methods for rating the certainty of evidence.

Following GRADE guidance, we rated the certainty of evidence for direct estimates of antiviral post-exposure prophylaxis as very low, low, moderate, or high by assessing limitations such as risk of bias, inconsistency, indirectness, and publication bias [31]. When outcome data were missing, we conducted a complete case analysis as the primary analysis. If the results of the primary analysis indicated a statistically significant effect of antiviral prophylaxis, we planned to evaluate the robustness by performing a plausible worst case sensitivity analysis for each outcome [32]. If the worst-case analyses significantly influenced the observed effect, we planned to rate down the certainty of evidence for risk of bias due to missing data.

To assess the evidence for indirect estimates, we focused on the dominant first order loop and rated the certainty of indirect evidence as the lowest certainty of the direct comparisons informing that dominant loop. When the distribution of plausible effect modification differed in the contributing direct comparisons, we further rated down indirect comparisons for intransitivity.

For the network analysis estimates, we started with the certainty of evidence that dominates the comparison (direct or indirect estimates). When direct and indirect estimates contributed similarly to the network estimate, we chose the higher of the two certainties. When incoherence existed between the direct and indirect estimates, we planned to rate down the certainty of evidence for incoherence and used the estimate with the higher certainty as the best estimate for effect.

We followed GRADE guidance in reporting results.^1^ We developed the summary of finding tables using MAGICapp [https://app.magicapp.org], providing both relative and absolute effects and including plain language summaries with wording following GRADE guidance.^1,2^

### Appendix 5 Character population at high risk of severe disease.

The high-risk population included individuals with conditions such as chronic lung disease, cardiac disease, hematologic disorders, kidney or liver disorders, metabolic disease, immunosuppression, pregnant persons and those up to 2 weeks postpartum, children younger than 59 months, persons 65 years and older, people with a body mass index of 40 or higher, and those younger than 19 years of age who are on long-term aspirin- or salicylate-containing medications. All persons exposed to symptomatic humans or to animals infected with novel influenza A viruses known to be associated with high mortality in humans, or exposed to influenza A viruses circulating among animals with unknown disease severity in infected humans, were considered to be at high risk of severe disease.

### Appendix 6 Basic characteristics of eligible studies

| Study | Publication status  Registration | Randomization design | Patients randomized | Country | Mean age,  age range (years) | Male (%) | Exposure status | Vaccinated (%) | Treatment for patients | Treatment for index patients | Outcomes |
| --- | --- | --- | --- | --- | --- | --- | --- | --- | --- | --- | --- |
| Zanamivir vs Placebo (9 RCTs) | | | | | | | | | | | |
| Ambrozai 2005 | Peer-reviewed publication  NR | Individual RCT | 489 | Lithuania, Netherlands, Israel | 67  20-107 | 52.6 | Pre-exposure | 9.2 | Zanamivir 10mg inhalations vs placebo once daily *14 days | - | Lab-confirmed symptomatic influenza, lab-confirmed influenza, lab-confirmed asymptomatic infection, mortality, adverse events related to drugs, serious adverse events |
| Hayden 2000 | Peer-reviewed publication  NAI30010 | Cluster RCT | 837 | USA | 26.2  NR | 44.9 | Post-exposure | 16.1 | Zanamivir 10mg inhalations vs placebo once daily *10 days | Yes | Lab-confirmed symptomatic influenza, lab-confirmed influenza, lab-confirmed asymptomatic infection, adverse events related to drugs, serious adverse events |
| Kaiser 2000 | Peer-reviewed publication  NAIA/B2009 | Individual RCT | 575 | Europe, North America | 34.3  13-77 | 40.4 | Post-exposure | 0 | Zanamivir 3.2mg intranasal vs 10mg inhalation vs 3.2mg intranasal and 10mg inhalation vs placebo once daily * 5 days | Unclear | Lab-confirmed symptomatic influenza, lab-confirmed influenza, lab-confirmed asymptomatic infection, adverse events related to drugs |
| LaForce 2007 | Peer-reviewed publication  NAI30034 | Individual RCT | 3363 | Canada, the Czech Republic, France, Germany, Latvia, USA | 60.4  12-94 | 42.1 | Pre-exposure | 67.2 | Zanamivir 10 mg oral inhaled vs placebo once daily*28 days | - | Lab-confirmed symptomatic influenza, lab-confirmed influenza, lab-confirmed asymptomatic infection, mortality, adverse events related to drugs, serious adverse outcomes |
| Monto 1999 | Peer-reviewed publication  NAIA3005 | Individual RCT | 1107 | USA | 28.8  18-69 | 40.6 | Pre-exposure | 14.4 | Zanamivir inhalation 10mg vs placebo once daily *28 days | - | Lab-confirmed symptomatic influenza, lab-confirmed influenza, lab-confirmed asymptomatic infection, adverse events related to drugs, serious adverse outcomes |
| Monto 2002 | Peer-reviewed publication  NAI30031 | Cluster RCT | 1291 | North America, Europe, Australia, New Zealand, and South Africa | 27.3  >=5 | 45.9 | Post-exposure | 10.2 | Zanamivir inhalation 10 mg vs placebo once daily * 10 days | No | Lab-confirmed symptomatic influenza, lab-confirmed influenza, lab-confirmed asymptomatic infection |
| 167-101 | Trial registration | Individual RCT | 317 | Japan | 33.6  >=18 | 26.5 | Pre-exposure | 0.3 | Zanamivir inhalation 10 mg vs placebo once daily * 28 days | - | Lab-confirmed symptomatic influenza, lab-confirmed influenza, lab-confirmed asymptomatic infection, mortality, adverse events related to drugs, serious adverse outcomes |
| NAIA2006 | Trial registration | Individual RCT | 64 | Canada, USA | 28.7  >=13 | 56.7 | Post-exposure | 0 | Zanamivir inhaled 5mg bid vs intranasal 6.4mg bid vs 10mg bid inhaled and 6.4mg bid intranasal placebo * 21 days | No | Lab-confirmed symptomatic influenza, lab-confirmed influenza, lab-confirmed asymptomatic infection, mortality, adverse events related to drugs, serious adverse outcomes |
| NAIB2006 | Trial registration | Individual RCT | 62 | France, Sweden and the United Kingdom | 35.45  >=18 | 54.9 | Post-exposure | 0 | Zanamivir inhalation 10mg bid vs placebo * 5 days | No | Lab-confirmed symptomatic influenza, lab-confirmed influenza, lab-confirmed asymptomatic infection, mortality, adverse events related to drugs, serious adverse outcomes |
| Oseltamivir vs placebo (6 RCTs) | | | | | | | | | | | |
| Kashiwagi 2000 | Peer-reviewed publication  NR | Individual RCT | 308 | Japan | 34.1  18-83 | 53.9 | Pre-exposure | NR | Oseltamivir 75 mg orals vs placebo once daily * 42 days | - | Lab-confirmed symptomatic influenza, lab-confirmed influenza, lab-confirmed asymptomatic infection, serious adverse events |
| Welliver 2001 | Peer-reviewed publication  Wv15799 | Cluster RCT | 957 | North America and Europe | 33.4  12-85 | 49.0 | Post-exposure | 12.6 | Oseltamivir 75mg vs placebo once daily * 7 days | No | lab-confirmed symptomatic influenza, lab-confirmed influenza, lab-confirmed asymptomatic infection, mortality, admission to hospital, adverse events related to drugs |
| Marianne 2014 | Peer-reviewed publication  NL27938.041.09 | Cluster RCT | 99 | Netherlands | 80.8  NR | 34.3 | Post-exposure | 87.9 | Oseltamivir 75 mg vs placebo once daily* 10 days | Yes | Lab-confirmed influenza, mortality |
| Hayden 1999 | Peer-reviewed publication  Wv15673 and Wv15697 | Individual RCT | 1559 | USA | 34.4  18-65 | 37.5 | Pre-exposure | 0 | Oseltamivir 75 mg once daily vs 75 mg twice daily vs placebo * 42 days | - | Lab-confirmed symptomatic influenza, lab-confirmed influenza, lab-confirmed asymptomatic infection, mortality, admission to hospital, adverse events related to drugs |
| WV15708 | Trial registration | Individual RCT | 385 | Australia, New Zealand, Saudi Arabia, Brazil | 76.3  >= 65 | 41.1 | Pre-exposure | 80.3 | Oseltamivir 75 mg vs placebo once daily * 42 days | - | Lab-confirmed symptomatic influenza, lab-confirmed influenza, lab-confirmed asymptomatic infection, mortality, admission to hospital, serious adverse events |
| WV15825 | Trial registration | Individual RCT | 548 | USA, France, Netherlands, Belgium, United Kingdom | 85.2  >= 65 | 31.0 | Pre-exposure | 97.3 | Oseltamivir 75 mg vs placebo once daily * 42 days | - | Lab-confirmed symptomatic influenza, lab-confirmed influenza, lab-confirmed asymptomatic infection, mortality, admission to hospital, serious adverse events |
| Laninamivir vs placebo (3 RCTs) | | | | | | | | | | | |
| Kashiwagi 2013 | Peer-reviewed publication  JapicCTI-111647 | Individual RCT | 1711 | Japan | 34.1  >=10 | 12.3 | Post-exposure | NR | Laninamivir 20mg once daily * 2 days vs 20mg once daily * 3 days vs placebo | Yes | Lab-confirmed symptomatic influenza, lab-confirmed influenza, lab-confirmed asymptomatic infection, adverse events related to antivirals |
| Kashiwagi 2016 | Peer-reviewed publication  JapicCTI-142679 | Individual RCT | 803 | Japan | 35.3  >=10 | 11.0 | Post-exposure | 37.8 | Laninamivir 40mg once vs 20mg once daily * 2 days vs placebo | Yes | Lab-confirmed symptomatic influenza, lab-confirmed influenza, lab-confirmed asymptomatic infection, adverse events related to antivirals |
| Nakano 2016 | Peer-reviewed publication  NR | Individual RCT | 343 | Japan | 6.8  <10 | 49.9 | Post-exposure | 41.1 | Laninamivir inhalation 20 mg once vs placebo | Yes | Lab-confirmed symptomatic influenza, lab-confirmed influenza, lab-confirmed asymptomatic infection, mortality, adverse events related to antivirals, serious adverse events |
| Baloxavir vs placebo (1 RCT) | | | | | | | | | | | |
| Ikematsu 2020 | Peer-reviewed publication  JapicCTI-184180 | Individual RCT | 752 | Japan | 33.6  NR | 21.6 | Post-exposure | 34.1 | Baloxavir oral (weight < 10 kg: 1mg/kg; 10 to < 20 kg, 10mg; 20 to < 40 kg, 20mg; ≥ 40 kg 40mg) vs placebo once | Yes | Lab-confirmed symptomatic influenza, lab-confirmed influenza, lab-confirmed asymptomatic infection, mortality, adverse events related to antivirals, serious adverse outcomes |
| Rimantadine vs Placebo (6 RCTs) | | | | | | | | | | | |
| Brady 1990 | Peer-reviewed publication  NR | Individual RCT | 228 | USA | 31.4  18-55 | 25.4 | Pre-exposure | 0 | Rimantadine 100 mg vs placebo orals once daily *14 days | - | Lab-confirmed influenza |
| Bricaire 1990 | Peer-reviewed publication  NR | Cluster RCT | 301 | France | NR  NR | 48.7 | Post-exposure | NR | Rimantadine (adults 200 mg/d, older 100 mg/d, child 5mg/kg/d) vs placebo orals daily for 10 days | Unclear | Lab-confirmed influenza |
| Carwford 1988 | Peer-reviewed publication  NR | Individual RCT | 107 | USA | NR  NR | NR | Pre-exposure | 0 | Rimantadine 5 mg/kg/d (maximum dose, 150 mg/d for children under 10 years of age and 200 mg/d for children more than 10 years) once vs placebo | - | Lab-confirmed influenza |
| Reuman 1988 | Peer-reviewed publication  NR | Individual RCT | 476 | USA | NR  18-55 | NR | Pre-exposure | 0 | Rimantadine 100mg/d vs 200mg/d vs placebo *42 days | - | Lab-confirmed influenza |
| Hayden 1989 | Peer-reviewed publication  NR | Cluster RCT | 115 | USA | 25.5  1-57 | 48.7 | Post-exposure | 0 | Rimantadine (the daily dose was 200 mg for adults and for children over nine or those who weighed more than 30 kg. The dose was 5 mg per kilogram of body weight up to a maximum of 150 mg per day for children less than 9 years of age or for those weighing less than 30kg) vs placebo once daily for 10 days. | Yes | Lab-confirmed influenza |
| Monto 1995 | Peer-reviewed publication  NR | Individual RCT | 328 | USA | 86.3  NR | 16.2 | Pre-exposure | 93.3 | Rimantadine 200 mg/day vs 100 mg/day vs placebo for 56 days | - | Lab-confirmed influenza, mortality |
| Amantadine vs placebo (4 RCTs) | | | | | | | | | | | |
| Oker-Blom 1970 | Peer-reviewed publication  NR | Individual RCT | 391 | Finland | 22  NR | 66.2 | Pre-exposure | NR | Amantadine 100 mg vs placebo bid *30 days | - | Lab-confirmed influenza |
| Payler 1984 | Peer-reviewed publication  NR | Individual RCT | 536 | Britain | NR  18 – 24 | NR | Pre-exposure | 100 | Amantadine 100 mg vs placebo once daily *14 days | - | Lab-confirmed influenza |
| Pettersson 1980 | Peer-reviewed publication  NR | Individual RCT | 192 | Finland | 50.6  >=10 | 71.2 | Pre-exposure | NR | Amantadine 100 mg bid * 21 days vs 100mg bid *35 days vs placebo | - | Lab-confirmed influenza, mortality |
| Nafta 1970 | Peer-reviewed publication  NR | Individual RCT | 215 | Romania | NR  3-50 | NR | Pre-exposure | NR | Amantadine 100 mg vs placebo bid * 20 days | - | Lab-confirmed influenza, mortality, serious adverse events |
| Zanamivir vs Rimantadine; Zanamivir vs standard care (1 RCT) | | | | | | | | | | | |
| Schilling 1988 | Peer-reviewed publication  NR | Cluster RCT | 140 | USA | NR  NR | NR | Pre-exposure | NR | 1. Zanamivir 10 mg inhaled and 4.4 mg intransally bid vs rimantadine 100 mg *14 days; 2. Zanamivir 10 mg inhaled and 4.4 mg intransally bid vs standard of care * 14 days | - | Lab-confirmed influenza |
| Rimantadine vs Amantadine vs placebo (2 RCT) | | | | | | | | | | | |
| Dolin 1982 | Peer-reviewed publication  NR | Individual RCT | 378 | USA | 25.6  18-45 | NR | Pre-exposure | 0 | Rimantadine 100mg bid vs amantadine 100mg bid vs placebo * 42 days | - | Lab-confirmed influenza |
| Quarles 1981 | Peer-reviewed publication  NR | Individual RCT | 308 | USA | NR  18–24 | NR | Pre-exposure | NR | Rimantadine 100mg bid vs amantadine 100mg bid vs placebo * 42 days | - | Lab-confirmed influenza |
| Rimantadine vs Zanamivir (1 RCT) | | | | | | | | | | | |
| Gravenstein 2005 | Peer-reviewed publication  NAIA3003 | Individual RCT | 469 | USA | 76.0  44-102 | 29.3 | Pre-exposure | 97.3 | Rimantadine 100mg once daily vs zanamivir oral inhalations 10mg once daily *14 days | - | Lab-confirmed symptomatic influenza, mortality, adverse events related to antivirals, serious adverse outcomes |

RCT: randomized controlled trial; NR = not report

Note: Of the included trials, 33 reported lab-confirmed influenza outcomes, among which 18 reported lab-confirmed symptomatic influenza and lab-confirmed asymptomatic influenza virus infection separately; one trial only reported lab-confirmed symptomatic influenza; 14 trials only reported lab-confirmed influenza without specifying symptom status. Seventeen trials reported all-cause mortality; 4 reported admission to hospital; 13 reported adverse events related to drugs; and 16 reported serious adverse events. No studies reported duration of symptoms, length of hospitalization, progression to invasive mechanical ventilation, length of mechanical ventilation, progression of disease severity, admission to ICU, or emergence of antiviral resistance.

### Appendix 7 Risk of bias for eligible studies

2a for individual randomized trials

| Study | Trial registration | Outcome | Sequence generation | Allocation concealment | Blinding | | | | | Lost to follow-up | Other bias |
| --- | --- | --- | --- | --- | --- | --- | --- | --- | --- | --- | --- |
|  |  |  |  |  | Patients | Health care providers | Data collectors | Outcome assessors and/or adjudicators | Data analysts |  |  |
| Ambrozai 2005 | NAIA3004 | Lab-confirmed symptomatic influenza | Low | Low | Low | Low | Low | Low | Low | Low | Low |
|  |  | Lab-confirmed influenza | Low | Low | Low | Low | Low | Low | Low | Low | Low |
|  |  | Lab-confirmed asymptomatic influenza | Low | Low | Low | Low | Low | Low | Low | Low | Low |
|  |  | Mortality | Low | Low | Low | Low | Low | Low | Low | Low | Low |
|  |  | Adverse events | Low | Low | Low | Low | Low | Low | High | Low | Low |
|  |  | Serious adverse events | Low | Low | Low | Low | Low | Low | High | Low | Low |
|  |  | Adverse events related to drugs | Low | Low | Low | Low | Low | Low | High | Low | Low |
| Brady 1990 | Not applicable | Lab-confirmed influenza | Low | Low | Low | Low | Low | Low | Low | Low | Low |
|  |  | Adverse events | Low | Low | Low | Low | High | High | High | Low | Low |
| Carwford 1988 | Not applicable | Lab-confirmed influenza | High | High | Low | Low | Low | Low | Low | Low | Low |
|  |  | Adverse events | High | High | Low | Low | High | High | High | Low | Low |
| Dolin 1982 | Not applicable | Lab-confirmed influenza | Low | Low | Low | Low | Low | Low | Low | Low | Low |
| Gravenstein 2005 | NAIA3003 | Lab-confirmed symptomatic influenza | Low | Low | Low | Low | Low | Low | Low | Low | Low |
|  |  | Mortality | Low | Low | Low | Low | Low | Low | Low | Low | Low |
|  |  | Adverse events | Low | Low | Low | Low | Low | Low | Low | Low | Low |
|  |  | Serious adverse events | Low | Low | Low | Low | Low | Low | Low | Low | Low |
|  |  | Adverse events related to drugs | Low | Low | Low | Low | Low | Low | Low | Low | Low |
| Hayden 1999 | Wv15673 Wv15697 | Lab-confirmed symptomatic influenza | Low | Low | Low | Low | Low | Low | Low | Low | Low |
|  |  | Lab-confirmed influenza | Low | Low | Low | Low | Low | Low | Low | Low | Low |
|  |  | Lab-confirmed asymptomatic influenza | Low | Low | Low | Low | Low | Low | Low | Low | Low |
|  |  | Mortality | Low | Low | Low | Low | Low | Low | Low | Low | Low |
|  |  | Admission to hospital | Low | Low | Low | Low | Low | Low | Low | Low | Low |
|  |  | Adverse events | Low | Low | Low | Low | High | High | High | Low | Low |
|  |  | Serious adverse events | Low | Low | Low | Low | High | High | High | Low | Low |
| Ikematsu 2020 | JapicCTI-184180 | Lab-confirmed symptomatic influenza | Low | Low | Low | Low | Low | Low | Low | Low | Low |
|  |  | Lab-confirmed influenza | Low | Low | Low | Low | Low | Low | Low | Low | Low |
|  |  | Lab-confirmed asymptomatic influenza | Low | Low | Low | Low | Low | Low | Low | Low | Low |
|  |  | Mortality | Low | Low | Low | Low | Low | Low | Low | Low | Low |
|  |  | Adverse events | Low | Low | Low | Low | High | High | High | Low | Low |
|  |  | Serious adverse events | Low | Low | Low | Low | High | High | High | Low | Low |
|  |  | Adverse events related to drugs | Low | Low | Low | Low | High | High | High | Low | Low |
| Kaiser 2000 | NAIA/B2009 | Lab-confirmed symptomatic influenza | High | Low | Low | Low | Low | Low | Low | Low | Low |
|  |  | Lab-confirmed influenza | High | Low | Low | Low | Low | Low | Low | Low | Low |
|  |  | Lab-confirmed asymptomatic influenza | High | Low | Low | Low | Low | Low | Low | Low | Low |
|  |  | Adverse events | High | Low | Low | Low | Low | Low | Low | Low | Low |
|  |  | Adverse events related to drugs | High | Low | Low | Low | Low | Low | Low | Low | Low |
| Kashiwagi 2000 | Not applicable | Lab-confirmed symptomatic influenza | Low | Low | Low | Low | Low | Low | Low | Low | Low |
|  |  | Lab-confirmed influenza | Low | Low | Low | Low | Low | Low | Low | Low | Low |
|  |  | Lab-confirmed asymptomatic influenza | Low | Low | Low | Low | Low | Low | Low | Low | Low |
|  |  | Adverse events | Low | Low | Low | Low | High | High | High | Low | Low |
|  |  | Serious adverse events | Low | Low | Low | Low | High | High | High | Low | Low |
| Kashiwagi 2013 | JapicCTI-111647 | Lab-confirmed symptomatic influenza | Low | Low | Low | Low | Low | Low | Low | Low | Low |
|  |  | Lab-confirmed influenza | Low | Low | Low | Low | Low | Low | Low | Low | Low |
|  |  | Lab-confirmed asymptomatic influenza | Low | Low | Low | Low | Low | Low | Low | Low | Low |
|  |  | Adverse events | Low | Low | Low | Low | Low | Low | Low | Low | Low |
|  |  | Serious adverse events | Low | Low | Low | Low | Low | Low | Low | Low | Low |
| Kashiwagi 2016 | JapicCTI-142679 | Lab-confirmed symptomatic influenza | Low | Low | Low | Low | Low | Low | Low | Low | Low |
|  |  | Lab-confirmed influenza | Low | Low | Low | Low | Low | Low | Low | Low | Low |
|  |  | Lab-confirmed asymptomatic influenza | Low | Low | Low | Low | Low | Low | Low | Low | Low |
|  |  | Adverse events | Low | Low | Low | Low | Low | Low | Low | Low | Low |
|  |  | Serious adverse events | Low | Low | Low | Low | Low | Low | Low | Low | Low |
| LaForce 2007 | NAI30034 | Lab-confirmed symptomatic influenza | High | Low | Low | Low | Low | Low | Low | Low | Low |
|  |  | Lab-confirmed influenza | High | Low | Low | Low | Low | Low | Low | Low | Low |
|  |  | Lab-confirmed asymptomatic influenza | High | Low | Low | Low | Low | Low | Low | Low | Low |
|  |  | Mortality | High | Low | Low | Low | Low | Low | Low | Low | Low |
|  |  | Adverse events | High | Low | Low | Low | Low | Low | High | Low | Low |
|  |  | Serious adverse events | High | Low | Low | Low | Low | Low | High | Low | Low |
|  |  | Adverse events related to drugs | High | Low | Low | Low | Low | Low | High | Low | Low |
| Monto 1995 | Not applicable | Lab-confirmed influenza | High | High | Low | Low | Low | Low | Low | Low | Low |
|  |  | Mortality | High | High | Low | Low | Low | Low | Low | Low | Low |
| Monto 1999 | NAIA3005 | Lab-confirmed influenza | Low | Low | Low | Low | Low | Low | Low | Low | Low |
|  |  | Lab-confirmed influenza without symptoms | Low | Low | Low | Low | Low | Low | Low | Low | Low |
|  |  | Lab-confirmed influenza with symptoms | Low | Low | Low | Low | Low | Low | Low | Low | Low |
|  |  | Adverse events | Low | Low | Low | Low | High | High | High | Low | Low |
|  |  | Serious adverse events | Low | Low | Low | Low | High | High | High | Low | Low |
|  |  | Adverse events related to drugs | Low | Low | Low | Low | High | High | High | Low | Low |
| Nafta 1970 | Not applicable | Lab-confirmed influenza | High | High | Low | Low | Low | Low | Low | Low | Low |
|  |  | Serious adverse events | High | High | Low | Low | High | High | High | Low | Low |
| Nakano 2016 | Not applicable | Lab-confirmed symptomatic influenza | Low | Low | Low | Low | Low | Low | Low | Low | Low |
|  |  | Lab-confirmed influenza | Low | Low | Low | Low | Low | Low | Low | Low | Low |
|  |  | Lab-confirmed asymptomatic influenza | Low | Low | Low | Low | Low | Low | Low | Low | Low |
|  |  | Mortality | Low | Low | Low | Low | Low | Low | Low | Low | Low |
|  |  | Adverse events | Low | Low | Low | Low | High | High | High | Low | Low |
|  |  | Serious adverse events | Low | Low | Low | Low | High | High | High | Low | Low |
|  |  | Adverse events related to drugs | Low | Low | Low | Low | High | High | High | Low | Low |
| Oker-Blom 1970 | Not applicable | Lab-confirmed influenza | High | Low | Low | Low | Low | Low | Low | Low | Low |
| Payler 1984 | Not applicable | Lab-confirmed influenza | High | High | Low | Low | Low | Low | Low | Low | Low |
| Pettersson 1980 | Not applicable | Lab-confirmed influenza | Low | Low | Low | Low | Low | Low | Low | Low | Low |
|  |  | Mortality | Low | Low | Low | Low | Low | Low | Low | Low | Low |
|  |  | Adverse events | Low | Low | Low | Low | High | High | High | Low | Low |
| Quarles 1981 | Not applicable | Lab-confirmed influenza | High | High | Low | Low | Low | Low | Low | Low | High |
| Reuman 1988 | Not applicable | Lab-confirmed influenza | Low | Low | Low | Low | Low | Low | Low | Low | Low |
|  |  | Adverse events | Low | Low | Low | Low | High | High | High | Low | Low |
| 167-101 | 167-101 | Lab-confirmed symptomatic influenza | Low | Low | Low | Low | Low | Low | Low | Low | Low |
|  |  | Lab-confirmed influenza | Low | Low | Low | Low | Low | Low | Low | Low | Low |
|  |  | Lab-confirmed asymptomatic influenza | Low | Low | Low | Low | Low | Low | Low | Low | Low |
|  |  | Mortality | Low | Low | Low | Low | Low | Low | Low | Low | Low |
|  |  | Adverse events | Low | Low | Low | Low | Low | Low | Low | Low | Low |
|  |  | Serious adverse events | Low | Low | Low | Low | Low | Low | Low | Low | Low |
|  |  | Adverse events related to drugs | Low | Low | Low | Low | Low | Low | Low | Low | Low |
| NAIA2006 | NAIA2006 | Lab-confirmed symptomatic influenza | Low | Low | Low | Low | Low | Low | Low | High | Low |
|  |  | Lab-confirmed influenza | Low | Low | Low | Low | Low | Low | Low | High | Low |
|  |  | Lab-confirmed asymptomatic influenza | Low | Low | Low | Low | Low | Low | Low | High | Low |
|  |  | Mortality | Low | Low | Low | Low | Low | Low | Low | High | Low |
|  |  | Adverse events | Low | Low | Low | Low | Low | Low | Low | High | Low |
|  |  | Serious adverse events | Low | Low | Low | Low | Low | Low | Low | High | Low |
|  |  | Adverse events related to drugs | Low | Low | Low | Low | Low | Low | Low | High | Low |
| NAIB2006 | NAIB2006 | Lab-confirmed symptomatic influenza | Low | Low | Low | Low | Low | Low | Low | Low | Low |
|  |  | Lab-confirmed influenza | Low | Low | Low | Low | Low | Low | Low | Low | Low |
|  |  | Lab-confirmed asymptomatic influenza | Low | Low | Low | Low | Low | Low | Low | Low | Low |
|  |  | Mortality | Low | Low | Low | Low | Low | Low | Low | Low | Low |
|  |  | Adverse events | Low | Low | Low | Low | Low | Low | Low | Low | Low |
|  |  | Serious adverse events | Low | Low | Low | Low | Low | Low | Low | Low | Low |
|  |  | Adverse events related to drugs | Low | Low | Low | Low | Low | Low | Low | Low | Low |
| WV15708 | WV15708 | Lab-confirmed symptomatic influenza | Low | Low | Low | Low | Low | Low | Low | Low | Low |
|  |  | Lab-confirmed influenza | Low | Low | Low | Low | Low | Low | Low | Low | Low |
|  |  | Lab-confirmed asymptomatic influenza | Low | Low | Low | Low | Low | Low | Low | Low | Low |
|  |  | Mortality | Low | Low | Low | Low | Low | Low | Low | Low | Low |
|  |  | Admission to hospital | Low | Low | Low | Low | Low | Low | Low | Low | Low |
|  |  | Adverse events | Low | Low | Low | Low | Low | Low | Low | Low | Low |
|  |  | Serious adverse events | Low | Low | Low | Low | Low | Low | Low | Low | Low |
|  |  | Adverse events related to drugs | Low | Low | Low | Low | Low | Low | Low | Low | Low |
| WV15825 | WV15825 | Lab-confirmed symptomatic influenza | Low | Low | Low | Low | Low | Low | Low | Low | Low |
|  |  | Lab-confirmed influenza | Low | Low | Low | Low | Low | Low | Low | Low | Low |
|  |  | Lab-confirmed asymptomatic influenza | Low | Low | Low | Low | Low | Low | Low | Low | Low |
|  |  | Mortality | Low | Low | Low | Low | Low | Low | Low | Low | Low |
|  |  | Admission to hospital | Low | Low | Low | Low | Low | Low | Low | Low | Low |
|  |  | Adverse events | Low | Low | Low | Low | Low | Low | Low | Low | Low |
|  |  | Serious adverse events | Low | Low | Low | Low | Low | Low | Low | Low | Low |

2b Risk of bias for cluster randomized trials.

| Study | Trial registration | Outcome | Bias arising from the randomization process | Bias arising from the timing of identification and recruitment of participants | Bias due to deviations from intended interventions | Bias due to missing outcome data | Bias in measurement of the outcome | Bias in selection of the reported result |
| --- | --- | --- | --- | --- | --- | --- | --- | --- |
| Bricaire 1990 | Not applicable | Lab-confirmed influenza | High | High | Low | Low | Low | Low |
|  |  | Adverse events | High | High | Low | Low | Low | Low |
| Hayden 1989 | Not applicable | Lab-confirmed influenza | High | High | Low | Low | Low | Low |
| Hayden 2000 | NAI30010 | Lab-confirmed symptomatic influenza | High | Low | Low | Low | Low | Low |
|  |  | Lab-confirmed influenza | High | Low | Low | Low | Low | Low |
|  |  | Lab-confirmed asymptomatic influenza | High | Low | Low | Low | Low | Low |
|  |  | Adverse events | High | Low | Low | Low | Low | Low |
|  |  | Serious adverse events | High | Low | Low | Low | Low | Low |
|  |  | Adverse events related to drugs | High | Low | Low | Low | Low | Low |
| Marianne 2014 | NL27938.041.09 | Lab-confirmed influenza | Low | Low | Low | Low | Low | Low |
|  |  | Mortality | Low | Low | Low | Low | Low | Low |
|  |  | Adverse events | Low | Low | Low | Low | Low | Low |
| Monto 2002 | NAI30031 | Lab-confirmed symptomatic influenza | High | High | Low | Low | Low | Low |
|  |  | Lab-confirmed influenza | High | High | Low | Low | Low | Low |
|  |  | Lab-confirmed asymptomatic influenza | High | High | Low | Low | Low | Low |
|  |  | Adverse events | High | High | Low | Low | Low | Low |
| Schilling 1988 | Not applicable | Lab-confirmed influenza | High | High | Low | Low | Low | Low |
| Welliver 2001 | Wv15799 | Lab-confirmed symptomatic influenza | Low | Low | Low | Low | Low | Low |
|  |  | Lab-confirmed influenza | Low | Low | Low | Low | Low | Low |
|  |  | Lab-confirmed asymptomatic influenza | Low | Low | Low | Low | Low | Low |
|  |  | Mortality | Low | Low | Low | Low | Low | Low |
|  |  | Admission to hospital | Low | Low | Low | Low | Low | Low |
|  |  | Adverse events | Low | Low | Low | Low | Low | Low |
|  |  | Serious adverse events | Low | Low | Low | Low | Low | Low |
|  |  | Adverse events related to drugs | Low | Low | Low | Low | Low | Low |

Note: Among the included trials, five were rated at high risk of bias for the objective outcomes (lab-confirmed influenza virus infection, lab-confirmed symptomatic influenza, lab-confirmed asymptomatic influenza virus infection, admission to hospital, all-cause mortality) due to sequence generation and allocation concealment limitations, and eleven were rated at high risk of bias for subjective outcomes (adverse events related to drugs, serious adverse events), mostly due to failure to blind data collectors, outcome assessors and/or adjudicators and data analysts. Among the cluster randomized trials, two were rated at high risk of bias for both objective outcomes and subjective outcomes [37, 56], and one was rated at high risk of bias for objective outcomes, all due to bias arising from the randomization process and bias arising from the timing of identification and recruitment of participants.

### Appendix 8 Network plots.

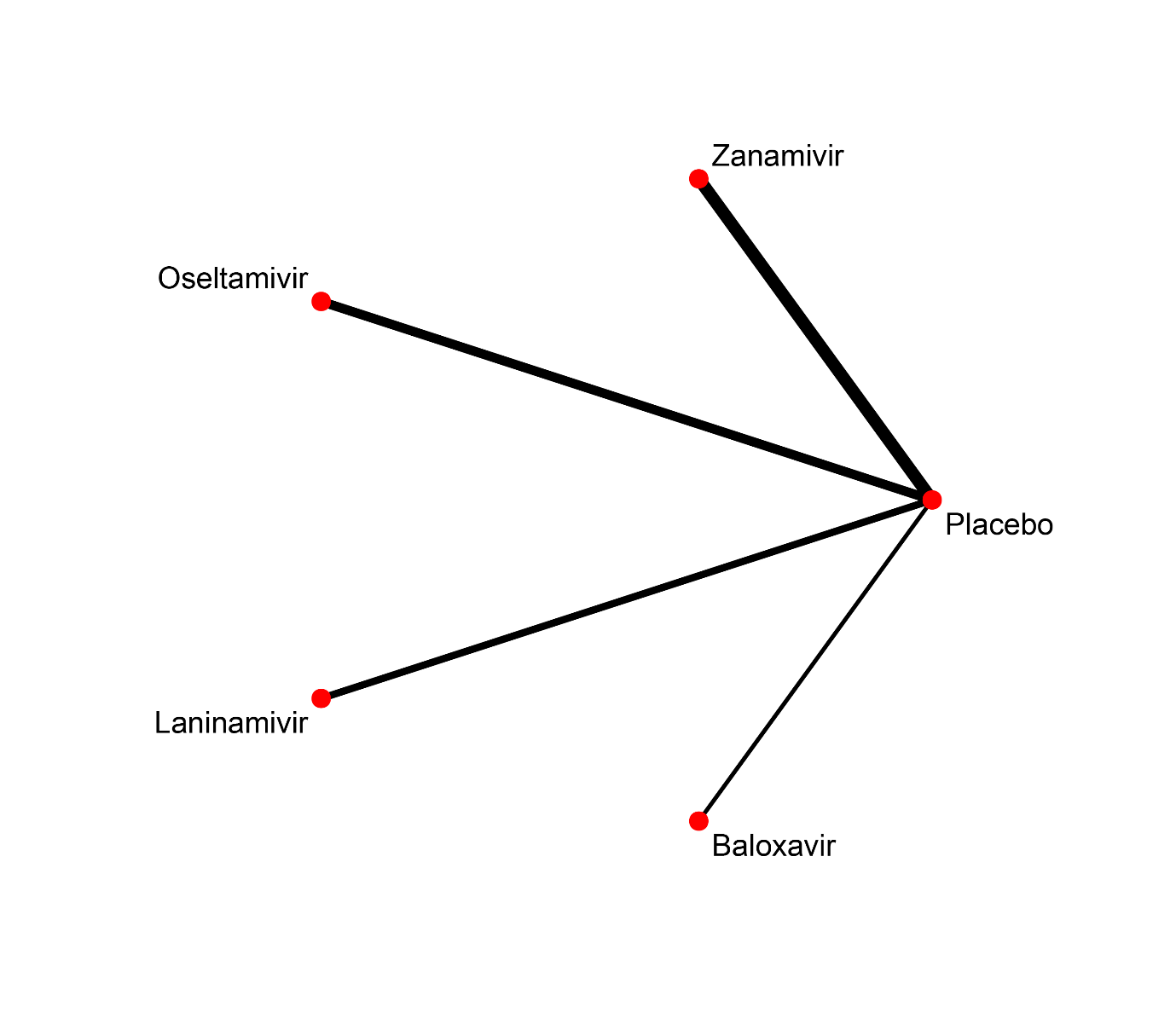

#### Figure 1 Network plot of trials evaluating antivirals prophylaxis against lab-confirmed asymptomatic influenza.

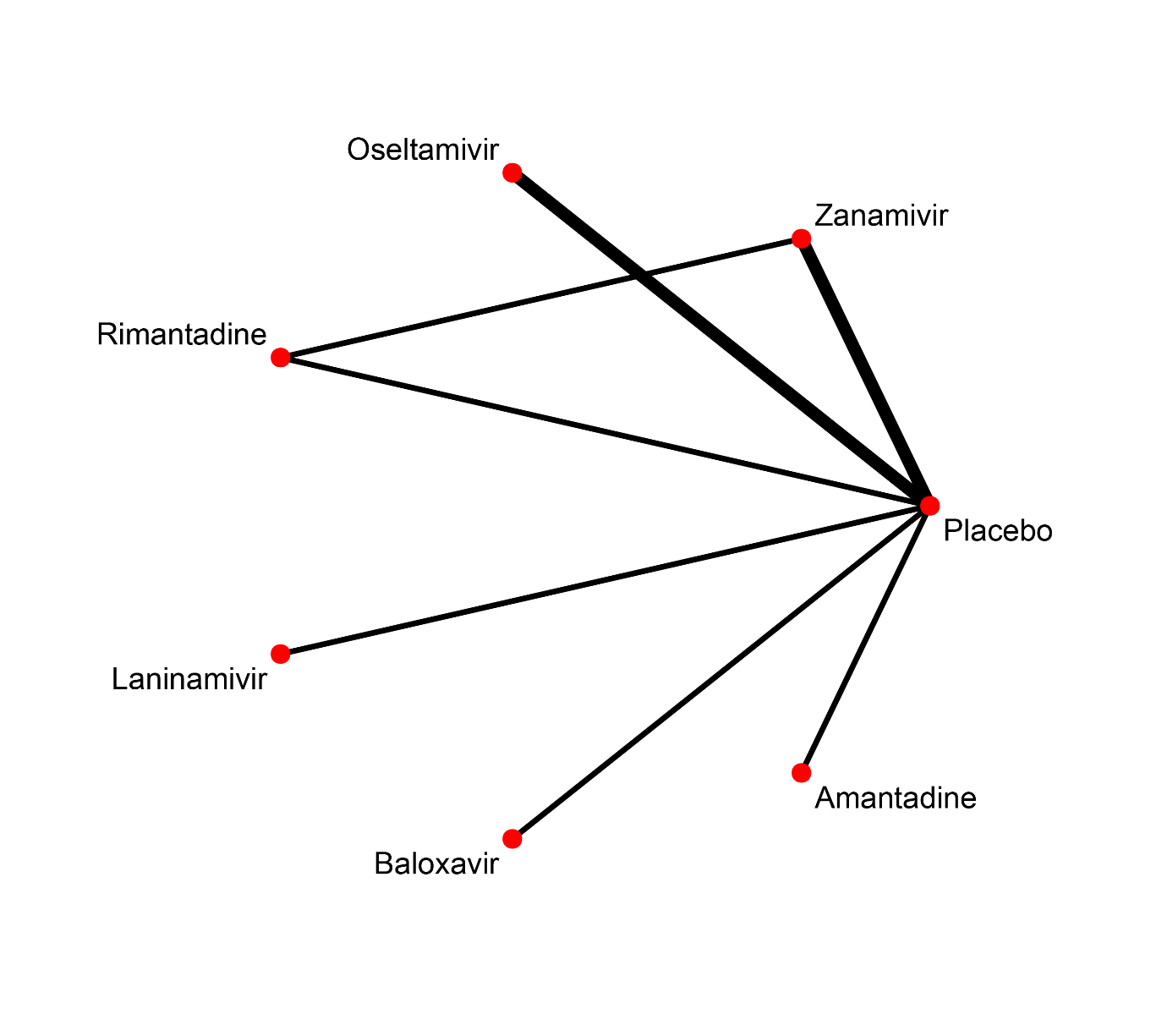

#### Figure 2 Network plot of trials evaluating antivirals prophylaxis against all-cause mortality.

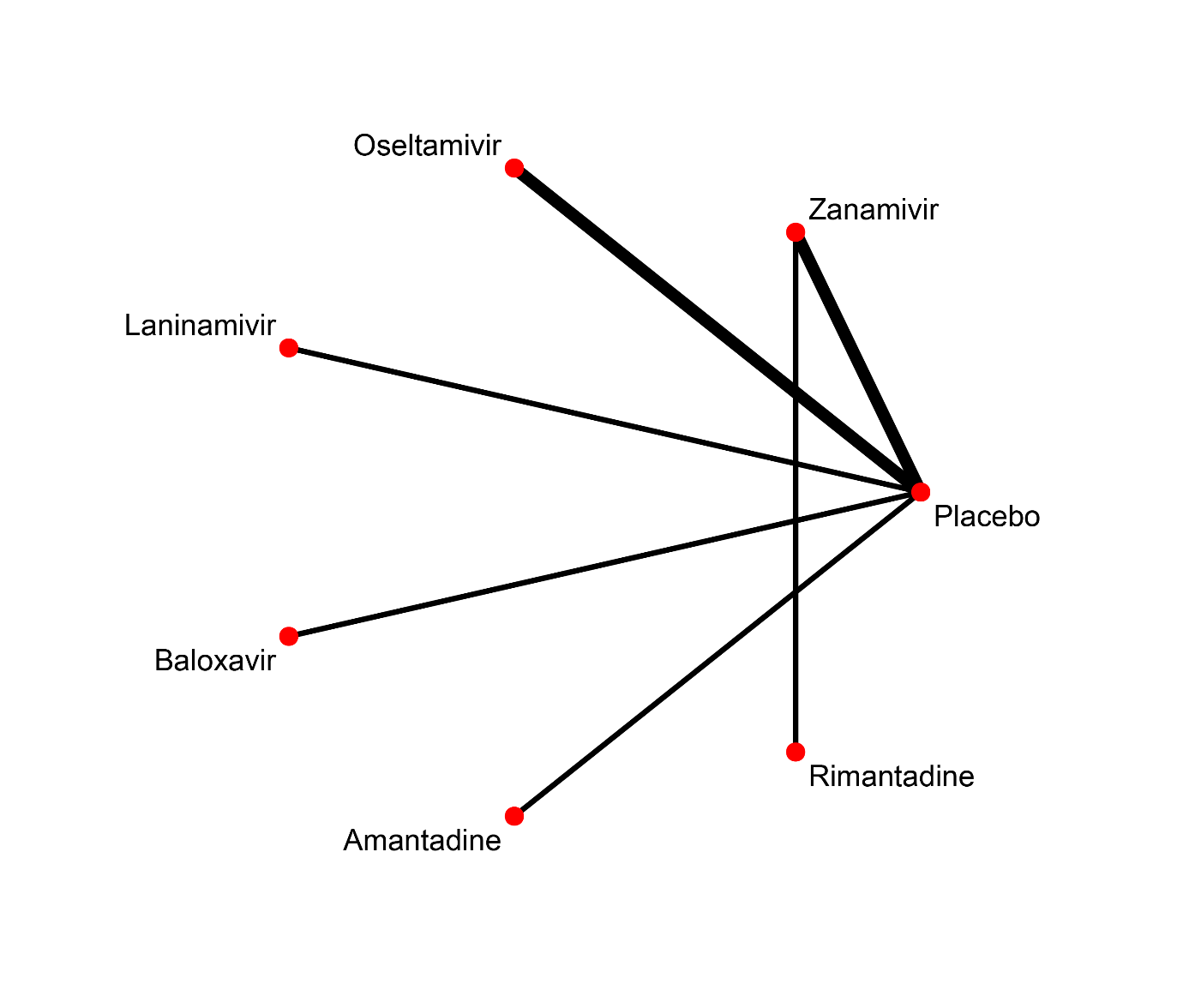

#### Figure 3 Network plot of trials evaluating antivirals prophylaxis against serious adverse events.

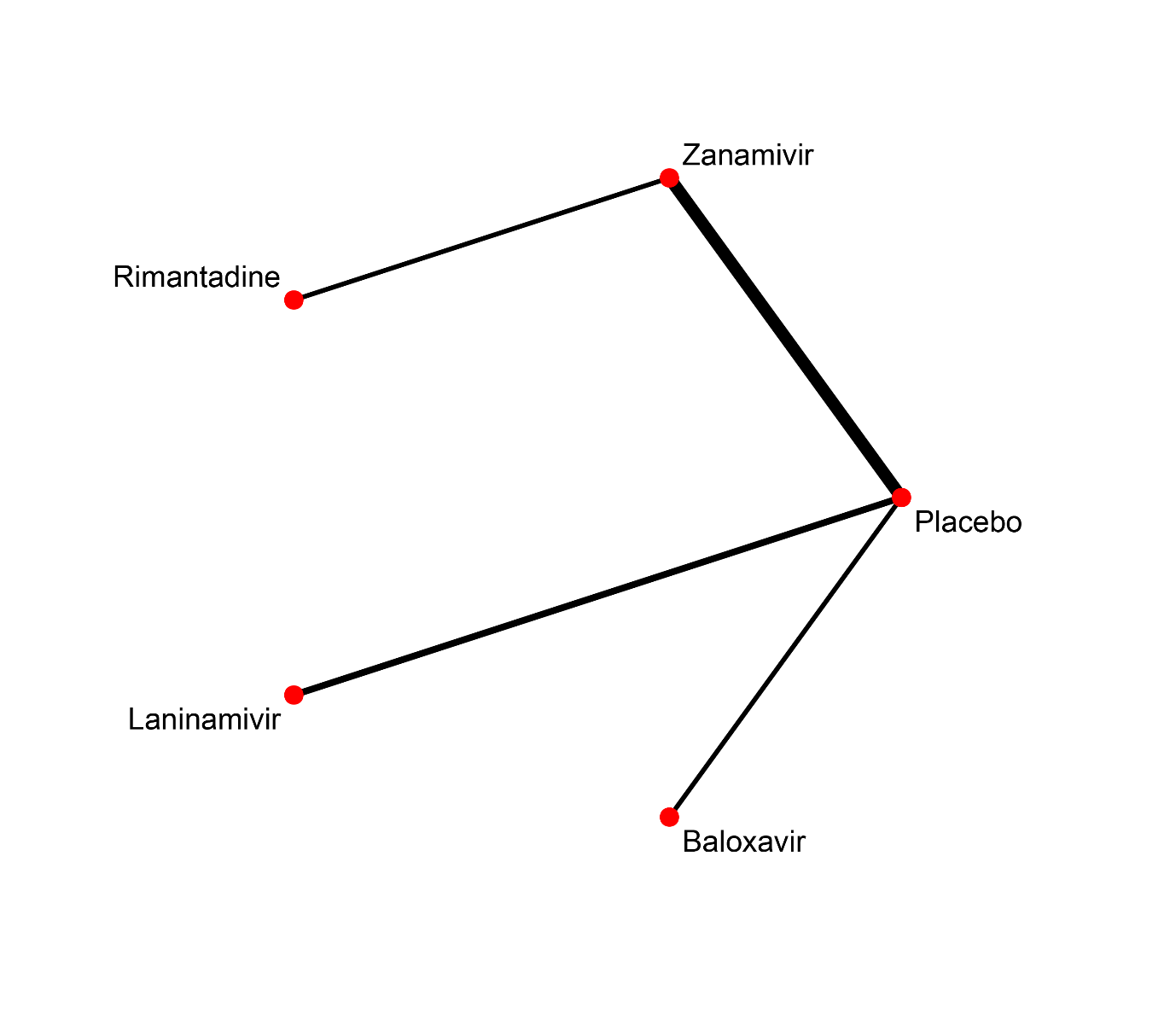

#### Figure 4 Network plot of trials evaluating antivirals prophylaxis against adverse events related to drugs.

### Appendix 9 Forest plots

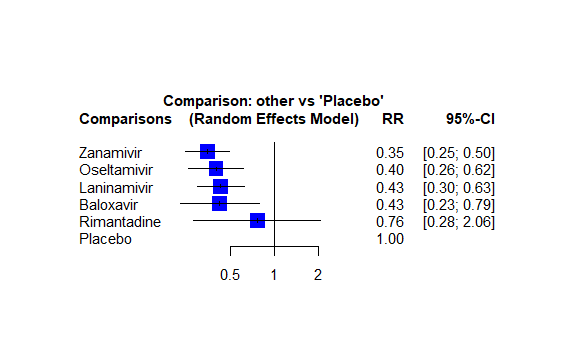

#### Figure 1 Forest plot for antivirals prophylaxis against lab-confirmed symptomatic influenza.

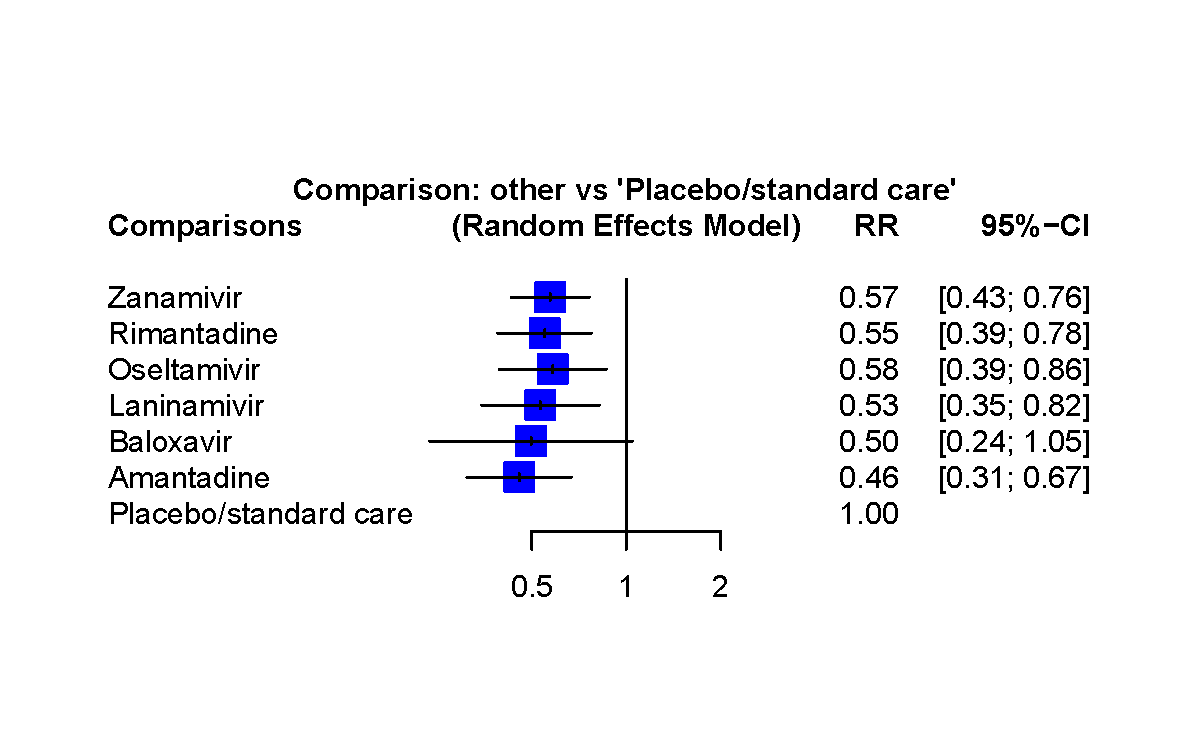

#### Figure 2 Forest plot for antivirals prophylaxis against lab-confirmed influenza.

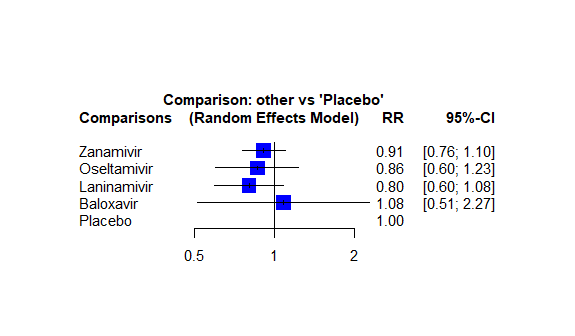

#### Figure 3 Forest plot for antivirals prophylaxis against lab-confirmed asymptomatic influenza.

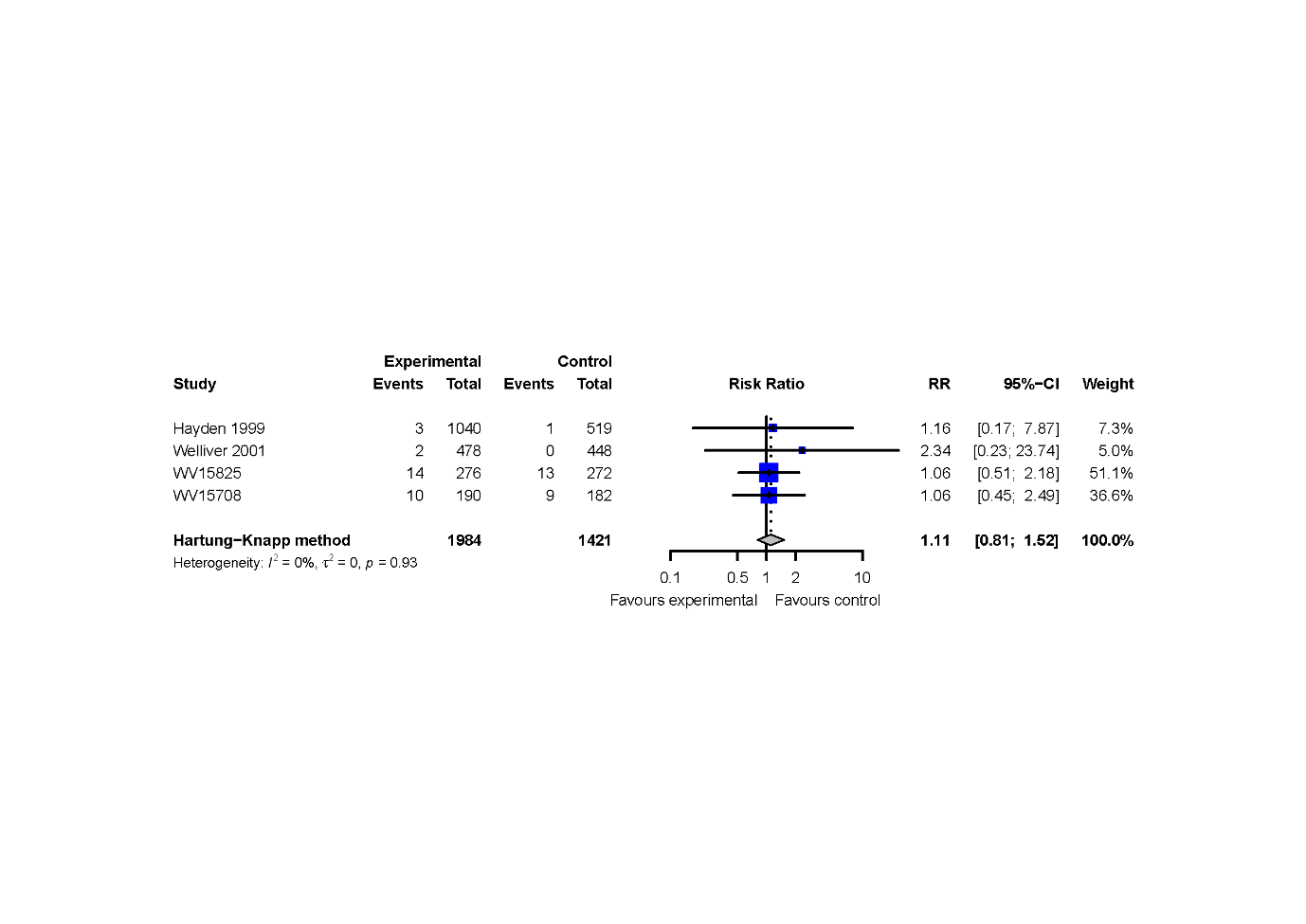

#### Figure 4 Forest plot for antivirals prophylaxis against admission to hospital.

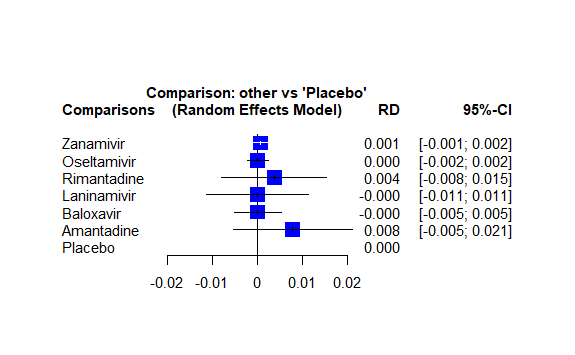

#### Figure 5 Forest plot for antivirals prophylaxis against all-cause mortality.

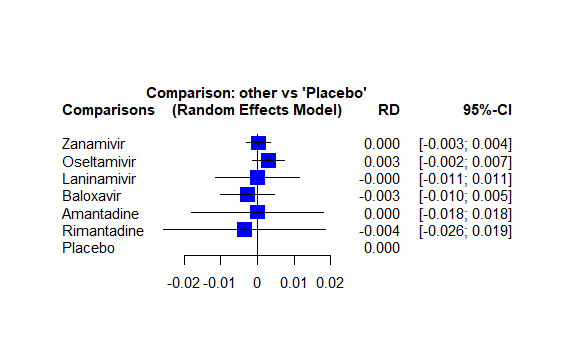

#### Figure 6 Forest plot for antivirals prophylaxis against serious adverse events.

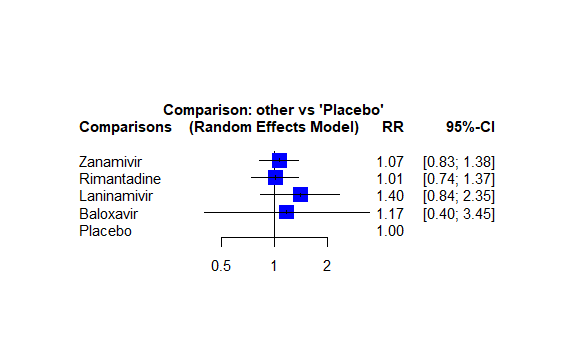

#### Figure 7 Forest plot for antivirals prophylaxis against adverse events related to drugs.

### Appendix 10 Direct, indirect, and network treatment estimates for each outcome.

#### Table 1 Direct, indirect, and network treatment estimates for lab-confirmed symptomatic influenza.

| **Comparison** | **k** | **Prop** | **NMA (95% CI)** | **Direct (95% CI)** | **Indirect (95% CI)** | **Diff (95% CI)** | **z** | **Incoherence p-value** |
| --- | --- | --- | --- | --- | --- | --- | --- | --- |
| Baloxavir vs. Laninamivir | 0 | 0 | 0.98 [0.48; 2.03] | NA | 0.98 [0.48; 2.03] | NA | NA | NA |
| Baloxavir vs. Oseltamivir | 0 | 0 | 1.06 [0.50; 2.26] | NA | 1.06 [0.50; 2.26] | NA | NA | NA |
| Baloxavir vs. Placebo | 1 | 1.00 | 0.43 [0.23; 0.79] | 0.43 [0.23; 0.79] | NA | NA | NA | NA |
| Baloxavir vs. Rimantadine | 0 | 0 | 0.56 [0.17; 1.80] | NA | 0.56 [0.17; 1.80] | NA | NA | NA |
| Baloxavir vs. Zanamivir | 0 | 0 | 1.21 [0.60; 2.47] | NA | 1.21 [0.60; 2.47] | NA | NA | NA |
| Laninamivir vs. Oseltamivir | 0 | 0 | 1.08 [0.61; 1.91] | NA | 1.08 [0.61; 1.91] | NA | NA | NA |
| Laninamivir vs. Placebo | 3 | 1.00 | 0.43 [0.30; 0.63] | 0.43 [0.30; 0.63] | NA | NA | NA | NA |
| Laninamivir vs. Rimantadine | 0 | 0 | 0.57 [0.20; 1.64] | NA | 0.57 [0.20; 1.64] | NA | NA | NA |
| Laninamivir vs. Zanamivir | 0 | 0 | 1.23 [0.74; 2.05] | NA | 1.23 [0.74; 2.05] | NA | NA | NA |
| Oseltamivir vs. Placebo | 5 | 1.00 | 0.40 [0.26; 0.62] | 0.40 [0.26; 0.62] | NA | NA | NA | NA |
| Oseltamivir vs. Rimantadine | 0 | 0 | 0.53 [0.18; 1.56] | NA | 0.53 [0.18; 1.56] | NA | NA | NA |
| Oseltamivir vs. Zanamivir | 0 | 0 | 1.14 [0.66; 1.99] | NA | 1.14 [0.66; 1.99] | NA | NA | NA |
| Rimantadine vs. Placebo | 0 | 0 | 0.76 [0.28; 2.06] | NA | 0.76 [0.28; 2.06] | NA | NA | NA |
| Zanamivir vs. Placebo | 9 | 1.00 | 0.35 [0.25; 0.50] | 0.35 [0.25; 0.50] | NA | NA | NA | NA |
| Rimantadine vs. Zanamivir | 1 | 1.00 | 2.17 [0.86; 5.53] | 2.17 [0.86; 5.53] | NA | NA | NA | NA |

Comparison: treatment comparison; k: number of studies providing direct evidence; prop: direct evidence proportion; NMA: estimated treatment effect (RR) in network meta-analysis; direct: estimated treatment effect (RR) derived from direct evidence; indirect: estimated treatment effect (RR) derived from indirect evidence; RoR: Ratio of Ratios (direct versus indirect); z: z-value of test for disagreement (direct versus indirect): Incoherence p-value: p-value of test for disagreement (direct versus indirect)
Table 2 Direct, indirect, and network treatment estimates for lab-confirmed influenza.

| **Comparison** | **k** | **Prop** | **NMA (95% CI)** | **Direct (95% CI)** | **Indirect (95% CI)** | **Diff (95% CI)** | **z** | **Incoherence p-value** |
| --- | --- | --- | --- | --- | --- | --- | --- | --- |
| Amantadine vs. Baloxavir | 0 | 0 | 0.92 [0.40; 2.12] | NA | 0.92 [0.40; 2.12] | NA | NA | NA |
| Amantadine vs. Laninamivir | 0 | 0 | 0.86 [0.48; 1.53] | NA | 0.86 [0.48; 1.53] | NA | NA | NA |
| Amantadine vs. Oseltamivir | 0 | 0 | 0.78 [0.45; 1.36] | NA | 0.78 [0.45; 1.36] | NA | NA | NA |
| Amantadine vs. Placebo/stardard care | 6 | 0.94 | 0.46 [0.31; 0.67] | 0.45 [0.30; 0.67] | 0.56 [0.12; 2.67] | 0.81 [0.16; 4.10] | -0.25 | 0.8002 |
| Amantadine vs. Rimantadine | 2 | 0.50 | 0.83 [0.52; 1.33] | 1.04 [0.54; 2.03] | 0.66 [0.34; 1.28] | 1.58 [0.62; 4.03] | 0.96 | 0.3379 |
| Amantadine vs. Zanamivir | 0 | 0 | 0.80 [0.50; 1.28] | NA | 0.80 [0.50; 1.28] | NA | NA | NA |
| Baloxavir vs. Laninamivir | 0 | 0 | 0.93 [0.39; 2.21] | NA | 0.93 [0.39; 2.21] | NA | NA | NA |
| Baloxavir vs. Oseltamivir | 0 | 0 | 0.85 [0.37; 1.98] | NA | 0.85 [0.37; 1.98] | NA | NA | NA |
| Baloxavir vs. Placebo/stardard care | 1 | 1.00 | 0.50 [0.24; 1.05] | 0.50 [0.24; 1.05] | NA | NA | NA | NA |
| Baloxavir vs. Rimantadine | 0 | 0 | 0.90 [0.40; 2.06] | NA | 0.90 [0.40; 2.06] | NA | NA | NA |
| Baloxavir vs. Zanamivir | 0 | 0 | 0.87 [0.39; 1.93] | NA | 0.87 [0.39; 1.93] | NA | NA | NA |
| Laninamivir vs. Oseltamivir | 0 | 0 | 0.91 [0.51; 1.64] | NA | 0.91 [0.51; 1.64] | NA | NA | NA |
| Laninamivir vs. Placebo/stardard care | 3 | 1.00 | 0.53 [0.35; 0.82] | 0.53 [0.35; 0.82] | NA | NA | NA | NA |
| Laninamivir vs. Rimantadine | 0 | 0 | 0.97 [0.56; 1.68] | NA | 0.97 [0.56; 1.68] | NA | NA | NA |
| Laninamivir vs. Zanamivir | 0 | 0 | 0.93 [0.55; 1.56] | NA | 0.93 [0.55; 1.56] | NA | NA | NA |
| Oseltamivir vs. Placebo/stardard care | 6 | 1.00 | 0.58 [0.39; 0.86] | 0.58 [0.39; 0.86] | NA | NA | NA | NA |
| Oseltamivir vs. Rimantadine | 0 | 0 | 1.06 [0.63; 1.79] | NA | 1.06 [0.63; 1.79] | NA | NA | NA |
| Oseltamivir vs. Zanamivir | 0 | 0 | 1.02 [0.63; 1.66] | NA | 1.02 [0.63; 1.66] | NA | NA | NA |
| Rimantadine vs. Placebo/stardard care | 8 | 0.85 | 0.55 [0.39; 0.78] | 0.52 [0.36; 0.75] | 0.76 [0.32; 1.84] | 0.68 [0.26; 1.76] | -0.80 | 0.4249 |
| Zanamivir vs. Placebo/stardard care | 10 | 0.93 | 0.57 [0.43; 0.76] | 0.62 [0.46; 0.84] | 0.21 [0.07; 0.59] | 3.00 [1.01; 8.97] | 1.97 | 0.0489 |
| Rimantadine vs. Zanamivir | 2 | 0.19 | 0.96 [0.63; 1.47] | 2.35 [0.88; 6.33] | 0.78 [0.49; 1.26] | 3.00 [1.01; 8.97] | 1.97 | 0.0489 |

Comparison: treatment comparison; k: number of studies providing direct evidence; prop: direct evidence proportion; NMA: estimated treatment effect (RR) in network meta-analysis; direct: estimated treatment effect (RR) derived from direct evidence; indirect: estimated treatment effect (RR) derived from indirect evidence; RoR: Ratio of Ratios (direct versus indirect); z: z-value of test for disagreement (direct versus indirect): Incoherence p-value: p-value of test for disagreement (direct versus indirect).

#### Table 3 Direct, indirect, and network treatment estimates for lab-confirmed asymptomatic influenza.

| **Comparison** | **k** | **Prop** | **NMA (95% CI)** | **Direct (95% CI)** | **Indirect (95% CI)** | **Diff (95% CI)** | **z** | **Incoherence p-value** |
| --- | --- | --- | --- | --- | --- | --- | --- | --- |
| Baloxavir vs. Laninamivir | 0 | 0 | 1.35 [0.61; 2.99] | NA | 1.35 [0.61; 2.99] | NA | NA | NA |
| Baloxavir vs. Oseltamivir | 0 | 0 | 1.25 [0.55; 2.86] | NA | 1.25 [0.55; 2.86] | NA | NA | NA |
| Baloxavir vs. Placebo | 1 | 1.00 | 1.08 [0.51; 2.27] | 1.08 [0.51; 2.27] | NA | NA | NA | NA |
| Baloxavir vs. Zanamivir | 0 | 0 | 1.18 [0.55; 2.54] | NA | 1.18 [0.55; 2.54] | NA | NA | NA |
| Laninamivir vs. Oseltamivir | 0 | 0 | 0.93 [0.58; 1.48] | NA | 0.93 [0.58; 1.48] | NA | NA | NA |
| Laninamivir vs. Placebo | 3 | 1.00 | 0.80 [0.60; 1.08] | 0.80 [0.60; 1.08] | NA | NA | NA | NA |
| Laninamivir vs. Zanamivir | 0 | 0 | 0.88 [0.62; 1.25] | NA | 0.88 [0.62; 1.25] | NA | NA | NA |
| Oseltamivir vs. Placebo | 5 | 1.00 | 0.86 [0.60; 1.23] | 0.86 [0.60; 1.23] | NA | NA | NA | NA |
| Oseltamivir vs. Zanamivir | 0 | 0 | 0.94 [0.63; 1.42] | NA | 0.94 [0.63; 1.42] | NA | NA | NA |
| Zanamivir vs. Placebo | 9 | 1.00 | 0.91 [0.76; 1.10] | 0.91 [0.76; 1.10] | NA | NA | NA | NA |

Comparison: treatment comparison; k: number of studies providing direct evidence; prop: direct evidence proportion; NMA: estimated treatment effect (RR) in network meta-analysis; direct: estimated treatment effect (RR) derived from direct evidence; indirect: estimated treatment effect (RR) derived from indirect evidence; RoR: Ratio of Ratios (direct versus indirect); z: z-value of test for disagreement (direct versus indirect): Incoherence p-value: p-value of test for disagreement (direct versus indirect).

#### Table 4 Direct, indirect, and network treatment estimates for mortality.

| **Comparison** | **k** | **Prop** | **NMA (95% CI)** | **Direct (95% CI)** | **Indirect (95% CI)** | **Diff (95% CI)** | **z** | **Incoherence p-value** |
| --- | --- | --- | --- | --- | --- | --- | --- | --- |
| Amantadine vs. Baloxavir | 0 | 0 | 0.008 [-0.006; 0.022] | NA | 0.008 [-0.006; 0.022] | NA | NA | NA |
| Amantadine vs. Laninamivir | 0 | 0 | 0.008 [-0.010; 0.025] | NA | 0.008 [-0.010; 0.025] | NA | NA | NA |
| Amantadine vs. Oseltamivir | 0 | 0 | 0.008 [-0.006; 0.021] | NA | 0.008 [-0.006; 0.021] | NA | NA | NA |
| Amantadine vs. Placebo | 1 | 1.00 | 0.008 [-0.005; 0.021] | 0.008 [-0.005; 0.021] | NA | NA | NA | NA |
| Amantadine vs. Rimantadine | 0 | 0 | 0.004 [-0.013; 0.022] | NA | 0.004 [-0.013; 0.022] | NA | NA | NA |
| Amantadine vs. Zanamivir | 0 | 0 | 0.007 [-0.006; 0.020] | NA | 0.007 [-0.006; 0.020] | NA | NA | NA |
| Baloxavir vs. Laninamivir | 0 | 0 | 0.000 [-0.013; 0.013] | NA | 0.000 [-0.013; 0.013] | NA | NA | NA |
| Baloxavir vs. Oseltamivir | 0 | 0 | -0.000 [-0.006; 0.006] | NA | -0.000 [-0.006; 0.006] | NA | NA | NA |
| Baloxavir vs. Placebo | 1 | 1.00 | -0.000 [-0.005; 0.005] | -0.000 [-0.005; 0.005] | NA | NA | NA | NA |
| Baloxavir vs. Rimantadine | 0 | 0 | -0.004 [-0.016; 0.009] | NA | -0.004 [-0.016; 0.009] | NA | NA | NA |
| Baloxavir vs. Zanamivir | 0 | 0 | -0.001 [-0.006; 0.005] | NA | -0.001 [-0.006; 0.005] | NA | NA | NA |
| Laninamivir vs. Oseltamivir | 0 | 0 | -0.000 [-0.012; 0.012] | NA | -0.000 [-0.012; 0.012] | NA | NA | NA |
| Laninamivir vs. Placebo | 3 | 1.00 | -0.000 [-0.011; 0.011] | -0.000 [-0.011; 0.011] | NA | NA | NA | NA |
| Laninamivir vs. Rimantadine | 0 | 0 | -0.004 [-0.020; 0.013] | NA | -0.004 [-0.020; 0.013] | NA | NA | NA |
| Laninamivir vs. Zanamivir | 0 | 0 | -0.001 [-0.012; 0.011] | NA | -0.001 [-0.012; 0.011] | NA | NA | NA |
| Oseltamivir vs. Placebo | 5 | 1.00 | 0.000 [-0.002; 0.002] | 0.000 [-0.002; 0.002] | NA | NA | NA | NA |
| Oseltamivir vs. Rimantadine | 0 | 0 | -0.004 [-0.016; 0.008] | NA | -0.004 [-0.016; 0.008] | NA | NA | NA |
| Oseltamivir vs. Zanamivir | 0 | 0 | -0.001 [-0.003; 0.002] | NA | -0.001 [-0.003; 0.002] | NA | NA | NA |
| Rimantadine vs. Placebo | 1 | 0.04 | 0.004 [-0.008; 0.015] | 0.069 [ 0.013; 0.126] | 0.001 [-0.011; 0.013] | 0.069 [ 0.011; 0.126] | 2.34 | 0.0191 |
| Zanamivir vs. Placebo | 5 | 1 | 0.001 [-0.001; 0.002] | 0.001 [-0.001; 0.002] | 0.069 [ 0.012; 0.127] | -0.069 [-0.126; -0.011] | -2.34 | 0.0191 |
| Rimantadine vs. Zanamivir | 1 | 0.96 | 0.003 [-0.009; 0.015] | 0.000 [-0.012; 0.012] | 0.069 [ 0.013; 0.125] | -0.069 [-0.126; -0.011] | -2.34 | 0.0191 |

Comparison: treatment comparison; k: number of studies providing direct evidence; prop: direct evidence proportion; NMA: estimated treatment effect (RD) in network meta-analysis; direct: estimated treatment effect (RD) derived from direct evidence; indirect: estimated treatment effect (RD) derived from indirect evidence; Diff: difference between direct and indirect treatment estimates; z: z-value of test for disagreement (direct versus indirect): Incoherence p-value: p-value of test for disagreement (direct versus indirect).

#### Table 5 Direct, indirect, and network treatment estimates for adverse events related to drugs.

| **Comparison** | **k** | **Prop** | **NMA (95% CI)** | **Direct (95% CI)** | **Indirect (95% CI)** | **Diff (95% CI)** | **z** | **Incoherence p-value** |
| --- | --- | --- | --- | --- | --- | --- | --- | --- |
| Baloxavir vs. Laninamivir | 0 | 0 | 0.83 [0.25; 2.76] | NA | 0.83 [0.25; 2.76] | NA | NA | NA |
| Baloxavir vs. Placebo | 1 | 1.00 | 1.17 [0.40; 3.45] | 1.17 [0.40; 3.45] | NA | NA | NA | NA |
| Baloxavir vs. Rimantadine | 0 | 0 | 1.16 [0.38; 3.57] | NA | 1.16 [0.38; 3.57] | NA | NA | NA |
| Baloxavir vs. Zanamivir | 0 | 0 | 1.10 [0.36; 3.33] | NA | 1.10 [0.36; 3.33] | NA | NA | NA |
| Laninamivir vs. Placebo | 2 | 1.00 | 1.40 [0.84; 2.35] | 1.40 [0.84; 2.35] | NA | NA | NA | NA |
| Laninamivir vs. Rimantadine | 0 | 0 | 1.39 [0.76; 2.54] | NA | 1.39 [0.76; 2.54] | NA | NA | NA |
| Laninamivir vs. Zanamivir | 0 | 0 | 1.32 [0.74; 2.34] | NA | 1.32 [0.74; 2.34] | NA | NA | NA |
| Rimantadine vs. Placebo | 0 | 0 | 1.01 [0.74; 1.37] | NA | 1.01 [0.74; 1.37] | NA | NA | NA |
| Zanamivir vs. Placebo | 7 | 1 | 1.07 [0.83; 1.38] | 1.07 [0.83; 1.38] | NA | NA | NA | NA |
| Rimantadine vs. Zanamivir | 1 | 1 | 0.95 [0.80; 1.12] | 0.95 [0.80; 1.12] | NA | NA | NA | NA |

Comparison: treatment comparison; k: number of studies providing direct evidence; prop: direct evidence proportion; NMA: estimated treatment effect (RR) in network meta-analysis; direct: estimated treatment effect (RR) derived from direct evidence; indirect: estimated treatment effect (RR) derived from indirect evidence; RoR: Ratio of Ratios (direct versus indirect); z: z-value of test for disagreement (direct versus indirect): Incoherence p-value: p-value of test for disagreement (direct versus indirect).

#### Table 6 Direct, indirect, and network treatment estimates for serious adverse events.

| **Comparison** | **k** | **Prop** | **NMA (95% CI)** | **Direct (95% CI)** | **Indirect (95% CI)** | **Diff (95% CI)** | **z** | **Incoherence p-value** |
| --- | --- | --- | --- | --- | --- | --- | --- | --- |
| Amantadine vs. Baloxavir | 0 | 0 | 0.003 [-0.017; 0.022] | NA | 0.003 [-0.017; 0.022] | NA | NA | NA |
| Amantadine vs. Laninamivir | 0 | 0 | -0.000 [-0.021; 0.021] | NA | -0.000 [-0.021; 0.021] | NA | NA | NA |
| Amantadine vs. Oseltamivir | 0 | 0 | -0.003 [-0.021; 0.016] | NA | -0.003 [-0.021; 0.016] | NA | NA | NA |
| Amantadine vs. Placebo | 1 | 1.00 | -0.000 [-0.018; 0.018] | -0.000 [-0.018; 0.018] | NA | NA | NA | NA |
| Amantadine vs. Rimantadine | 0 | 0 | 0.004 [-0.025; 0.032] | NA | 0.004 [-0.025; 0.032] | NA | NA | NA |
| Amantadine vs. Zanamivir | 0 | 0 | -0.000 [-0.019; 0.018] | NA | -0.000 [-0.019; 0.018] | NA | NA | NA |
| Baloxavir vs. Laninamivir | 0 | 0 | -0.003 [-0.016; 0.011] | NA | -0.003 [-0.016; 0.011] | NA | NA | NA |
| Baloxavir vs. Oseltamivir | 0 | 0 | -0.006 [-0.014; 0.003] | NA | -0.006 [-0.014; 0.003] | NA | NA | NA |
| Baloxavir vs. Placebo | 1 | 1.00 | -0.003 [-0.010; 0.005] | -0.003 [-0.010; 0.005] | NA | NA | NA | NA |
| Baloxavir vs. Rimantadine | 0 | 0 | 0.001 [-0.022; 0.024] | NA | 0.001 [-0.022; 0.024] | NA | NA | NA |
| Baloxavir vs. Zanamivir | 0 | 0 | -0.003 [-0.011; 0.005] | NA | -0.003 [-0.011; 0.005] | NA | NA | NA |
| Laninamivir vs. Oseltamivir | 0 | 0 | -0.003 [-0.015; 0.009] | NA | -0.003 [-0.015; 0.009] | NA | NA | NA |
| Laninamivir vs. Placebo | 1 | 1.00 | 0.000 [-0.011; 0.011] | 0.000 [-0.011; 0.011] | NA | NA | NA | NA |
| Laninamivir vs. Rimantadine | 0 | 0 | 0.004 [-0.021; 0.029] | NA | 0.004 [-0.021; 0.029] | NA | NA | NA |
| Laninamivir vs. Zanamivir | 0 | 0 | -0.000 [-0.012; 0.012] | NA | -0.000 [-0.012; 0.012] | NA | NA | NA |
| Oseltamivir vs. Placebo | 5 | 1.00 | 0.003 [-0.002; 0.007] | 0.003 [-0.002; 0.007] | NA | NA | NA | NA |
| Oseltamivir vs. Rimantadine | 0 | 0 | 0.006 [-0.016; 0.029] | NA | 0.006 [-0.016; 0.029] | NA | NA | NA |
| Oseltamivir vs. Zanamivir | 0 | 0 | 0.003 [-0.003; 0.008] | NA | 0.003 [-0.003; 0.008] | NA | NA | NA |
| Rimantadine vs. Placebo | 0 | 0 | -0.004 [-0.026; 0.019] | NA | -0.004 [-0.026; 0.019] | NA | NA | NA |
| Zanamivir vs. Placebo | 5 | 1 | 0.000 [-0.003; 0.004] | 0.000 [-0.003; 0.004] | NA | NA | NA | NA |
| Rimantadine vs. Zanamivir | 1 | 1.00 | -0.004 [-0.026; 0.018] | -0.004 [-0.026; 0.018] | NA | NA | NA | NA |

Comparison: treatment comparison; k: number of studies providing direct evidence; prop: direct evidence proportion; NMA: estimated treatment effect (RD) in network meta-analysis; direct: estimated treatment effect (RD) derived from direct evidence; indirect: estimated treatment effect (RD) derived from indirect evidence; Diff: difference between direct and indirect treatment estimates; z: z-value of test for disagreement (direct versus indirect): Incoherence p-value: p-value of test for disagreement (direct versus indirect).

### Appendix 11 Network estimates with GRADE ratings

#### Table 1 Network estimates with GRADE ratings for lab-confirmed symptomatic influenza (seasonal influenza)

| **Comparison groups** | | **Network relative estimate (RR)** | | | **Low-risk populations** | | | | **High-risk populations** | | | |
| --- | --- | --- | --- | --- | --- | --- | --- | --- | --- | --- | --- | --- |
|  |  |  |  |  | **Network absolute** **estimate (per 1000)** | | | **GRADE rating** | **Network absolute estimate (per 1000)** | | | **GRADE rating** |
| **Treatment 1** | **Treatment 2** | **Point estimate** | **95% CI lower limit** | **95% CI upper limit** | **Point estimate** | **95% CI lower limit** | **95% CI upper limit** |  | **Point estimate** | **95% CI lower limit** | **95% CI upper limit** |  |
| Baloxavir | Laninamivir | 0.98 | 0.48 | 2.03 | -2 | -41 | 80 | Moderate | -2 | -41 | 80 | Moderate |
| Baloxavir | Oseltamivir | 1.06 | 0.50 | 2.26 | 5 | -39 | 98 | Moderate | 5 | -39 | 98 | Moderate |
| Baloxavir | Placebo | 0.42 | 0.23 | 0.79 | -44 | -60 | -16 | Moderate | -44 | -60 | -16 | Moderate |
| Baloxavir | Rimantadine | 0.56 | 0.17 | 1.80 | -34 | -65 | 62 | Moderate | -34 | -65 | 62 | Moderate |
| Baloxavir | Zanamivir | 1.21 | 0.60 | 2.47 | 16 | -31 | 115 | Moderate | 16 | -31 | 115 | Moderate |
| Laninamivir | Oseltamivir | 1.08 | 0.61 | 1.91 | 6 | -30 | 71 | Moderate | 6 | -30 | 71 | Moderate |
| Laninamivir | Placebo | 0.43 | 0.30 | 0.63 | -44 | -55 | -29 | Moderate | -44 | -55 | -29 | Moderate |
| Laninamivir | Rimantadine | 0.57 | 0.20 | 1.64 | -34 | -62 | 50 | Moderate | -34 | -62 | 50 | Moderate |
| Laninamivir | Zanamivir | 1.23 | 0.74 | 2.05 | 18 | -20 | 82 | Moderate | 18 | -20 | 82 | Moderate |
| Oseltamivir | Placebo | 0.40 | 0.26 | 0.62 | -47 | -58 | -30 | Moderate | -47 | -58 | -30 | Moderate |
| Oseltamivir | Rimantadine | 0.53 | 0.18 | 1.56 | -37 | -64 | 44 | Moderate | -37 | -64 | 44 | Moderate |
| Oseltamivir | Zanamivir | 1.14 | 0.66 | 1.99 | 11 | -27 | 77 | Moderate | 11 | -27 | 77 | Moderate |
| Rimantadine | Placebo | 0.76 | 0.28 | 2.06 | -19 | -56 | 83 | Moderate | -19 | -56 | 83 | Moderate |
| Zanamivir | Placebo | 0.35 | 0.25 | 0.50 | -51 | -58 | -39 | Moderate | -51 | -58 | -39 | Moderate |
| Zanamivir | Rimantadine | 2.17 | 0.86 | 5.53 | 91 | -11 | 353 | Moderate | 91 | -11 | 353 | Moderate |

NA, not applicable.

#### Table 2 Network estimates with GRADE ratings for lab-confirmed influenza (seasonal influenza)

| **Comparison groups** | | | **Network relative estimate** | | | **Network absolute estimate (per 1000)** | | | **GRADE rating** |
| --- | --- | --- | --- | --- | --- | --- | --- | --- | --- |
| **Treatment 1** | **Treatment 2** | **Point estimate** | | **95% CI lower limit** | **95% CI upper limit** | **Point estimate** | **95% CI lower limit** | **95% CI upper limit** |  |
| Amantadine | Baloxavir | 0.92 | | 0.40 | 2.12 | -14 | -106 | 198 | Moderate |
| Amantadine | Laninamivir | 0.86 | | 0.48 | 1.53 | -80 | -108 | 94 | Moderate |
| Amantadine | Oseltamivir | 0.78 | | 0.45 | 1.36 | -39 | -97 | 64 | Moderate |
| Amantadine | Placebo/standard care | 0.46 | | 0.31 | 0.67 | -96 | -122 | -58 | Moderate |
| Amantadine | Rimantadine | 0.83 | | 0.52 | 1.33 | -30 | -85 | 58 | Moderate |
| Amantadine | Zanamivir | 0.80 | | 0.50 | 1.28 | -35 | -88 | 50 | Moderate |
| Baloxavir | Laninamivir | 0.93 | | 0.39 | 2.21 | -12 | -108 | 214 | Moderate |
| Baloxavir | Oseltamivir | 0.85 | | 0.37 | 1.98 | -27 | -112 | 173 | Moderate |
| Baloxavir | Placebo/standard care | 0.50 | | 0.24 | 1.05 | -88 | -129 | 9 | Moderate |
| Baloxavir | Rimantadine | 0.90 | | 0.40 | 2.06 | -18 | -106 | 188 | Moderate |
| Baloxavir | Zanamivir | 0.87 | | 0.39 | 1.93 | -23 | -108 | 165 | Moderate |
| Laninamivir | Oseltamivir | 0.91 | | 0.51 | 1.64 | -16 | -87 | 113 | Moderate |
| Laninamivir | Placebo/standard care | 0.53 | | 0.35 | 0.82 | -83 | -115 | -32 | Moderate |
| Laninamivir | Rimantadine | 0.97 | | 0.56 | 1.68 | -5 | -78 | 120 | Moderate |
| Laninamivir | Zanamivir | 0.93 | | 0.55 | 1.56 | -12 | -80 | 99 | Moderate |
| Oseltamivir | Placebo/standard care | 0.58 | | 0.39 | 0.86 | -74 | -108 | -25 | Moderate |
| Oseltamivir | Rimantadine | 1.06 | | 0.63 | 1.79 | 11 | -65 | 140 | Moderate |
| Oseltamivir | Zanamivir | 1.02 | | 0.63 | 1.66 | 4 | -65 | 117 | Moderate |
| Rimantadine | Placebo/standard care | 0.55 | | 0.39 | 0.78 | -80 | -108 | -39 | Low |
| Zanamivir | Placebo/standard care | 0.57 | | 0.43 | 0.76 | -74 | -101 | -42 | Low |
| Zanamivir | Rimantadine | 0.96 | | 0.63 | 1.47 | -7 | -65 | 83 | Moderate |

#### Table 3 Network estimates with GRADE ratings for lab-confirmed asymptomatic influenza (seasonal influenza)

| **Comparison groups** | | | **Network relative estimate** | | | **Network absolute estimate** | | | **GRADE rating** |
| --- | --- | --- | --- | --- | --- | --- | --- | --- | --- |
| **Treatment 1** | **Treatment 2** | **Point estimate** | | **95% CI lower limit** | **95% CI upper limit** | **Point estimate** | **95% CI lower limit** | **95% CI upper limit** |  |
| Baloxavir | Laninamivir | 1.35 | | 0.61 | 2.99 | 17 | -19 | 98 | Moderate |
| Baloxavir | Oseltamivir | 1.25 | | 0.55 | 2.86 | 12 | -22 | 91 | Moderate |
| Baloxavir | Placebo | 1.08 | | 0.51 | 2.27 | 4 | -24 | 62 | Moderate |
| Baloxavir | Zanamivir | 1.18 | | 0.55 | 2.54 | 9 | -22 | 75 | Moderate |
| Laninamivir | Oseltamivir | 0.93 | | 0.58 | 1.48 | -3 | -21 | 24 | High |
| Laninamivir | Placebo | 0.80 | | 0.60 | 1.08 | -10 | -20 | 4 | High |
| Laninamivir | Zanamivir | 0.88 | | 0.62 | 1.25 | -6 | -19 | 12 | High |
| Oseltamivir | Placebo | 0.86 | | 0.60 | 1.23 | -7 | -20 | 11 | High |
| Oseltamivir | Zanamivir | 0.94 | | 0.63 | 1.42 | -3 | -18 | 21 | High |
| Zanamivir | Placebo | 0.91 | | 0.76 | 1.10 | -4 | -12 | 5 | High |

NA, not applicable.

#### Table 4 Network estimates with GRADE ratings for all-cause mortality (seasonal influenza)

| **Comparison groups** | | | **Network relative estimate (RR)** | | | **Network absolute estimate (per 1000)** | | | **GRADE rating** |
| --- | --- | --- | --- | --- | --- | --- | --- | --- | --- |
| **Treatment 1** | **Treatment 2** | **Point estimate** | | **95% CI lower limit** | **95% CI upper limit** | **Point estimate** | **95% CI lower limit** | **95% CI upper limit** |  |
| Amantadine | Baloxavir | NA | | NA | NA | 8 | -6 | 22 | Moderate |
| Amantadine | Laninamivir | NA | | NA | NA | 8 | -10 | 25 | Moderate |
| Amantadine | Oseltamivir | NA | | NA | NA | 8 | -6 | 21 | Moderate |
| Amantadine | Placebo | NA | | NA | NA | 8 | -5 | 21 | Moderate |
| Amantadine | Rimantadine | NA | | NA | NA | 4 | -13 | 22 | Moderate |
| Amantadine | Zanamivir | NA | | NA | NA | 7 | -6 | 20 | Moderate |
| Baloxavir | Laninamivir | NA | | NA | NA | 0 | -13 | 13 | Moderate |
| Baloxavir | Oseltamivir | NA | | NA | NA | 0 | -6 | 6 | Moderate |
| Baloxavir | Placebo | NA | | NA | NA | 0 | -5 | 5 | Moderate |
| Baloxavir | Rimantadine | NA | | NA | NA | -4 | -16 | 9 | Moderate |
| Baloxavir | Zanamivir | NA | | NA | NA | -1 | -6 | 5 | Moderate |
| Laninamivir | Oseltamivir | NA | | NA | NA | 0 | --12 | 12 | Moderate |
| Laninamivir | Placebo | NA | | NA | NA | 0 | -11 | 11 | Moderate |
| Laninamivir | Rimantadine | NA | | NA | NA | -4 | -20 | 13 | Moderate |
| Laninamivir | Zanamivir | NA | | NA | NA | -1 | -12 | 11 | Moderate |
| Oseltamivir | Placebo | NA | | NA | NA | 0 | -2 | 2 | High |
| Oseltamivir | Rimantadine | NA | | NA | NA | -4 | -16 | 8 | Moderate |
| Oseltamivir | Zanamivir | NA | | NA | NA | -1 | -3 | 2 | Moderate |
| Rimantadine | Placebo | NA | | NA | NA | 4 | -8 | 15 | Very Low |
| Zanamivir | Placebo | NA | | NA | NA | 1 | -1 | 2 | Very Low |
| Zanamivir | Rimantadine | NA | | NA | NA | 3 | -9 | 15 | Very Low |

NA, not applicable.

#### Table 5 Network estimates with GRADE ratings for adverse events related to drugs (seasonal influenza and zoonotic influenza)

| **Comparison groups** | | | **Network relative estimate** | | | **Network absolute estimate** | | | **GRADE rating** |
| --- | --- | --- | --- | --- | --- | --- | --- | --- | --- |
| **Treatment 1** | **Treatment 2** | **Point estimate** | | **95% CI lower limit** | **95% CI upper limit** | **Point estimate** | **95% CI lower limit** | **95% CI upper limit** |  |
| Baloxavir | Laninamivir | 0.83 | | 0.25 | 2.76 | -6 | -27 | 63 | Low |
| Baloxavir | Placebo | 1.17 | | 0.40 | 3.45 | 6 | -22 | 88 | Low |
| Baloxavir | Rimantadine | 1.16 | | 0.38 | 3.75 | 6 | -22 | 99 | Low |
| Baloxavir | Zanamivir | 1.10 | | 0.36 | 3.33 | 4 | -23 | 84 | Low |
| Laninamivir | Placebo | 1.40 | | 0.84 | 2.35 | 14 | -6 | 49 | Moderate |
| Laninamivir | Rimantadine | 1.39 | | 0.76 | 2.54 | 14 | -9 | 55 | Moderate |
| Laninamivir | Zanamivir | 1.32 | | 0.74 | 2.34 | 12 | -9 | 48 | Moderate |
| Rimantadine | Placebo | 1.01 | | 0.74 | 1.37 | 0 | -9 | 13 | Moderate |
| Zanamivir | Placebo | 1.07 | | 0.83 | 1.38 | 3 | -6 | 14 | Moderate |
| Rimantadine | Zanamivir | 0.95 | | 0.80 | 1.12 | -2 | -7 | 4 | High |

#### Table 6 Network estimates with GRADE ratings for serious adverse events (seasonal influenza and zoonotic influenza)

| **Comparison groups** | | | **Network relative estimate** | | | **Network absolute estimate (per 1000)** | | | **GRADE rating** |
| --- | --- | --- | --- | --- | --- | --- | --- | --- | --- |
| **Treatment 1** | **Treatment 2** | **Point estimate** | | **95% CI lower limit** | **95% CI upper limit** | **Point estimate** | **95% CI lower limit** | **95% CI upper limit** |  |
| Amantadine | Baloxavir | NA | | NA | NA | 3 | -17 | 22 | Low |
| Amantadine | Laninamivir | NA | | NA | NA | -0 | -21 | 21 | Low |
| Amantadine | Oseltamivir | NA | | NA | NA | -3 | -21 | 16 | Low |
| Amantadine | Placebo | NA | | NA | NA | -0 | -18 | 18 | Low |
| Amantadine | Rimantadine | NA | | NA | NA | 4 | -25 | 32 | Low |
| Amantadine | Zanamivir | NA | | NA | NA | -0 | -19 | 18 | Low |
| Baloxavir | Laninamivir | NA | | NA | NA | -3 | -16 | 11 | Low |
| Baloxavir | Oseltamivir | NA | | NA | NA | -6 | -14 | 3 | Low |
| Baloxavir | Placebo | NA | | NA | NA | -3 | -10 | 5 | Low |
| Baloxavir | Rimantadine | NA | | NA | NA | 1 | -22 | 24 | Low |
| Baloxavir | Zanamivir | NA | | NA | NA | -3 | -11 | 5 | Low |
| Laninamivir | Oseltamivir | NA | | NA | NA | -3 | --15 | 9 | Low |
| Laninamivir | Placebo | NA | | NA | NA | 0 | -11 | 11 | Low |
| Laninamivir | Rimantadine | NA | | NA | NA | 4 | -21 | 29 | Low |
| Laninamivir | Zanamivir | NA | | NA | NA | -0 | -12 | 12 | Low |
| Oseltamivir | Placebo | NA | | NA | NA | 3 | -2 | 7 | Moderate |
| Oseltamivir | Rimantadine | NA | | NA | NA | 6 | -16 | 29 | Low |
| Oseltamivir | Zanamivir | NA | | NA | NA | 3 | -3 | 8 | Low |
| Rimantadine | Placebo | NA | | NA | NA | -4 | -26 | 19 | Moderate |
| Zanamivir | Placebo | NA | | NA | NA | 0 | -3 | 4 | High |
| Zanamivir | Rimantadine | NA | | NA | NA | -4 | -26 | 18 | Low |

NA, not applicable.

#### Table 7 Network estimates with GRADE ratings for lab-confirmed symptomatic influenza (zoonotic influenza)

| **Comparison groups** | | **Network relative estimate** | | | **Network absolute estimate (per 1000)** | | | **GRADE rating** |
| --- | --- | --- | --- | --- | --- | --- | --- | --- |
| **Treatment 1** | **Treatment 2** | **Point estimate** | **95% CI lower limit** | **95% CI upper limit** | **Point estimate** | **95% CI lower limit** | **95% CI upper limit** |  |
| Baloxavir | Laninamivir | 0.98 | 0.48 | 2.03 | NA | NA | NA | NA |
| Baloxavir | Oseltamivir | 1.06 | 0.50 | 2.26 | NA | NA | NA | NA |
| Baloxavir | Placebo | 0.42 | 0.23 | 0.79 | -17 | -23 | -6 | Low |
| Baloxavir | Rimantadine | 0.56 | 0.17 | 1.80 | NA | NA | NA | NA |
| Baloxavir | Zanamivir | 1.21 | 0.60 | 2.47 | NA | NA | NA | NA |
| Laninamivir | Oseltamivir | 1.08 | 0.61 | 1.91 | NA | NA | NA | NA |
| Laninamivir | Placebo | 0.43 | 0.30 | 0.63 | -17 | -21 | -11 | Low |
| Laninamivir | Rimantadine | 0.57 | 0.20 | 1.64 | NA | NA | NA | NA |
| Laninamivir | Zanamivir | 1.23 | 0.74 | 2.05 | NA | NA | NA | NA |
| Oseltamivir | Placebo | 0.40 | 0.26 | 0.62 | -18 | -22 | -11 | Low |
| Oseltamivir | Rimantadine | 0.53 | 0.18 | 1.56 | NA | NA | NA | NA |
| Oseltamivir | Zanamivir | 1.14 | 0.66 | 1.99 | NA | NA | NA | NA |
| Rimantadine | Placebo | 0.76 | 0.28 | 2.06 | -7 | -22 | 32 | Very Low |
| Zanamivir | Placebo | 0.35 | 0.25 | 0.50 | -19 | -22 | -15 | Low |
| Zanamivir | Rimantadine | 2.17 | 0.86 | 5.53 | NA | NA | NA | NA |

NA, not applicable. *Estimate was informed by direct evidence because of serious incoherence in these comparisons.

#### Table 8 Network estimates with GRADE ratings for admission to hospital (zoonotic influenza)

| **Comparison groups** | | **Network relative estimate** | | | **Network absolute estimate (per 1000)** | | | **GRADE rating** |
| --- | --- | --- | --- | --- | --- | --- | --- | --- |
| **Treatment 1** | **Treatment 2** | **Point estimate** | **95% CI lower limit** | **95% CI upper limit** | **Point estimate** | **95% CI lower limit** | **95% CI upper limit** |  |
| Baloxavir | Placebo | NA | NA | NA | -14 | -18 | -5 | Very Low |
| Laninamivir | Placebo | NA | NA | NA | -14 | -17 | -9 | Very Low |
| Oseltamivir | Placebo | NA | NA | NA | -15 | -18 | 9 | Very Low |
| Zanamivir | Placebo | NA | NA | NA | -15 | -18 | -12 | Very Low |

NA, not applicable.

#### Table 9 Network estimates with GRADE ratings for all-cause mortality (zoonotic influenza)

| **Comparison groups** | | **Network relative estimate** | | | **Network absolute estimate (per 1000)** | | | **GRADE rating** |
| --- | --- | --- | --- | --- | --- | --- | --- | --- |
| **Treatment 1** | **Treatment 2** | **Point estimate** | **95% CI lower limit** | **95% CI upper limit** | **Point estimate** | **95% CI lower limit** | **95% CI upper limit** |  |
| Baloxavir | Placebo | NA | NA | NA | -3 | -4 | -1 | Very Low |
| Laninamivir | Placebo | NA | NA | NA | -3 | -3 | -2 | Very Low |
| Oseltamivir | Placebo | NA | NA | NA | -3 | -4 | 2 | Very Low |
| Zanamivir | Placebo | NA | NA | NA | -3 | -4 | -2 | Very Low |

NA, not applicable.

### Appendix 12 Summary of findings table for antivirals versus placebo/standard care or placebo.

#### Table 1 Summary of findings table for Zanamivir versus placebo or standard care for prophylaxis against seasonal influenza

| **Outcome**  Timeframe | **Study results and measurements** | **Absolute effect estimates** | | **Certainty of the Evidence**  (Quality of evidence) | **Summary** |
| --- | --- | --- | --- | --- | --- |
|  |  | Placebo/standard care | Zanamivir |  |  |
| Lab-confirmed symptomatic influenza | Relative risk: 0.35  (CI 95% 0.25 - 0.5)  Based on data from 8104 participants in 9 studies | 78  per 1000 | 27  per 1000 | **Moderate**  Due to serious imprecision | Low-risk populations and high-risk populations: compared to placebo, zanamivir probably decreases lab-confirmed symptomatic influenza. |
|  |  | Difference: 51 fewer per 1000  (CI 95% 58 fewer - 39 fewer) | |  |  |
| Lab-confirmed influenza | Relative risk: 0.57  (CI 95% 0.43 – 0.76)  Based on data from 8156 participants in 10 studies | 177  per 1000 | 101  per 1000 | **Low**  Due to serious incoherent and imprecision | Compared to placebo/standard of care, zanamivir may decrease lab-confirmed influenza. |
|  |  | Difference: 74 fewer per 1000  (CI 95% 101 fewer - 42 fewer) | |  |  |
| Lab-confirmed asymptomatic influenza | Relative risk: 0.91  (CI 95% 0.76 – 1.10)  Based on data from 8104 participants in 9 studies | 49  per 1000 | 45  per 1000 | **High** | Compared to placebo, zanamivir has little or no effect on lab-confirmed asymptomatic influenza. |
|  |  | Difference: 4 fewer per 1000  (CI 95% 12 fewer – 5 more) | |  |  |
| Admission to hospital (No data from RCT) |  |  |  |  | Zanamivir has little or no effect on admission to hospital due to the baseline risk being lower than the minimal important difference. |
| All-cause mortality | Risk difference: 0.001  (CI 95% -0.001 - 0.002)  Based on data from 4295 participants in 5 studies | 0.2  per 1000 | 1.2  per 1000 | **Moderate**  Due to serious incoherent | Compared to placebo, zanamivir probably has little or no effect on all-cause mortality |
|  |  | Difference: 1 more per 1000  (CI 95% 1 fewer - 2 more) | |  |  |
| Adverse events related to drugs | Relative risk: 1.07  (CI 95% 0.83 - 1.38)  Based on data from 6814 participants in 8 studies | 36  per 1000 | 39  per 1000 | **Moderate**  Due to serious imprecision | Zanamivir probably has little or no effect on the risk of adverse events related to drugs. |
|  |  | Difference: 3 more per 1000  (CI 95% 6 fewer - 14 more) | |  |  |
| Serious adverse events | Risk difference: 0.0  (CI 95% -0.003 - 0.004)  Based on data from 6239 participants in 7 studies | 0.2  per 1000 | 0.2  per 1000 | **High** | Zanamivir has little or no effect on the risk of serious adverse events |
|  |  | Difference: 0 fewer per 1000  (CI 95% 3 fewer - 4 more) | |  |  |

#### Table 2 Summary of findings table for Oseltamivir versus placebo for prophylaxis against seasonal influenza

| **Outcome**  Timeframe | **Study results and measurements** | **Absolute effect estimates** | | **Certainty of the Evidence**  (Quality of evidence) | **Summary** |
| --- | --- | --- | --- | --- | --- |
|  |  | Placebo | Oseltamivir |  |  |
| Lab-confirmed symptomatic influenza | Relative risk: 0.4  (CI 95% 0.26 - 0.62)  Based on data from 3742 participants in 5 studies | 78  per 1000 | 31  per 1000 | **Moderate**  Due to serious imprecision | Low-risk populations: compared to placebo, oseltamivir probably has little or no effect on lab-confirmed symptomatic influenza. |
|  |  | Difference: 47 fewer per 1000  (CI 95% 58 fewer - 30 fewer) | |  | High-risk populations: compared to placebo, oseltamivir probably decreases lab-confirmed symptomatic influenza. |
| Lab-confirmed influenza | Relative risk: 0.58  (CI 95% 0.39- 0.86)  Based on data from 3821 participants in 6 studies | 177  per 1000 | 101  per 1000 | **Moderate**  Due to serious imprecision | Compared to placebo/standard of care, oseltamivir probably decreases lab-confirmed influenza. |
|  |  | Difference: 74 fewer per 1000  (CI 95% 108 fewer - 25 fewer) | |  |  |
| Lab-confirmed asymptomatic influenza | Relative risk: 0.86  (CI 95% 0.60 – 1.23)  Based on data from 3742 participants in 5 studies | 49  per 1000 | 42  per 1000 | **High** | Compared to placebo, oseltamivir has little or no effect on lab-confirmed asymptomatic influenza. |
|  |  | Difference: 7 fewer per 1000  (CI 95% 20 fewer – 11 more) | |  |  |
| Admission to hospital | Relative risk: 1.11  (CI 95% 0.81 - 1.52)  Based on data from 3434 participants in 4 studies | 25  per 1000 | 28  per 1000 | **Moderate**  Due to serious imprecision | Oseltamivir probably has little or no effect on admission to hospital. |
|  |  | Difference: 3 more per 1000  (CI 95% 8 fewer - 22 more) | |  |  |
| All-cause mortality | Risk difference: 0  (CI 95% -0.002 - 0.002)  Based on data from 3507 participants in 5 studies | 0.2  per 1000 | 0.2  per 1000 | **High** | Oseltamivir has little or no impact on all-cause mortality. |
|  |  | Difference: 0 fewer per 1000  (CI 95% 2 fewer - 2 more) | |  |  |
| Adverse events related to drugs (No data from RCT) |  |  |  |  | Whether oseltamivir increases adverse events related to drugs is very uncertain. |
| Serious adverse events | Risk difference: 0.0  (CI 95% -0.002 - 0.007)  Based on data from 3742 participants in 5 studies | 0.2  per 1000 | 3.2  per 1000 | **Moderate**  Due to serious imprecision | Oseltamivir probably has little or no effect on adverse events related to drugs |
|  |  | Difference: 3.0 more  (CI 95% 2.0 fewer - 7.0 more) | |  |  |

#### Table 3 Summary of findings table for laninamivir versus placebo for prophylaxis against seasonal influenza

| **Outcome**  Timeframe | **Study results and measurements** | **Absolute effect estimates** | | **Certainty of the Evidence**  (Quality of evidence) | **Summary** |
| --- | --- | --- | --- | --- | --- |
|  |  | Placebo | Laninamivir |  |  |
| Lab-confirmed symptomatic influenza | Relative risk: 0.43  (CI 95% 0.3 - 0.63)  Based on data from 2593 participants in 3 studies | 78  per 1000 | 34  per 1000 | **Moderate**  Due to serious imprecision | Low-risk populations: compared to placebo, laninamivir probably has little or no effect on lab-confirmed symptomatic influenza. |
|  |  | Difference: 44 fewer per 1000  (CI 95% 55 fewer - 29 fewer) | |  | High-risk populations: compared to placebo, laninamivir probably decreases lab-confirmed symptomatic influenza |
| Lab-confirmed influenza | Relative risk: 0.53  (CI 95% 0.35 - 0.82)  Based on data from 2593 participants in 3 studies | 177  per 1000 | 94  per 1000 | **Moderate**  Due to serious imprecision | Compared to placebo/standard of care, laninamivir probably decreases lab-confirmed influenza. |
|  |  | Difference: 83 fewer per 1000  (CI 95% 115 fewer - 32 fewer) | |  |  |
| Lab-confirmed asymptomatic influenza | Relative risk: 0.80  (CI 95% 0.60 – 1.08)  Based on data from 2593 participants in 3 studies | 49  per 1000 | 39  per 1000 | **High** | Compared to placebo, laninamivir has little or no effect on lab-confirmed asymptomatic influenza. |
|  |  | Difference: 10 fewer per 1000  (CI 95% 20 fewer – 4 more) | |  |  |
| Admission to hospital (No data from RCT) |  |  |  |  | Laninamivir has little or no effect on admission to hospital due to the baseline risk being lower than the minimal important difference. |
| All-cause mortality | Risk difference: 0  (CI 95% -0.011 - 0.011)  Based on data from 341 participants in 1 study | 0.2  per 1000 | 0.2  per 1000 | **Moderate**  Due to serious imprecision | Laninamivir probably has little or no impact on all-cause mortality. |
|  |  | Difference: 0.0 fewer per 1000  (CI 95% 11.0 fewer - 11.0 more) | |  |  |
| Adverse events related to drugs | Relative risk: 1.4  (CI 95% 0.84 - 2.35)  Based on data from 2806 participants in 3 studies | 36  per 1000 | 50  per 1000 | **Moderate**  Due to serious imprecision | Laninamivir probably increases adverse events related to drugs |
|  |  | Difference: 14 more per 1000  (CI 95% 6 fewer - 49 more) | |  |  |
| Serious adverse events | Risk difference: 0  (CI 95% -0.011 – 0.011)  Based on data from 341 participants in 1 study | 0.2  per 1000 | 0.2  per 1000 | **Low**  Due to serious risk of bias, Due to serious imprecision^1^ | Laninamivir may have little or no effect on serious adverse events |
|  |  | Difference: 0 fewer per 1000  (CI 95% 11 fewer - 11 more) | |  |  |

#### Table 4 Summary of findings table for baloxavir versus placebo for prophylaxis against seasonal influenza

| **Outcome**  Timeframe | **Study results and measurements** | **Absolute effect estimates** | | **Certainty of the Evidence**  (Quality of evidence) | **Summary** |
| --- | --- | --- | --- | --- | --- |
|  |  | Placebo | Baloxavir |  |  |
| Lab-confirmed symptomatic influenza | Relative risk: 0.43  (CI 95% 0.23 - 0.79)  Based on data from 749 participants in 1 study | 78  per 1000 | 34  per 1000 | **Moderate**  Due to serious imprecision | Low-risk populations: compared to placebo, baloxavir probably has little or no effect on lab-confirmed symptomatic influenza. |
|  |  | Difference: 44 fewer per 1000  (CI 95% 60 fewer - 16 fewer) | |  | High-risk populations: compared to placebo, baloxavir probably decreases lab-confirmed symptomatic influenza. |
| Lab-confirmed influenza | Relative risk: 0.50  (CI 95% 0.24 – 1.05)  Based on data from 749 participants in 1 study | 177  per 1000 | 89  per 1000 | **Moderate**  Due to serious imprecision | Compared to placebo/standard of care, baloxavir probably decreases lab-confirmed influenza. |
|  |  | Difference: 88 fewer per 1000  (CI 95% 129 fewer - 9 more) | |  |  |
| Lab-confirmed asymptomatic influenza | Relative risk: 1.08  (CI 95% 0.51 – 2.27)  Based on data from 749 participants in 1 study | 49  per 1000 | 53  per 1000 | **Moderate**  Due to serious imprecision | Compared to placebo, baloxavir probably has little or no effect on lab-confirmed asymptomatic influenza. |
|  |  | Difference: 4 more per 1000  (CI 95% 24 fewer - 62 more) | |  |  |
| Admission to hospital (No data from RCT) |  |  |  |  | Baloxavir has little or no effect on admission to hospital due to the baseline risk being lower than the minimal important difference. |
| All-cause mortality | Risk difference: 0  (CI 95% -0.005 – 0.005)  Based on data from 749 participants in 1 study | 0.2  per 1000 | 0.2  per 1000 | **Moderate**  Due to serious imprecision | Baloxavir probably has little or no effect on all-cause mortality |
|  |  | Difference: 0.0 fewer  (CI 95% 5.0 fewer - 5.0 more) | |  |  |
| Adverse events related to drugs | Relative risk: 1.17  (CI 95% 0.4 - 3.45)  Based on data from 749 participants in 1 study | 36  per 1000 | 42  per 1000 | **Low**  Due to serious risk of bias, Due to serious imprecision | Baloxavir may have little or no difference on adverse events related to drugs |
|  |  | Difference: 6 more per 1000  (CI 95% 22 fewer - 88 more) | |  |  |
| Serious adverse events | Risk difference: 3  (CI 95% -0.010 - 0.005)  Based on data from 749 participants in 1 study | 0.2  per 1000 | 3.2  per 1000 | **Low**  Due to serious risk of bias, Due to serious imprecision | Compared to placebo, baloxavir may have little or no effect on serious adverse events |
|  |  | Difference: 3.0 fewer  (CI 95% 10.0 fewer - 5.0 more) | |  |  |

#### Table 5 Summary of findings table for Amantadine versus placebo for prophylaxis against seasonal influenza

| **Outcome**  Timeframe | **Study results and measurements** | **Absolute effect estimates** | | **Certainty of the Evidence**  (Quality of evidence) | **Summary** |
| --- | --- | --- | --- | --- | --- |
|  |  | Placebo | Amantadine |  |  |
| Lab-confirmed symptomatic influenza |  |  |  |  | Not applicable |
| Lab-confirmed influenza | Relative risk: 0.46  (CI 95% 0.31 - 0.67)  Based on data from 1703 participants in 6 studies | 177  per 1000 | 81  per 1000 | **Moderate**  Due to heterogeneity | Compared to placebo/standard care, amantadine probably decreases lab-confirmed influenza. |
|  |  | Difference: 96 fewer per 1000  (CI 95% 122 fewer - 58 fewer) | |  |  |
| Lab-confirmed asymptomatic influenza |  |  |  |  | Not applicable |
| Admission to hospital (No data from RCT) |  |  |  |  | Not applicable |
| All-cause mortality | Risk difference: 0.008  (CI 95% -0.005 - 0.021)  Based on data from 511 participants in 1 study | 0.2  per 1000 | 8.2  per 1000 | **Very low**  Due to extremely serious imprecision | Whether amantadine increases all-cause mortality is very uncertain |
|  |  | Difference: 8.0 more  (CI 95% 5.0 fewer - 21 more) | |  |  |
| Adverse events related to drugs |  |  |  |  | Not applicable |
| Serious adverse events | Risk difference: 0  (CI 95% -0.018 -0.018)  Based on data from 215 participants in 1 study | 0.2  per 1000 | 3.2  per 1000 | **Low**  Due to serious risk of bias, Due to serious imprecision | Compared to placebo, amantadine may have little or no effect on serious adverse events |
|  |  | Difference: 0.0 fewer  (CI 95% 18.0 fewer - 18.0 more) | |  |  |

#### Table 6 Summary of findings table for Rimantadine versus placebo for prophylaxis against seasonal influenza

| **Outcome**  Timeframe | **Study results and measurements** | **Absolute effect estimates** | | **Certainty of the Evidence**  (Quality of evidence) | **Summary** |
| --- | --- | --- | --- | --- | --- |
|  |  | Placebo | Rimantadine |  |  |
| Lab-confirmed symptomatic influenza | Relative risk: 0.76  (CI 95% 0.28 - 2.06)  Indirect evidence | 78  per 1000 | 59  per 1000 | **Moderate**  Due to serious imprecision | Low-risk and high-risk populations: compared to placebo, rimantadine probably has little or no effect on lab-confirmed symptomatic influenza. |
|  |  | Difference: 19 fewer per 1000  (CI 95% 56 fewer - 83 more) | |  |  |
| Lab-confirmed influenza | Relative risk: 0.55  (CI 95% 0.39 - 0.78)  Based on data from 1763 participants in 8 studies | 177  per 1000 | 97  per 1000 | **Low**  Due to serious imprecision and heterogeneity | Compared to placebo/standard of care, rimantadine may decrease lab-confirmed influenza. |
|  |  | Difference: 80 fewer per 1000  (CI 95% 108 fewer - 39 fewer) | |  |  |
| Lab-confirmed asymptomatic influenza |  |  |  |  | Not applicable |
| Admission to hospital (No data from RCT) |  |  |  |  | Not applicable |
| All-cause mortality | Risk difference: 0.004  (CI 95% -0.008 – 0.015)  Based on data from 196 participants in 1 study | 0.2  per 1000 | 4.2  per 1000 | **Very low**  Due to serious incoherent and extremely serious imprecision | Whether rimantadine increases all-cause mortality is very uncertain. |
|  |  | Difference: 4.0 more  (CI 95% 8.0 fewer - 15 more) | |  |  |
| Adverse events related to drugs | Relative risk: 1.05  (CI 95% 0.79 - 1.4)  Indirect evidence | 36  per 1000 | 38  per 1000 | **Moderate**  Due to serious imprecision | Compared to placebo, rimantadine probably has little or effect on adverse events related to drugs. |
|  |  | Difference: 2 more per 1000  (CI 95% 8 fewer - 14 more) | |  |  |
| Serious adverse events | Risk difference: -0.004  (CI 95% -0.026 – 0.019)  Indirect evidence | 0.2  per 1000 | 0.2  per 1000 | **Low**  Due to serious risk of bias, Due to serious imprecision | Compared to placebo, rimantadine probably has little or no effect on serious adverse events. |
|  |  | Difference: 4 fewer per 1000  (CI 95% 26 fewer - 19 more) | |  |  |

#### Table 7 Summary of findings table for Zanamivir versus placebo for prophylaxis against zoonotic influenza

| **Outcome**  Timeframe | **Study results and measurements** | **Absolute effect estimates** | | **Certainty of the Evidence**  (Quality of evidence) | **Summary** |
| --- | --- | --- | --- | --- | --- |
|  |  | Placebo/standard of care | Zanamivir |  |  |
| Lab-confirmed symptomatic influenza | Relative risk: 0.35  (CI 95% 0.25 - 0.5)  Based on data from 8104 participants in 9 studies | 30  per 1000 | 11  per 1000 | **Low**  Due to very serious indirectness | Zanamivir may reduce lab-confirmed symptomatic influenza |
|  |  | Difference: 19 fewer per 1000  (CI 95% 22 fewer - 15 fewer) | |  |  |
| Admission to hospital (model) | Risk difference: -0.015  (CI 95% -0.018 – -0.012)  Zoonotic influenza model | 24  per 1000 | 9  per 1000 | **Very low**  Due to very serious indirectness, Due to serious imprecision | Whether zanamivir reduces admission to hospital is very uncertain. |
|  |  | Difference: 15 fewer per 1000  (CI 95% 18 fewer - 12 fewer) | |  |  |
| All-cause mortality  (model) | Risk difference: 0.003  (CI 95% -0.004 – -0.002)  Zoonotic influenza model | 5  per 1000 | 2  per 1000 | **Very low**  Due to very serious indirectness, Due to serious imprecision | Whether zanamivir reduces all-cause mortality  is very uncertain. |
|  |  | Difference: 3 fewer per 1000  (CI 95% 4 fewer - 2 fewer) | |  |  |
| Adverse events related to drugs | Relative risk: 1.07  (CI 95% 0.83 - 1.38)  Based on data from 6814 participants in 8 studies | 36  per 1000 | 39  per 1000 | **Moderate**  Due to serious imprecision | Zanamivir probably has little or no effect on the risk of adverse events related to drugs. |
|  |  | Difference: 3 more per 1000  (CI 95% 6 fewer - 14 more) | |  |  |
| Serious adverse events | Risk difference: 0  (CI 95% -0.003 - 0.004)  Based on data from 6239 participants in 7 studies | 0.2  per 1000 | 0.2  per 1000 | **High** | Zanamivir has little or no effect on the risk of serious adverse events |
|  |  | Difference: 0 fewer per 1000  (CI 95% 3 fewer - 4 more) | |  |  |

#### Table 8 Summary of findings table for oseltamivir versus placebo for prophylaxis against zoonotic influenza

| **Outcome**  Timeframe | **Study results and measurements** | **Absolute effect estimates** | | **Certainty of the Evidence**  (Quality of evidence) | **Summary** |
| --- | --- | --- | --- | --- | --- |
|  |  | Placebo | Oseltamivir |  |  |
| Lab-confirmed symptomatic influenza | Relative risk: 0.4  (CI 95% 0.26 - 0.62)  Based on data from 3742 participants in 5 studies | 30  per 1000 | 12  per 1000 | **Low**  Due to very serious indirectness | Oseltamivir may reduce lab-confirmed symptomatic influenza |
|  |  | Difference: 18 fewer per 1000  (CI 95% 22 fewer - 11 fewer) | |  |  |
| Admission to hospital (model) | Risk difference: -0.015  (CI 95% -0.018 – 0.009)  Zoonotic influenza model | 24  per 1000 | 9  per 1000 | **Very low**  Due to very serious indirectness, Due to serious imprecision. | Whether oseltamivir reduces admission to hospital is very uncertain. |
|  |  | Difference: 15 fewer per 1000  (CI 95% 18 fewer - 9 more) | |  |  |
| All-cause mortality  (model) | Risk difference: -0.003  (CI 95% -0.004 – 0.002)  Zoonotic influenza model | 5  per 1000 | 2  per 1000 | **Very low**  Due to very serious indirectness, Due to serious imprecision | Whether oseltamivir reduces all-cause mortality  mortality is very uncertain. |
|  |  | Difference: 3 fewer per 1000  (CI 95% 4 fewer - 2 more) | |  |  |
| Adverse events related to drugs (No data from RCT) |  |  |  |  | Whether oseltamivir increases adverse events related to drugs is very uncertain. |
| Serious adverse events^4^ | Risk difference: 0.003  (CI 95% -0.002 - 0.007)  Based on data from 3742 participants in 5 studies | 0.2  per 1000 | 0.2  per 1000 | **Moderate**  Due to serious imprecision | Oseltamivir has little or no effect on the risk of adverse events related to drugs |
|  |  | Difference: 3.0 more  (CI 95% 2.0 fewer - 7.0 more) | |  |  |

#### Table 9 Summary of findings table for laninamivir versus placebo for prophylaxis against zoonotic influenza

| **Outcome**  Timeframe | **Study results and measurements** | **Absolute effect estimates** | | **Certainty of the Evidence**  (Quality of evidence) | **Summary** |
| --- | --- | --- | --- | --- | --- |
|  |  | Placebo | Laninamivir |  |  |
| Lab-confirmed symptomatic influenza | Relative risk: 0.43  (CI 95% 0.3 - 0.63)  Based on data from 2593 participants in 3 studies | 30  per 1000 | 13  per 1000 | **Low**  Due to very serious indirectness | Laninamivir may reduce lab-confirmed symptomatic influenza |
|  |  | Difference: 17 fewer per 1000  (CI 95% 21 fewer - 11 fewer) | |  |  |
| Admission to hospital (model) | Risk difference: -0.014  (CI 95% -0.017 – 0.009)  Zoonotic influenza model | 24  per 1000 | 10  per 1000 | **Very low**  Due to very serious indirectness, Due to serious imprecision | Whether laninamivir reduces admission to hospital is very uncertain. |
|  |  | Difference: 14 fewer per 1000  (CI 95% 17 fewer - 9 fewer) | |  |  |
| All-cause mortality  (model) | Risk difference: -0.003  (CI 95% -0.003 – 0.002)  Zoonotic influenza model | 5  per 1000 | 2  per 1000 | **Very low**  Due to serious imprecision, Due to very serious indirectness. | Whether laninamivir reduces all-cause mortality is very uncertain. |
|  |  | Difference: 3 fewer per 1000  (CI 95% 3 fewer - 2 fewer) | |  |  |
| Adverse events related to drugs | Relative risk: 1.4  (CI 95% 0.84 - 2.35)  Based on data from 2806 participants in 3 studies | 36  per 1000 | 50  per 1000 | **Moderate**  Due to serious imprecision | Laninamivir probably increases the incidence of adverse events related to drugs |
|  |  | Difference: 14 more per 1000  (CI 95% 6 fewer - 49 more) | |  |  |
| Serious adverse events | Risk difference: 0  (CI 95% -0.011 – 0.011)  Based on data from 341 participants in 1 study | 0.2  per 1000 | 0.2  per 1000 | **Low**  Due to serious risk of bias, Due to serious imprecision^1^ | Laninamivir may have little or no effect on the risk of serious adverse events |
|  |  | Difference: 0 fewer per 1000  (CI 95% 11 fewer - 11 more) | |  |  |

#### Table 10 Summary of findings table for baloxavir versus placebo for prophylaxis against zoonotic influenza

| **Outcome**  Timeframe | **Study results and measurements** | **Absolute effect estimates** | | **Certainty of the Evidence**  (Quality of evidence) | **Summary** |
| --- | --- | --- | --- | --- | --- |
|  |  | Placebo | Baloxavir |  |  |
| Lab-confirmed symptomatic influenza | Relative risk: 0.43  (CI 95% 0.23 - 0.79)  Based on data from 749 participants in 1 study | 30  per 1000 | 13  per 1000 | **Low**  Due to very serious indirectness | Baloxavir may reduce lab-confirmed symptomatic influenza |
|  |  | Difference: 17 fewer per 1000  (CI 95% 23 fewer - 6 fewer) | |  |  |
| Admission to hospital (model) | Risk difference: -0.014  (CI 95% -0.018 - -0.005)  Zoonotic influenza model | 24  per 1000 | 10  per 1000 | **Very low**  Due to very serious indirectness, Due to serious imprecision | Whether baloxavir reduces admission to hospital is very uncertain. |
|  |  | Difference: 14 fewer per 1000  (CI 95% 18 fewer - 5 fewer) | |  |  |
| All-cause mortality  (model) | Risk difference: -0.003  (CI 95% -0.004 - -0.001)  Zoonotic influenza model | 5  per 1000 | 2  per 1000 | **Very low**  Due to serious imprecision, Due to very serious indirectness | Whether baloxavir reduces all-cause mortality is very uncertain. |
|  |  | Difference: 3 fewer per 1000  (CI 95% 4 fewer - 1 fewer) | |  |  |
| Adverse events related to drugs | Relative risk: 1.17  (CI 95% 0.4 - 3.45)  Based on data from 749 participants in 1 study | 36  per 1000 | 42  per 1000 | **Low**  Due to serious risk of bias, Due to serious imprecision | Baloxavir may have little or no difference on the incidence of adverse events related to drugs |
|  |  | Difference: 6 more per 1000  (CI 95% 22 fewer - 88 more) | |  |  |
| Serious adverse events | Risk difference: -0.003  (CI 95% -0.010 - 0.005)  Based on data from 749 participants in 1 study | 0.2  per 1000 | 0.2  per 1000 | **Moderate**  Due to serious risk of bias | Baloxavir probably not increase the risk of serious adverse events. |
|  |  | Difference: 3.0 fewer  (CI 95% 10.0 fewer - 5.0 more) | |  |  |

### Appendix 13 Assessment of global inconsistency

| Clinical outcome | P value |
| --- | --- |
| Lab-confirmed symptomatic influenza | NA |
| Lab-confirmed influenza | 0.1130 |
| Lab-confirmed asymptomatic influenza | NA |
| Admission to hospital | NA |
| All-cause mortality | 0.019 |
| Adverse events related to drugs | NA |
| Serious adverse events | NA |

### Appendix 14 Assessment of between-study heterogeneity.

| **Outcome** | **Comparison** | **No. study** | **I^2^** |
| --- | --- | --- | --- |
| Lab-confirmed symptomatic influenza | Zanamivir vs. Placebo/standard care | 9 | 43% |
|  | Oseltamivir vs. Placebo | 5 | 0% |
|  | Laninamivir vs. Placebo | 3 | 66% |
|  | Baloxavir vs. Placebo | 1 | NA |
|  | Rimantadine vs. Placebo | 0 | NA |
|  | Zanamivir vs. Rimantadine | 1 | NA |
| Lab-confirmed influenza | Zanamivir vs. Placebo/standard care | 10 | 50% |
|  | Oseltamivir vs. Placebo/standard care | 6 | 0% |
|  | Laninamivir vs. Placebo/standard care | 3 | 72% |
|  | Baloxavir vs. Placebo/standard care | 1 | NA |
|  | Amantadine vs. Placebo/standard care | 6 | 83% |
|  | Rimantadine vs. Placebo/standard care | 8 | 51% |
|  | Zanamivir vs. Rimantadine | 2 | 0% |
|  | Rimantadine vs. Amantadine | 2 | 0% |
| Lab-confirmed asymptomatic influenza | Zanamivir vs. Placebo | 9 | 0% |
|  | Oseltamivir vs. Placebo | 5 | 0% |
|  | Laninamivir vs. Placebo | 3 | 0% |
|  | Baloxavir vs. Placebo | 1 | NA |
| Admission to hospital | Oseltamivir vs. Placebo | 4 | 0% |
| Mortality | Zanamivir vs. Placebo | 5 | 0% |
|  | Oseltamivir vs. Placebo | 5 | 0% |
|  | Laninamivir vs. Placebo | 1 | NA |
|  | Baloxavir vs. Placebo | 1 | NA |
|  | Amantadine vs. Placebo | 1 | NA |
|  | Rimantadine vs. Placebo | 1 | NA |
|  | Zanamivir vs. Rimantadine | 1 | NA |
| Adverse events related to treatments | Zanamivir vs. Placebo | 8 | 0% |
|  | Laninamivir vs. Placebo | 3 | 0% |
|  | Baloxavir vs. Placebo | 1 | NA |
|  | Rimantadine vs. Placebo | 0 | NA |
|  | Zanamivir vs. Rimantadine | 1 | NA |
| Serious adverse events | Zanamivir vs. Placebo | 7 | 0% |
|  | Oseltamivir vs. Placebo | 5 | 0% |
|  | Laninamivir vs. Placebo | 1 | NA |
|  | Baloxavir vs. Placebo | 1 | NA |
|  | Amantadine vs. Placebo | 1 | NA |
|  | Rimantadine vs. Placebo | 0 | NA |
|  | Zanamivir vs. Rimantadine | 1 | NA |

NA, not applicable.

### Appendix 15 Funnel plot

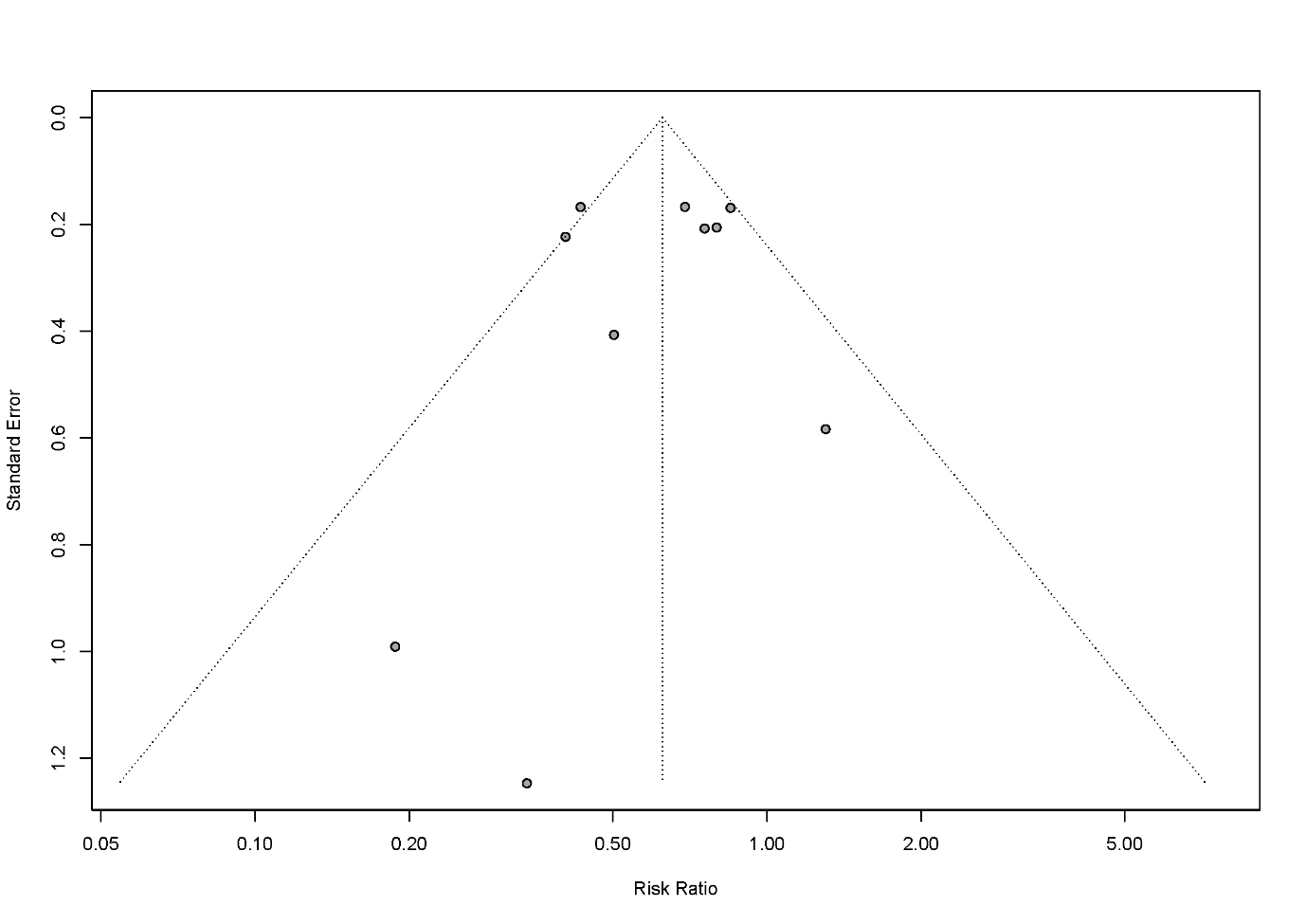

#### Figure 1 Funnel plot zanamivir vs placebo/ standard care for lab-confirmed influenza (Harbord test p = 0.77).

### Appendix 16 Sensitivity analysis by only including the trials with lower risk of bias

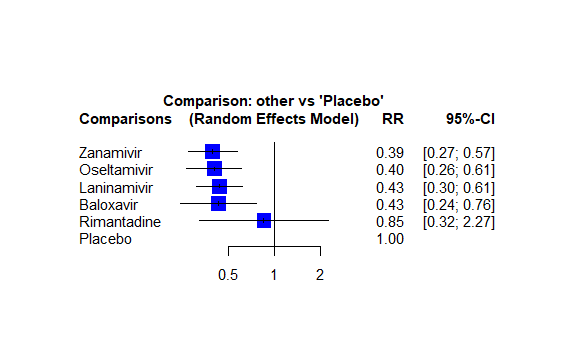

#### Figure 1 Sensitivity analysis for antivirals prophylaxis against lab-confirmed symptomatic influenza by only including the trials with lower risk of bias.

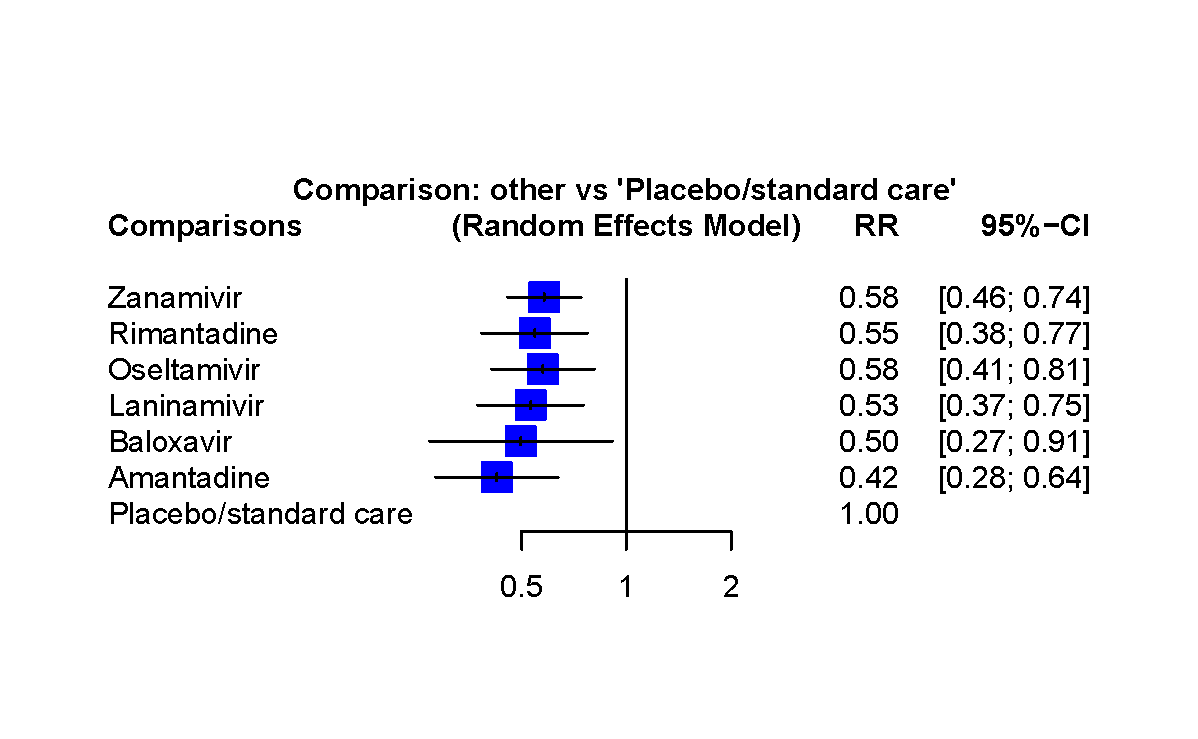

#### Figure 2 Sensitivity analysis for antivirals prophylaxis against lab-confirmed influenza by only including the trials with lower risk of bias.

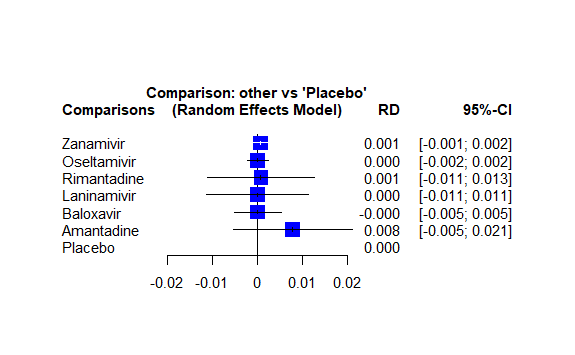

#### Figure 3 Sensitivity analysis for antivirals prophylaxis against mortality by only including the trials with lower risk of bias.

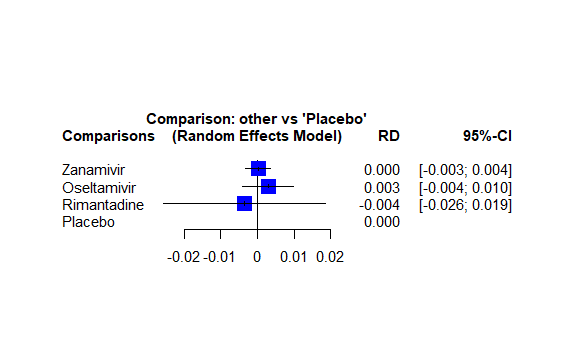

#### Figure 4 Sensitivity analysis for antivirals prophylaxis against serious adverse events by only including the trials with lower risk of bias.

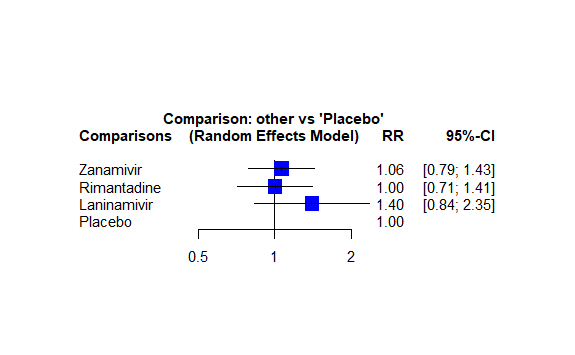

#### Figure 5 Sensitivity analysis for antivirals prophylaxis against adverse events related to drugs by only including the trials with lower risk of bias.

### Appendix 17 Plausible worst-case sensitivity analyses

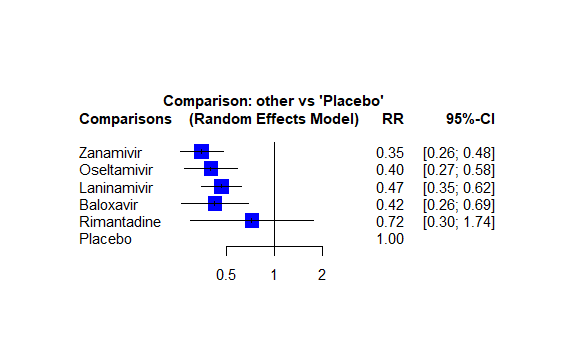

#### Figure 1 Plausible worst-case sensitivity analyses for antivirals prophylaxis against lab-confirmed symptomatic influenza.

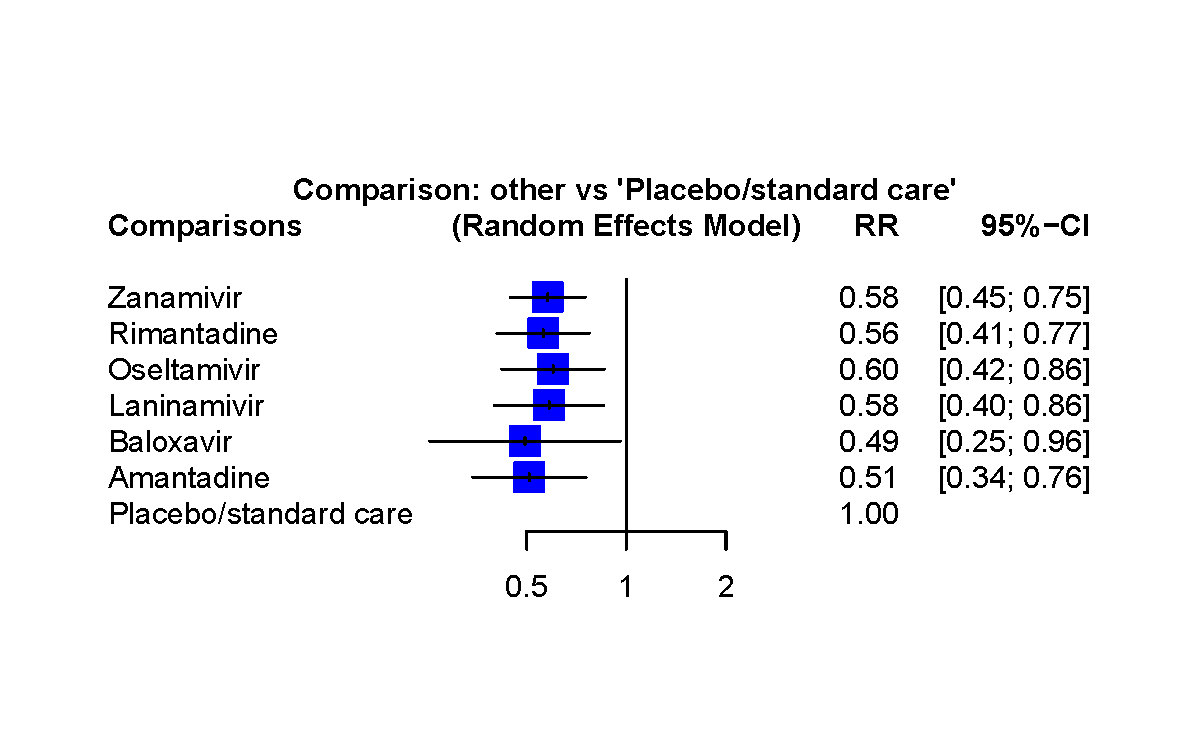

#### Figure 2 Plausible worst-case sensitivity analyses for antivirals prophylaxis against lab-confirmed influenza.

### Appendix 18 Sensitivity analysis according to the evidence of exposure to influenza

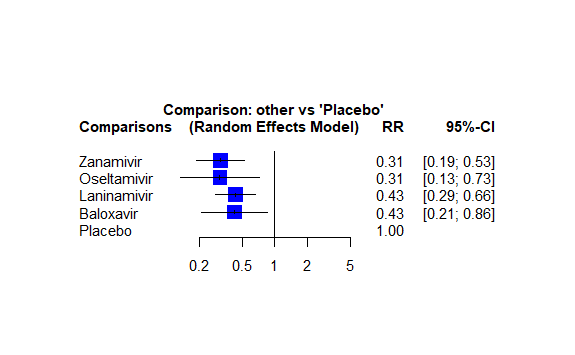

#### Figure 1 Sensitivity analysis for antivirals prophylaxis against lab-confirmed symptomatic influenza by including the trials with clear evidence of exposure to influenza.

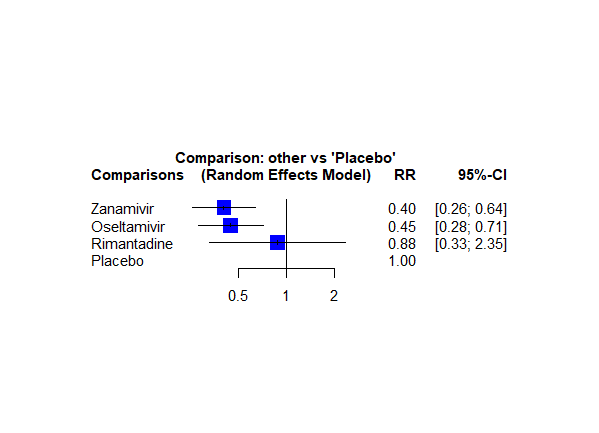

#### Figure 2 Sensitivity analysis for antivirals prophylaxis against lab-confirmed symptomatic influenza by excluding the trials with clear evidence of exposure to influenza.

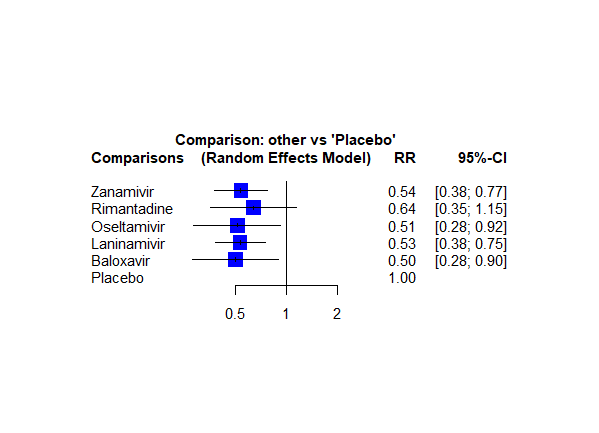

#### Figure 3 Sensitivity analysis for antivirals prophylaxis against lab-confirmed influenza by including the trials with clear evidence of exposure to influenza.

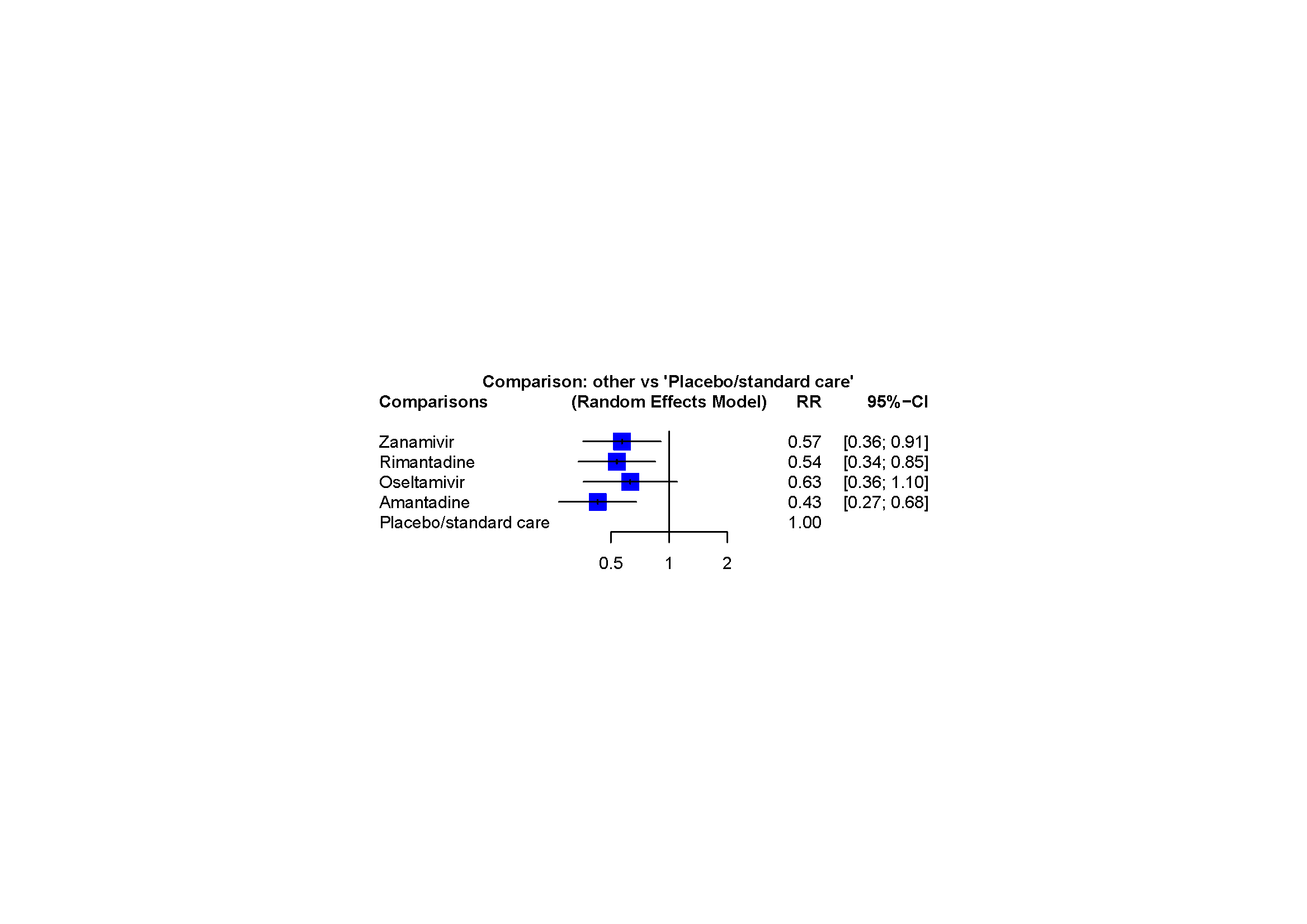

#### Figure 4 Sensitivity analysis for antivirals prophylaxis against lab-confirmed influenza by including the trials with less evidence of exposure to influenza.

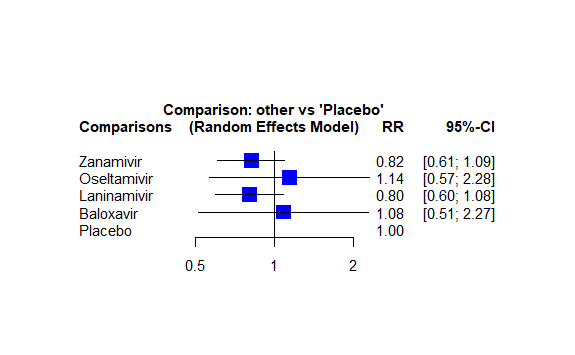

#### Figure 5 Sensitivity analysis for antivirals prophylaxis against lab-confirmed asymptomatic influenza by including the trials with clear evidence of exposure to influenza.

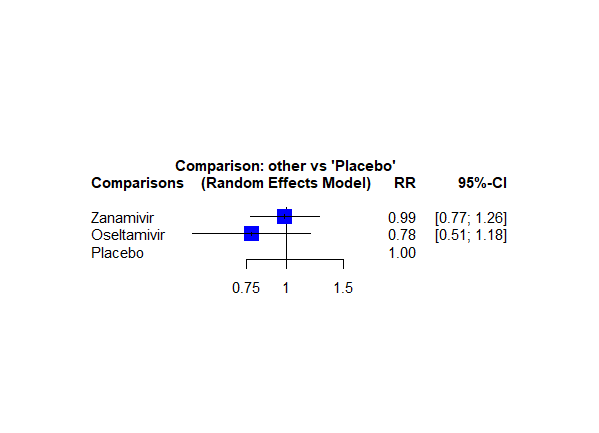

#### Figure 6 Sensitivity analysis for antivirals prophylaxis against lab-confirmed asymptomatic influenza by excluding the trials with clear evidence of exposure to influenza.

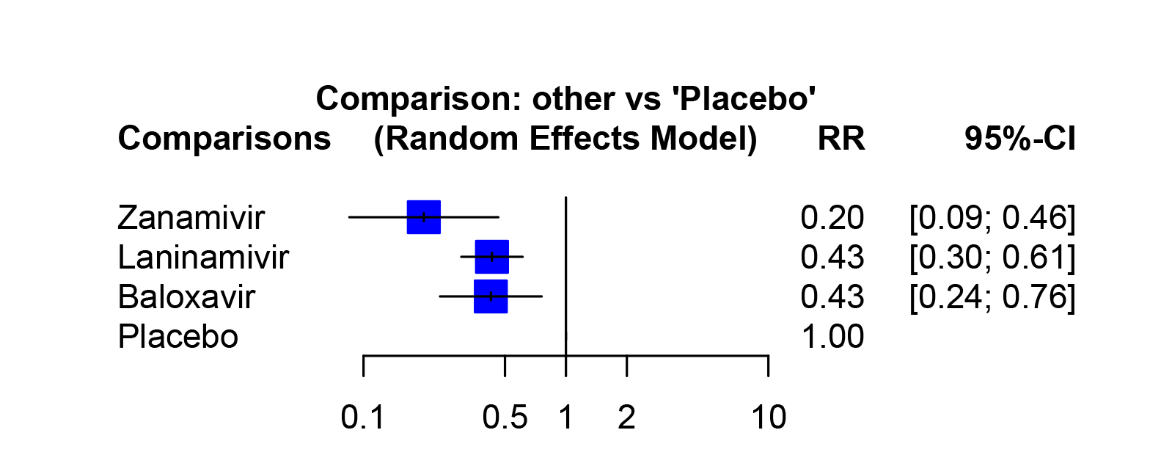

#### Figure 7 Sensitivity analysis for antivirals prophylaxis against lab-confirmed symptomatic influenza by only including the trials with close contact with the index with confirmed influenza.

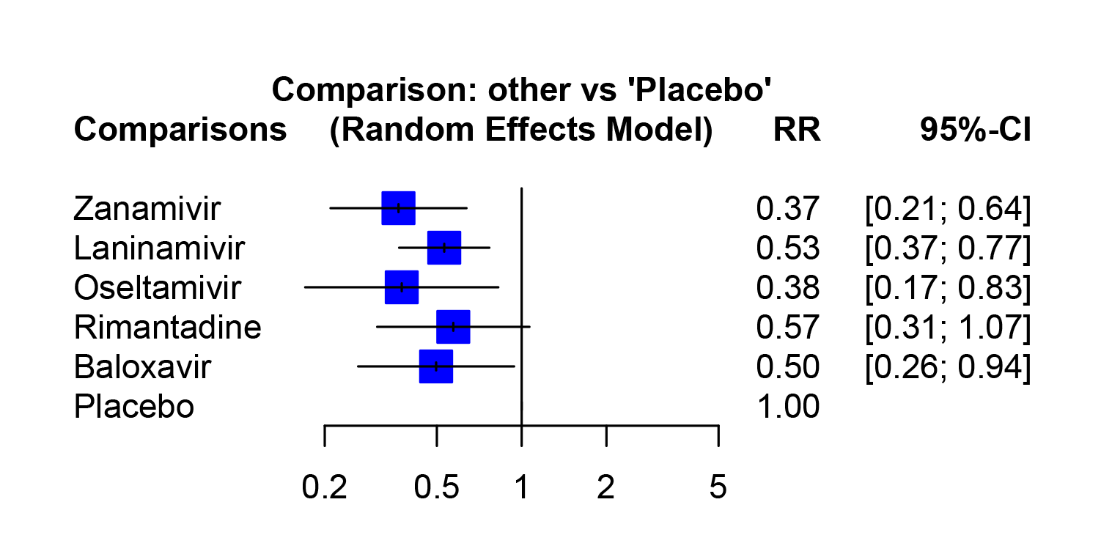

#### Figure 8 Sensitivity analysis for antivirals prophylaxis against lab-confirmed influenza by only including the trials with close contact with the index with confirmed influenza.

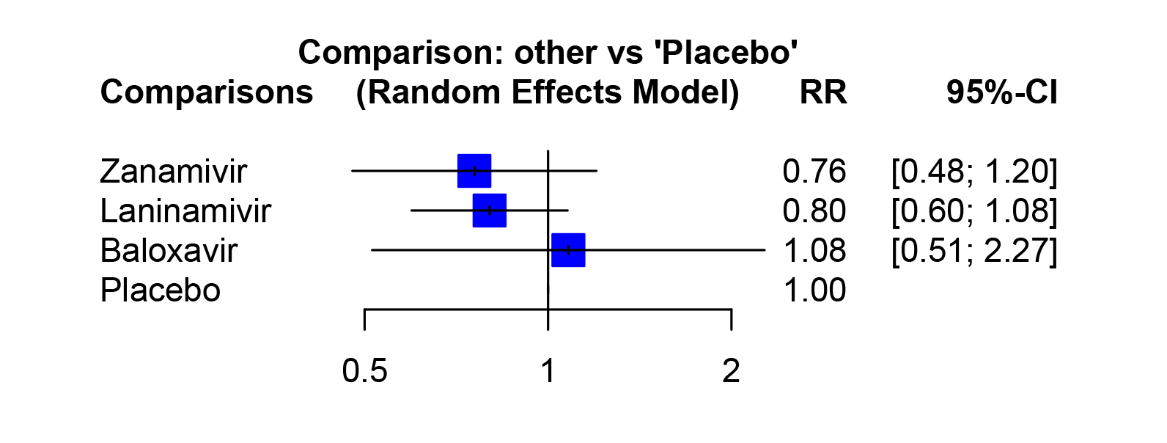

#### Figure 9 Sensitivity analysis for antivirals prophylaxis against lab-confirmed asymptomatic influenza by only including the trials with close contact with the index with confirmed influenza.

### Appendix 19 Sensitivity analysis by using a higher ICC (0.10) to adjust cluster design effect.

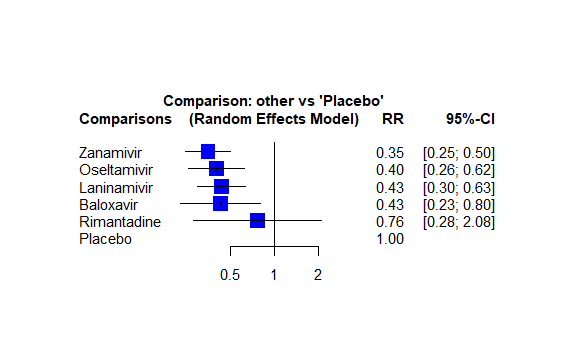

#### Figure 1 Sensitivity analysis for antivirals prophylaxis against lab-confirmed symptomatic influenza (ICC = 0.1).

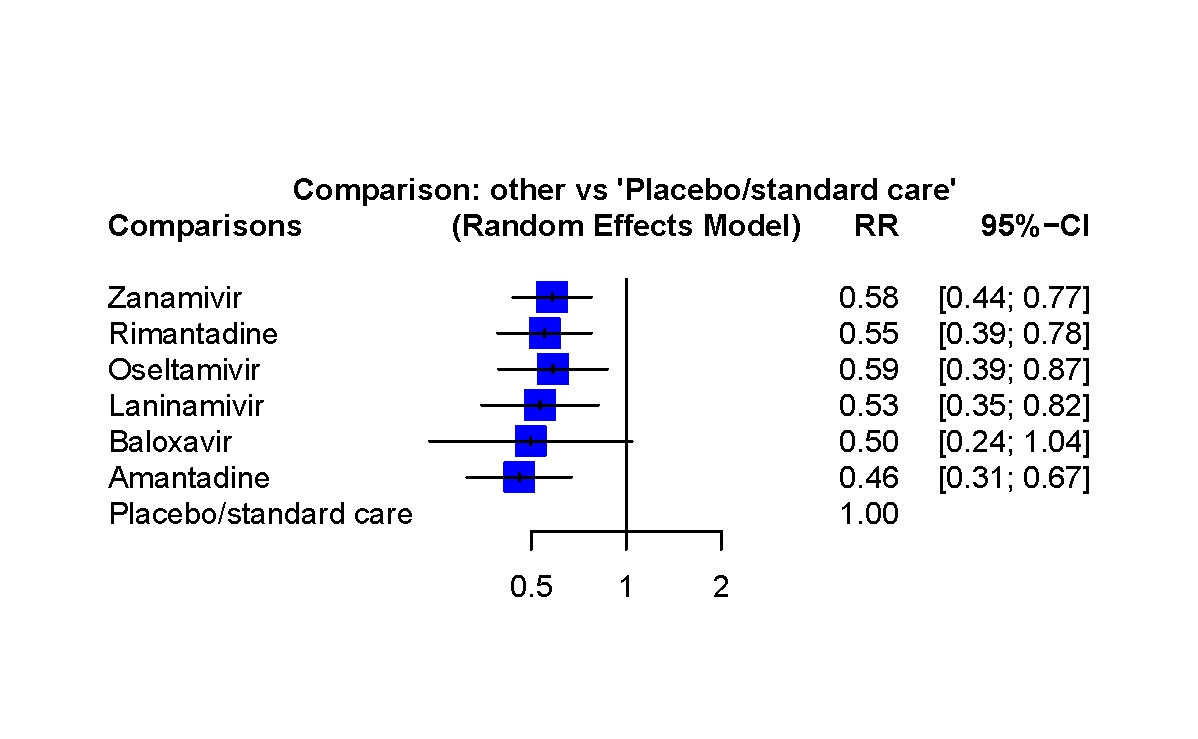

#### Figure 2 Sensitivity analysis for antivirals prophylaxis against lab-confirmed influenza (ICC = 0.1).

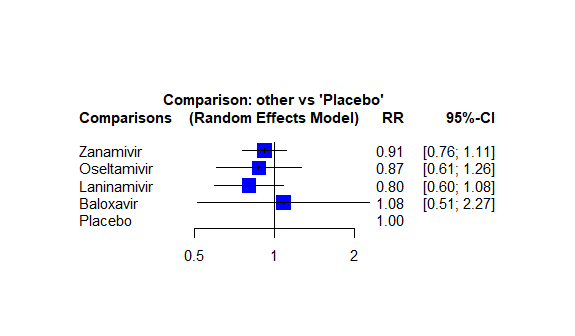

#### Figure 3 Sensitivity analysis for antivirals prophylaxis against lab-confirmed asymptomatic influenza (ICC = 0.1).

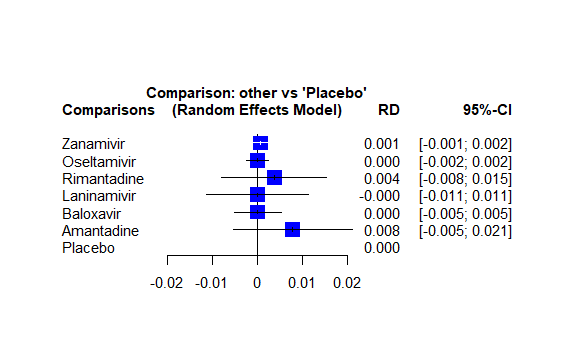

#### Figure 4 Sensitivity analysis for antivirals prophylaxis against all-cause mortality (ICC = 0.1).

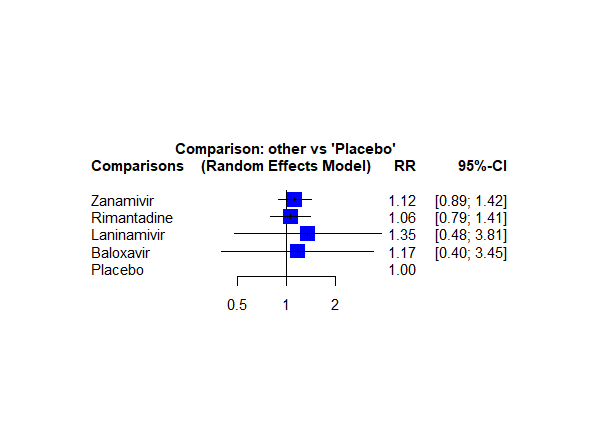

#### Figure 5 Sensitivity analysis for antivirals prophylaxis against serious adverse events (ICC = 0.1).

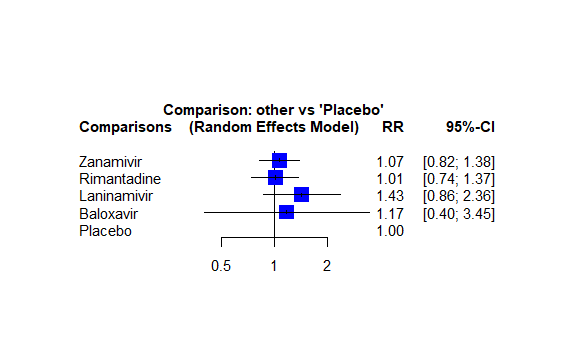

#### Figure 6 Sensitivity analysis for antivirals prophylaxis against adverse events related to drugs (ICC = 0.1).

### Appendix 20 Subgroup analysis

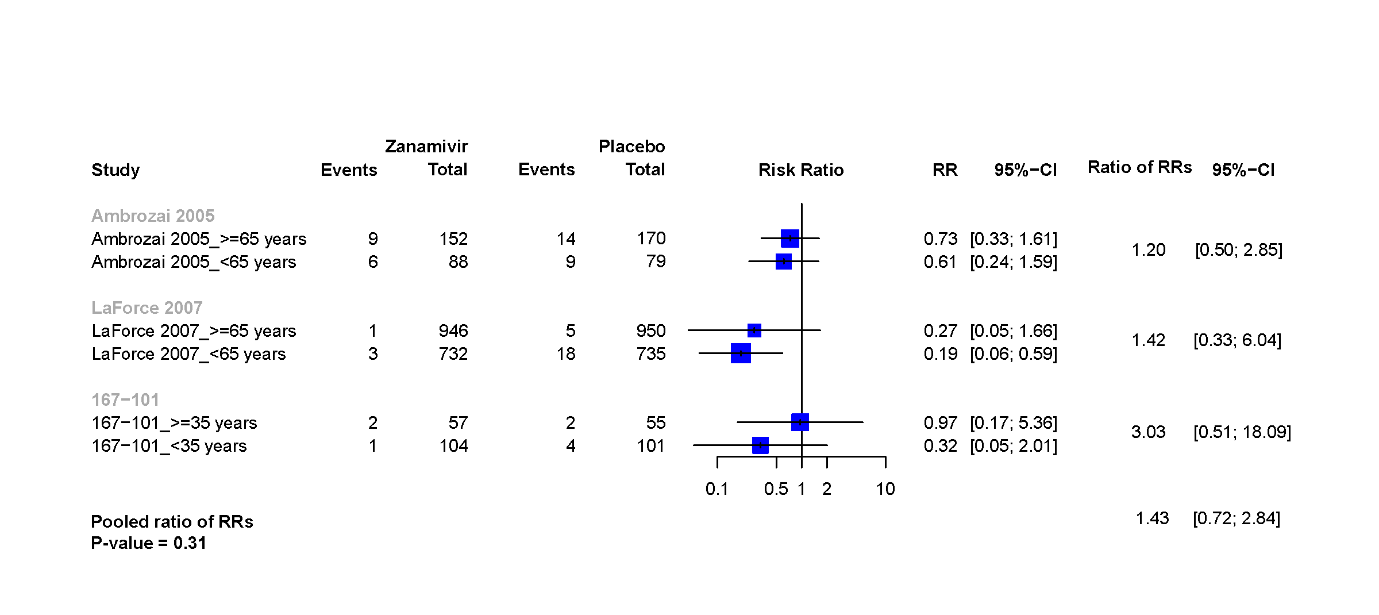

#### Figure 1 Within-subgroup analysis of lab-confirmed influenza according to age (zanamivir versus placebo)

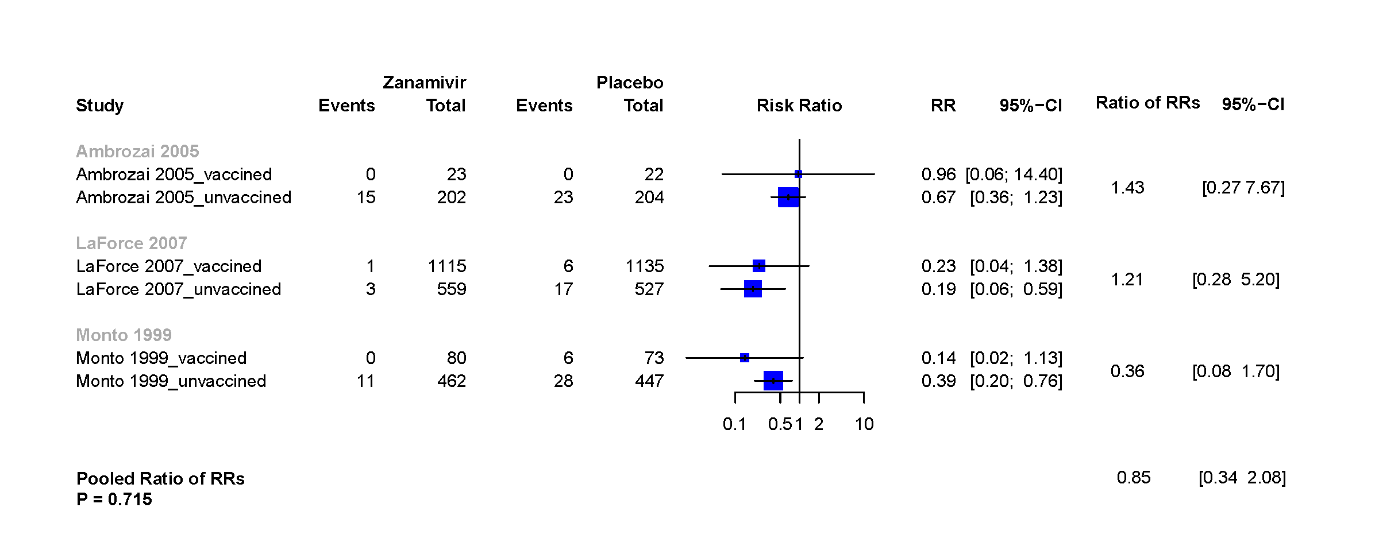

#### Figure 2 Within-subgroup analysis of lab-confirmed influenza according to vaccination status (zanamivir versus placebo)

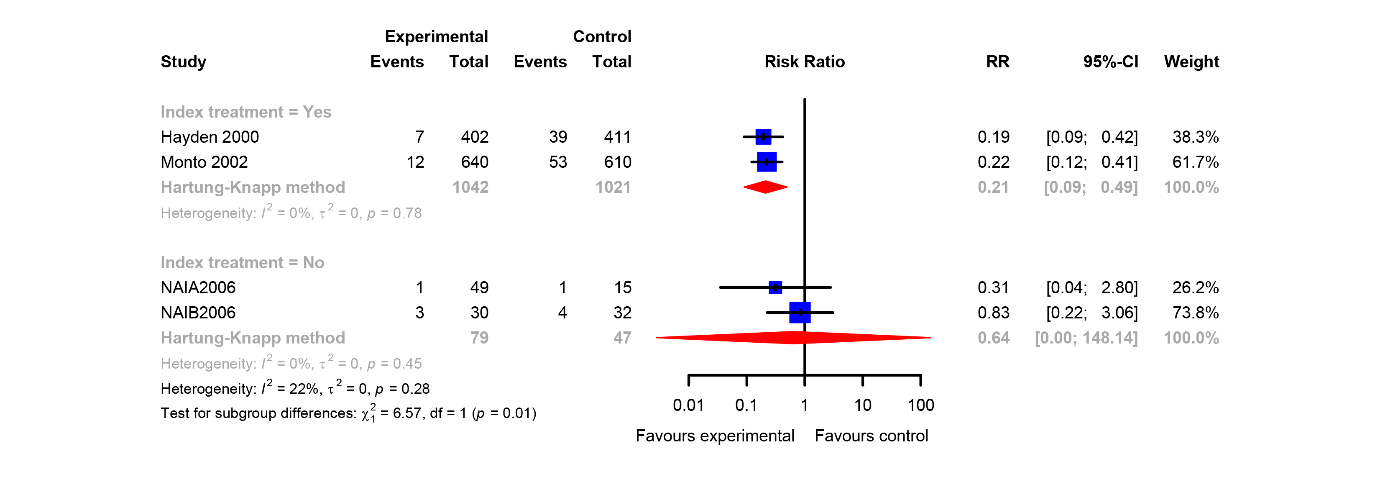

#### Figure 3 Within-subgroup analysis of lab-confirmed influenza according to the status of index patients treatment (zanamivir versus placebo)

### Appendix 21: Credibility assessment of subgroup analysis

#### Text 1 Credibility assessment of treatment of index patients

| **Credibility assessment** | | | |
| --- | --- | --- | --- |
| **1: Is the analysis of effect modification based on comparison within rather than between trials?** | | | |
| [**X** ] Completely between | [ ] Mostly between or unclear | [ ] Mostly within | [ ] Completely within |
| *Subgroup analysis or meta-regression comparing overall effects of each individual trial. This is typical for aggregate data meta-analysis.* | *Subgroup analysis or meta-regression with most information coming from overall effects, but some trials providing within-trial subgroup information* | *Most trials providing within-trial subgroup information; or individual participant data analysis that combines within and between trial information* | *All trials providing within-trial subgroup information or individual participant data; and the analysis separates within from between trial information, e.g., meta-analysis of interactions* |
| **2: For within-trial comparisons, is the effect modification similar from trial to trial?** [**X**] Not applicable: no or one within-RCT comparison | | | |
| [ ] Definitely not similar | [ ] Probably not similar or unclear | [ ] Mostly similar | [ ] Definitely similar |
| *Effect modification reported for two or more trials and clearly different directions* | *Effect modification not reported for individual trials or too imprecise to tell* | *Effect modification reported for two or more trials, mostly similar in direction, but considerable differences in magnitude* | *Effect modification reported for two or more trials, similar in direction, only some differences in magnitude* |
| Comment: | | | |
| **3: For between-trial comparisons, is the number of trials large?** [ ] Not applicable: no between RCT comparison | | | |
| [**X** ] Very small | [ ] Rather small or unclear | [ ] Rather large | [ ] Large |
| *1 or 2 or in smallest subgroup; 5 or less in continuous meta-regression* | *3-4 in smallest subgroup; 6-10 in continuous meta-regression* | *5-9 in smallest subgroup; 11 to 15 in continuous meta-regression* | *10 or more in smallest subgroup; more than 15 in continuous meta-regression* |
| Comment: | | | |
| **4: Was the direction of effect modification correctly hypothesized a priori?** | | | |
| [ **X** ] Definitely no | [ ] Probably no or unclear | [ ] Probably yes | [ ] Definitely yes |
| *Clearly post-hoc or results inconsistent with hypothesized direction or biologically very implausible* | *Vague hypothesis or hypothesized direction unclear* | *No prior protocol available but unequivocal statement of a priori hypothesis with correct direction of effect modification* | *Prior protocol available and includes correct specification of direction of effect modification, e.g., based on a biologic rationale* |
| Comment: We didn’t hypothesize a priori in the protocol. | | | |
| **5: Does a test for interaction suggest that chance is an unlikely explanation of the apparent effect modification?** (consider irrespective of number of effect modifiers) | | | |
| [ ] Chance a very likely explanation | [ ] Chance a likely explanation or unclear | [**X** ] Chance may not explain | [ ] Chance an unlikely explanation |
| *Interaction or meta-regression p-value >0.05* | *Interaction or meta-regression p-value ≤0.05 and >0.01, or no test of interaction reported and not computable* | *Interaction or meta-regression p-value ≤0.01 and >0.005* | *Interaction or meta-regression p-value ≤0.005* |
| Comment: The interaction P-value = 0.01. | | | |
| **6: Did the authors test only a small number of effect modifiers or consider the number in their statistical analysis?** | | | |
| [ ] Definitely no | [ ] Probably no or unclear | [**X** ] Probably yes | [ ] Definitely yes |
| *Explicitly exploratory analysis or large number of effect modifiers tested (e.g., greater than 10) and multiplicity not considered in analysis* | *No mention of number or 4-10 effect modifiers tested and number not considered in analysis* | *No protocol available but unequivocal statement of 3 or fewer effect modifiers tested* | *Protocol available and 3 or fewer effect modifiers tested or number considered in analysis* |
| Comment: Three effect modifiers were tested in this review. | | | |
| **7: Did the authors use a random effects model?** [ ] Not applicable | | | |
| [ ] Definitely no | [ ] Probably no or unclear | [ ] Probably yes | [ **X** ] Definitely yes |
| *Fixed (or common) effect or fixed effects model explicitly stated* | *Probably fixed effect(s) model* | *Probably random (or mixed) effects* | *Random (or mixed) effects explicitly stated* |
| Comment: | | | |
| **8: If the effect modifier is a continuous variable, were arbitrary cut points avoided?** [**X** ] not applicable: not continuous | | | |
| [ ] Definitely no | [ ] Probably no or unclear | [ ] Probably yes | [ ] Definitely yes |
| *Analysis based on exploratory cut point(s), e.g., picking cut point associated with highest interaction p-value* | *Analysis based on cut point(s) of unclear origin* | *Analysis based on pre-specified cut point(s), e.g., suggested by prior RCT* | *Analysis based on the full continuum, e.g., assuming a linear or logarithmic relationship* |
| **9 Optional: Are there any additional considerations that may increase or decrease credibility?** (manual section 3.9) [ **X** ] not applicable | | | |
|  | [ ] Yes, probably decrease  Biologically implausible  Expect similar severe critical  Opposite effects unlikely | [ ] Yes, probably increase | |
| Comment: **10: How would you rate the overall credibility of the proposed effect modification?** The overall rating should be driven by the items that decrease credibility. The following provides a sensible strategy: - All responses definitely or probably decrease credibility or unclear 🡪 very low - Two or more responses definitely decrease credibility 🡪 maximum usually low even if all other responses satisfy credibility criteria - One response definitely decreases credibility 🡪 maximum usually moderate even if all other responses satisfy credibility criteria - Two responses probably decrease credibility 🡪 maximum usually moderate even if all other responses satisfy credibility criteria - No response options definitely or probably decrease credibility 🡪 high very likely Place a mark on the continuous line (or type “x” in editable version) \| \| \| \| \|; \| **X** \| \| \| \|; \| **Very low credibility** \| **Low credibility** \| **Moderate credibility** \| **High credibility** \|; \| Very likely no effect modification Use overall effect for each subgroup \| Likely no effect modification Use overall effect for each subgroup but note remaining uncertainty \| Likely effect modification Use separate effects for each subgroup but note remaining uncertainty \| Very likely effect modification Use separate effects for each subgroup \|; Comment: \| \| \| \| \| | | | |
